# Using routine clinical features to classify adult-onset diabetes at diagnosis: the StartRight prospective observational study

**DOI:** 10.64898/2026.05.01.26352217

**Authors:** Julieanne Knupp, Anita Hill, Nicholas JM Thomas, Timothy J McDonald, Katherine G Young, Diane P Fraser, Andrew T Hattersley, Trevelyan J McKinley, Beverley M Shields, Angus G Jones

**Affiliations:** Department of Clinical and Biomedical Sciences, University of Exeter, RILD Building North, Barrack Road, Exeter, EX2 5GL; Academic Department of Blood Sciences, Royal Devon University Healthcare NHS Foundation Trust, Exeter EX25DW, UK

**Author notes:** **Corresponding author:** Angus G Jones, Department of Clinical and Biomedical Sciences, University of Exeter, RILD Building North, Barrack Road, Exeter, EX2 5GL.

## Abstract

**Background:** It is not known which clinical features optimally differentiate type 1 and 2 diabetes at diagnosis. We aimed to determine which clinical features differentiate these conditions at diagnosis and develop classification models combining clinical features with and without islet-autoantibodies.

**Methods:** In this prospective cohort study, we recruited 1,800 adults (aged ≥18 years) diagnosed with diabetes within the preceding 12 months, excluding secondary or monogenic diabetes. The primary outcome was diabetes subtype defined by a combination of insulin treatment and endogenous insulin production (measured by C-peptide) at ≥3 years post-diagnosis. Models were developed in participants aged 18–50 years and validated internally, alongside validation in an older cohort (aged >50 years) and using UK primary care data (n=188,232).

**Results:** Eleven clinical features and routinely measured biomarkers discriminated type 1 from type 2 diabetes independently of diagnosis age and BMI. Lower age of diagnosis, BMI and waist-hip ratio, unintentional weight-loss, and higher presentation HbA1c or glucose were the most discriminative, with other features only weakly discriminative. Models integrating routine features with and without islet-autoantibodies, developed in those age 18–50 years at diabetes diagnosis, had high performance in internal validation (clinical features only: Area Under the Receiver Operating Characteristic curve (AUCROC) (95% CI) 0.94 (0.93, 0.96), clinical features and islet-autoantibodies: AUCROC 0.97 (0.96, 0.98), and maintained high discrimination in older adults (age >50 years AUCROC 0.93 (0.90, 0.96), and 0.97 (0.94, 0.99). Simplifying the models to a points-based score resulted in similar performance. In primary care data models and score were strongly predictive of outcomes associated with type 1 diabetes, including in those initially treated as type 2.

**Interpretation:** Lower age-at-diagnosis, BMI, and wait-hip ratio, unintentional weight loss and high presentation glycaemia are the most discriminative features for diagnosis of type 1 diabetes in adults. Models combining routine clinical features, with or without islet-autoantibodies, have high accuracy and could assist clinical classification and prioritisation of classification biomarker testing.

**Funding:** UK National Institute of Health and Care Research (NIHR) and Diabetes UK.

**Research in context:** *Evidence before this study:* Most type 1 diabetes occurs in adults, but differentiating it from type 2 diabetes, which is much more common, is challenging, and misclassification is common. Two systematic reviews, published in 2015 (Shields et al.) and 2022 (UK National Institute for Health and Care Excellence) have identified that age at diagnosis and BMI are the only clinical features available at diagnosis which robustly discriminate type 1 and 2 diabetes, based on cross sectional studies, with rapid progression to insulin after diagnosis also discriminative. Many other features included in textbooks and guidelines have little supporting evidence. Guideline bodies, including the UK National Institute for Health and Care Excellence (NICE), have therefore identified the need for evidence on what features discriminate type 1 and 2 diabetes and how they can be combined. We repeated a search of PubMed and Google Scholar for articles published since the NICE review (1^st^ January 2021 to 3rd June 2026), alongside citation search of previous key articles, and identified no additional studies.

*Added value of this study:* This is the first study to prospectively assess utility of clinical features for diabetes subtype diagnosis and the first to develop classification models for adult-onset type 1 and 2 diabetes at diagnosis. The five most discriminative routine clinical features for distinguishing type 1 from type 2 diabetes at diagnosis are age-at-diagnosis, BMI, waist-hip ratio, pre-diagnosis unintentional weight-loss, and presentation glycaemia (HbA1c or glucose). Many features included in current guidelines were only very weakly discriminative of subtype, and no single clinical feature was able to adequately differentiate between type 1 and type 2 diabetes alone. A clinical prediction model combining nine routinely available clinical features, with or without islet-autoantibodies, as both a prototype calculator and a points-based score (the StartRight Score), had high accuracy in differentiating type 1 from type 2 diabetes and outperforms current clinical guidance and islet-autoantibody assessment alone.

*Implications of all available evidence:* Differentiating type 1 and 2 diabetes at diagnosis of adult-onset diabetes is challenging and misclassification common. Lower age-at-diagnosis, BMI, and wait-hip ratio, unintentional weight loss and high presentation glycaemia are the most discriminative features for diagnosis of adult-onset type 1 diabetes but are inadequate in isolation. Models combining clinical features with or without islet autoantibodies have potential to assist clinical classification and prioritisation of classification biomarker testing.

## Introduction

The vast majority of adults developing diabetes will have type 1 (T1D) or 2 diabetes (T2D). These conditions have very different treatment guidelines, primarily due to the rapid and severe endogenous insulin deficiency that occurs in type 1 diabetes as a result of autoimmunity against insulin-secreting pancreatic beta cells.^1–4^ Contrary to common belief most T1D develops in adults.^5,6^ However, diagnosis in this age group can be challenging: T2D is far more common, and discriminative features and biomarkers overlap between these conditions at diagnosis.^2,7^ Consequently, misclassification is common, with approximately one in three adults developing T1D initially diagnosed and treated as T2D, and one in six adults diagnosed as T1D having misclassified T2D on detailed investigation.^7–10^

Despite the high global prevalence of diabetes there is limited robust evidence on which clinical features should be used to differentiate T1D and T2D. A systematic review in 2015 demonstrated that age-at-diagnosis, BMI, and early insulin requirement were the only evidence-based clinical features, with other features commonly included in textbooks and guidelines either not addressed or non-discriminative.^11^ This was further confirmed by systematic review by the UK National Institute of Health and Care Excellence (NICE) in 2022,^12^ who recommended research to determine which clinical features differentiate T1D and T2D, and how they can best be combined, as a high priority for patient care.^1^ While classification biomarker testing such as islet-autoantibodies and C-peptide can improve subtype diagnosis, C-peptide (a marker of insulin secretion) has limited utility at diagnosis, and costs preclude routine islet-autoantibody testing in all those developing diabetes.^9,13^ In addition, overlap of islet-autoantibodies between T1D and T2D means interpretation depends on prior likelihood.^7,14,15^ Therefore, we aimed to: 1) identify routinely available presentation features that robustly differentiate between adult-onset T1D and T2D at diagnosis, and 2) develop classification models that combine routine features, with or without classification biomarkers to assist classification of new adult-onset diabetes.

## Methods

We undertook a prospective study to determine the relationship between routine clinical features and biomarkers at diabetes diagnosis, and diabetes subtype confirmed by assessment of endogenous insulin secretion (C-peptide) after ≥3 years diabetes duration. We developed and assessed the performance of classification models combining discriminatory features with or without classification biomarkers (islet autoantibodies and/or a type 1 diabetes genetic risk score (T1DGRS)) using this dataset, with additional validation undertaken in primary healthcare records. This study adhered to the TRIPOD-AI reporting (Appendix p.3) and STROBE guidelines.

### Study design

We prospectively studied participants aged ≥18 years, diagnosed with diabetes in the previous 12 months, who were followed annually for ≥3 years from recruitment (ClinicalTrials.gov ID: NCT03737799). Exclusion criteria were a diagnosis of gestational diabetes, monogenic diabetes, or secondary diabetes (considered likely due to another condition, e.g. exocrine pancreatic disease). The study consisted of two cohorts. The primary study (StartRight) recruited participants aged 18–50 years (inclusive) at diabetes diagnosis, regardless of treatment, from year 2015-2020. A second, separately funded, cohort (StartRight Prime) recruited participants aged >50 at diabetes diagnosis between years 2017-2020. To ensure sufficient numbers of participants with T1D (rare in comparison to T2D in older adults), recruitment of StartRight Prime was enriched for T1D by aiming for equal numbers of participants treated with and without insulin at study recruitment at the site level. Because of this enrichment, analysis of the two cohorts is presented separately. A study flow diagram is shown in Appendix p5. The study was approved by the South West (Cornwall and Plymouth) NHS Research Ethics Committee ref: 16/SW/0130. All participants gave written informed consent.

### Data collection

Participants were identified from primary and secondary care and enrolled at 55 UK NIHR Research Delivery Network sites. At recruitment, diabetes presentation characteristics (unintentional weight-loss, osmotic symptoms (nocturia, polyuria, thirst), parental history of diabetes (assessed separately as insulin- and non-insulin-treated), ethnicity, and personal and parental history of autoimmune disease were self-reported. Acanthosis Nigricans was recorded based on examination. Hypertension was recorded based on self-report confirmed by current blood pressure lowering medication. Glycaemia (HbA1c, glucose), weight, and ketoacidosis at diabetes presentation were determined by reviewing participants medical notes and laboratory records. Diabetic ketoacidosis (DKA) was defined based on the Joint British Diabetes Societies guidelines:^16^ In the absence of an available pH measurement, cases were included as DKA if DKA was recorded in the hospital notes alongside a supportive blood/urine ketone value. Participants not admitted to hospital were assumed not to have had DKA. Recruitment height, weight, and waist and hip circumference, were measured by the recruiting researcher and non-fasted (within 1–5 hours of a carbohydrate containing meal) blood was collected for measurement of glucose, islet-autoantibodies (GAD, IA2, and ZnT8), and DNA extraction.

Participants were followed annually with self-report of diabetes treatment and HbA1c recorded from laboratory records. At the final visit, and ≥3 years from diabetes diagnosis, participants’ blood C-peptide was assessed 1–5 hours after a meal on an EDTA sample collected in person, or capillary sample collected at home using the Neoteryx Mitra micro-sampling device (Trajan Scientific), alongside concurrent glucose (fluoride or capillary).

### Definition of type 1 and 2 diabetes

The primary diabetes subtype definition was based on insulin requirement and C-peptide at ≥3 years post-diagnosis:

- Type 1 diabetes: continuous insulin treatment within 3-years of diagnosis and C-peptide <600pmol/L at ≥3 years diabetes duration.
- Type 2 diabetes: treatment without insulin at ≥3 years from diagnosis or, where insulin treated, C-peptide at ≥3 years diabetes duration ≥600pmol/L.

Sensitivity analysis used a restrictive secondary definition of diabetes subtype including autoantibody assessment (Appendix p6):

- Type 1 diabetes: continuous insulin treatment within 3-years of diagnosis and, either C-peptide <200pmol/L or 2+ positive islet-autoantibodies.
- Type 2 diabetes: 3 negative islet-autoantibodies combined with either lack of insulin treatment at ≥3 years diabetes duration or, if on insulin, C-peptide at ≥3 years diabetes duration ≥600pmol/L

### Laboratory analysis

Biochemical markers, including C-peptide and islet-autoantibodies (GAD, IA2, ZNT8) were analysed by the academic Blood Sciences Department at the Royal Devon University Healthcare NHS Foundation Trust. Islet-autoantibodies were measured using ELISA assays (RSR Limited, Cardiff, U.K.) on a DYNEX DS2 automated ELISA system (Launch Diagnostics, Longfield, U.K) and considered positive if exceeding the 97·5^th^ centile of 1,559 control subjects without diabetes: GAD ≥11 World Health Organization (WHO) units/mL, IA2 ≥7·5 WHO units/mL, ZNT8 ≥65 units/mL age <30 years and ≥10 units/mL age ≥30 years.^9,17,18^ Specificity for all three assays was 99% in the 2020 International Islet Autoantibody Standardization Program Exeter Laboratory certification, with sensitivity of 74% for GAD and ZNT8, and 72% for IA2.

C-peptide was measured using an electrochemiluminescence immunoassay on a Roche Diagnostics E170 analyser (Roche, Mannheim, Germany; limit of detection 3·3pmol/L; inter- and intra-assay coefficients of variation <4·5% and <3·3%, respectively). Capillary C-peptide was reported with a limit of detection of 200pmol/L.

### Generation of Type 1 genetic risk score

A type 1 diabetes genetic risk score (T1DGRS) was calculated using 67 published T1D associated variants as previously reported.^19^

### Statistical analysis

All analysis was undertaken in R version 4·4·0 (R Core Team, 2024). All statistical code for this study is available on https://github.com/Exeter-Diabetes/T1DvsT2D_atDiagnosis_adults

#### Establishing clinical features that differentiate type 1 and 2 diabetes

We used logistic regression to assess whether routinely available presentation clinical features, selected based on previous literature, improved differentiation of T1D and T2D independently of age-at-diagnosis and BMI.

Presentation clinical features that added statistically significantly to age-at-diagnosis and BMI were examined separately in complete case data in StartRight, with individual performance of these features presented based on univariate analysis against the primary study outcome. Diagnostic performance was assessed using area under the Receiver Operating Characteristic curve (AUCROC), and sensitivity, specificity, positive predictive values (PPV), negative predictive values (NPV), and accuracy were calculated at the identified optimal threshold for identifying T1D, based on the Youden’s Index.^20^ We then repeated this analysis to assess the performance of these features for discrimination in StartRight Prime (diagnosis age >50 years) and against the restricted secondary outcome in both cohorts.

#### Building a clinical prediction model for diabetes classification

Logistic regression was used to build models for predicting T1D versus T2D defined by the primary study outcome. Initial models were based on routinely available features independent of age-at-diagnosis and BMI in the above analysis, informed by clinical opinion of whether the measure represented a barrier to clinical implementation. Subsequent models were developed for combinations of routine features with classification markers that are not routinely measured and incur additional cost: islet-autoantibodies (GAD/IA2/ZNT8), and/or T1DGRS. Continuous variables were standardised and associations with the primary outcome were assessed for linearity through visual inspection. Interactions were assessed between clinical features considered biologically plausible (age-at-diagnosis and all of BMI, parental diabetes, previous autoimmune disease; sex and both BMI and previous autoimmune disease). Internal validation was performed by using 100 bootstraps in the development data to assess model prediction stability and 1,000 bootstraps to assess optimism.^21,22^ Models were compared with using islet-autoantibodies alone and T1DGRS alone. Discrimination was assessed through AUCROC and Precision-Recall (PR) curve analysis. Youden’s Index^20^ was used to identify optimal thresholds. Model calibration was assessed using calibration curves, the Brier score, calibration-in-the-large and calibration slope. Using the approach of Riley et al.^23^ (‘pmsampsize’ package in R), based on a C-statistic of 0·80, 11 model parameters, and 25% outcome prevalence to achieve small overfitting (expected shrinkage of predictor effects <10%) and estimation of outcome within a small margin of error (+/-0·05), a sample size of 420 is needed for prediction model development, well within the number of participants included in analysis.

##### Model performance in StartRight Prime

The performance of all developed models were assessed in StartRight Prime (diagnosis age >50 years) using model objects developed in StartRight.

##### Subgroup and sensitivity analysis

Models were tested (in StartRight and StartRight Prime separately) in subgroups defined by islet-autoantibody status, clinical diagnosis at recruitment, sex, and self-reported ethnicity.

The clinical features only, and clinical features and T1DGRS models (without islet-autoantibodies) were also assessed for discrimination of T1D and T2D defined by the restrictive secondary definition of subtype.

To assess potential utility of the clinical features only model for guiding islet-autoantibody testing we assessed the relationship between the clinical features only model probability and positive islet-autoantibodies (≥1 positive) and assessed the proportion of participants with T1D defined by primary study outcome, split by islet-autoantibody status, across deciles of model probability.

##### Development of a points-based score

To aid clinical utility, models were converted to a simplified points-based scoring system (the StartRight Score) by categorising continuous variables and rounding the model beta coefficients.

##### Exploration of model utility in population data (Clinical Practice Research Datalink (CPRD))

To explore the potential implications of using the models to reduce missed T1D in a population setting, we assessed whether the clinical features only model probability was associated with features suggesting misclassified T1D in Clinical Practice Research Datalink (CPRD) Aurum,^24^ which includes general practice records and linked hospital episode statistics for over 1·6 million people living with diabetes. Due to data availability any diabetes parental history replaced parental non-insulin treated diabetes. In participants with incident diabetes age ≥18 initially treated as T2D (without insulin for ≥4 weeks) between 2018 and June 2024 we assessed the subsequent occurrence of insulin treatment within 3 years, ketoacidosis, a T1D diagnostic code, or basal-bolus insulin regime at latest follow-up across model probability and StartRight Score ranges. This analysis was repeated in subgroups of recorded ethnicity and in all those with diabetes, regardless of initial treatment. Further details of CPRD analysis are given in Appendix pp7-8. Full data processing and quality checks can be found at https://github.com/Exeter-Diabetes/CPRD-Cohort-scripts/tree/main/01-At-diagnosis.

##### Clinical utility for prioritisation of islet-autoantibody testing

To further explore model utility to guide islet-autoantibody testing, we compared the illustrative performance of the clinical features only model (test ≥10% probability of T1D) and the StartRight Score (test ≥7 points; closest cut-off to 10% full model probability) against three additional islet-autoantibody testing strategies:

1. Universal islet-autoantibody testing
2. Islet-autoantibody testing where T1D is suspected according to criteria listed in UK NICE guidance:^1^ having one or more of: ketosis, unintentional weight-loss, age-at-diagnosis <50 years, BMI <25 kg/m^2^, personal, and/or family history of autoimmune disease, with ketoacidosis used in place of ketosis.
3. Islet-autoantibody testing where T1D is suspected according to criteria listed in joint American/European Diabetes Association (ADA/EASD) Guidance:^2^ age-at-diagnosis <35 years, BMI <25 kg/m^2^, unintentional weight-loss, ketoacidosis, presentation glucose >20 mmol/L. Clinical features and cut offs included in this guidance were informed by analysis of baseline visit data from the StartRight cohort.^25^

These strategies were applied to participants in CPRD with diabetes diagnosis ≥18 years (Appendix p7) to identify the proportion of individuals from the population eligible for islet-autoantibody testing; and illustrative sensitivity and specificity calculated in combined StartRight and StartRight Prime by assuming that those islet-autoantibody negative or not meeting criteria for islet-autoantibody testing were had predicted T2D, and those meeting criteria who were islet-autoantibody positive were had predicted T1D.

## Results

### Cohort characteristics

Flow charts showing included participants and reasons for exclusion from the analysis are shown in Appendix p5 (StartRight and StartRight Prime).

For the StartRight cohort (diagnosed 18–50 years inclusive), 881 participants were included for assessment of features associated with diabetes subtype, (T1D/T2D by study primary outcome definition 323/558). 833 participants with complete data for included features were included in model development. Participant characteristics are shown in Table 1 (all included participants), and Appendix p9 (model development cohort). Mean duration of diabetes at recruitment and ≥3-year follow up was 17·4 weeks [IQR: 8,33] and 4·0 years [IQR: 3·5,4·7] respectively.

**Table 1:** Characteristics of included participant (StartRight, age 18-50 cohort), combined and split by study definition of type 1 and type 2 diabetes defined by 3-year insulin-use and C-peptide. Numerical characteristics are described by median [Interquartile range], and categorical characteristics by n (percentage). *Recruitment BMI used where missing diagnosis weight (n=286). **Plasma or capillary C-peptide. Capillary C-peptide limit of detection for reported results 200pmol/L, lower/undetectable results imputed at 199pmol/L. ***Osmotic symptoms defined by self-report nocturia, polyuria, and/or increased thirst.

| Characteristic |  | All | Type 1 | Type 2 |
| --- | --- | --- | --- | --- |
| N |  | 881 | 323 | 558 |
| Sex | Female | 377 (42.8%) | 157 (48.6%) | 220 (39.4%) |
|  | Male | 504 (57.2%) | 166 (51.4%) | 338 (60.6%) |
| Ethnicity | White | 787 (89.3%) | 304 (94.1%) | 483 (86.6%) |
|  | South Asian | 43 (4.9%) | 8 (2.5%) | 35 (6.3%) |
|  | Black | 27 (3.1%) | 4 (1.2%) | 23 (4.1%) |
|  | Other | 10 (1.1%) | 4 (1.2%) | 6 (1.1%) |
|  | Mixed | 14 (1.6%) | 3 (0.9%) | 11 (2.0%) |
| Clinical diagnosis at study recruitment | Type 1 | 372 (42.2%) | 295 (91.3%) | 77 (13.8%) |
|  | Type 2 | 440 (49.9%) | 13 (4.0%) | 427 (76.5%) |
|  | Uncertain | 69 (7.8%) | 15 (4.6%) | 54 (9.7%) |
| Duration of diabetes at recruitment (weeks) |  | 17.4 [8.0;33.0] | 14.4 [6.5;30.6] | 18.4 [9.2;33.8] |
| Duration of diabetes at latest follow up (years) |  | 4.0 [3.5;4.7] | 3.9 [3.5;4.7] | 4.0 [3.5;4.7] |
| Age at diagnosis (years) |  | 40 [33;46] | 33 [26;40] | 44 [38;47] |
| BMI (kg/m<sup>2</sup>) at diagnosis* |  | 29.0 [24.1;35.3] | 23.5 [20.9;26.5] | 33 [28;38] |
| Waist-hip ratio |  | 0.93 [0.86;0.99] | 0.87 [0.82;0.93] | 0.96 [0.9;1.0] |
| HbA1c at diagnosis (mmol/mol) |  | 89 [60;111] | 104 [84;124] | 78 [54;103] |
| ≥3-year C-peptide (pmol/L) ** |  | 792 [199;1,620] | 199 [73;255] | 1,505 [1,000;2,191] |
| Parent history of non-insulin-treated diabetes |  | 230 (26.1%) | 40 (12.4%) | 190 (34.1%) |
| Presentation Ketoacidosis |  | 85 (9.7%) | 64 (19.8%) | 21 (3.9%) |
| Presentation unintentional weight-loss |  | 437 (49.6%) | 270 (83.6%) | 167 (29.9%) |
| Presentation osmotic symptoms*** |  | 718 (81.5%) | 306 (94.7%) | 412 (73.8%) |
| Presence of additional autoimmune disorder/s |  | 97 (11.0%) | 44 (13.6%) | 53 (9.5%) |
| Parent history of autoimmune disorder |  | 214 (24.3%) | 97 (30.0%) | 117 (21.0%) |
| Number of positive islet autoantibodies (of GAD/IA2/Znt8) | 0 | 524 (59.5%) | 48 (14.9%) | 476 (85.3%) |
|  | 1 | 158 (17.9%) | 97 (30.0%) | 61 (10.9%) |
|  | 2 | 88 (10.0%) | 75 (23.2%) | 13 (2.3%) |
|  | 3 | 110 (12.5%) | 102 (31.6%) | 8 (1.4%) |

For assessment of the performance of clinical features for diabetes classification in older adults (StartRight Prime, diagnosis age >50 years) 470 participants (T1D/T2D 109/361) were included in analysis (Appendix p10). Of these, 441participants had complete data for assessment of model performance (Appendix p11). Median duration at recruitment and ≥3-year follow up was 21·1 weeks and 4·1 [IQR: 3·7,4·6] years.

805 and 419 participants in StartRight and StartRight Prime met criteria for the secondary subtype definition (subtype defined by both islet-autoantibody and C-peptide testing; Appendix p6). In those who could be classified by both the primary and secondary subtype definition, concordance was very high: 97·4% and 99·3% in StartRight and StartRight Prime respectively.

### Unintentional weight-loss and high presentation glycaemia are the most discriminant features for type 1 diabetes after age-at-diagnosis and BMI-at-diagnosis

Of 14 routinely available clinical features assessed, 11 added significantly to age-at-diagnosis and BMI in differentiating T1D and T2D (Appendix p12). The distribution of these features in T1D and T2D and univariate classification performance in complete case analysis is shown in Figure 1 and Appendix p13. Note presentation glucose is not shown in complete case analysis due to missingness (see below). The most discriminative features in isolation are BMI-at-diagnosis, unintentional weight-loss, waist-hip ratio, age-at-diagnosis, and HbA1c-at-diagnosis (Figure 1; Appendix p13). No single feature robustly differentiated these conditions in isolation, with the highest overall accuracy for a cut-off defined by Youden’s index 79% (BMI <28kg/m^2^), and highest T1D PPV 75% (ketoacidosis) (Appendix p13).

**Figure 1:**
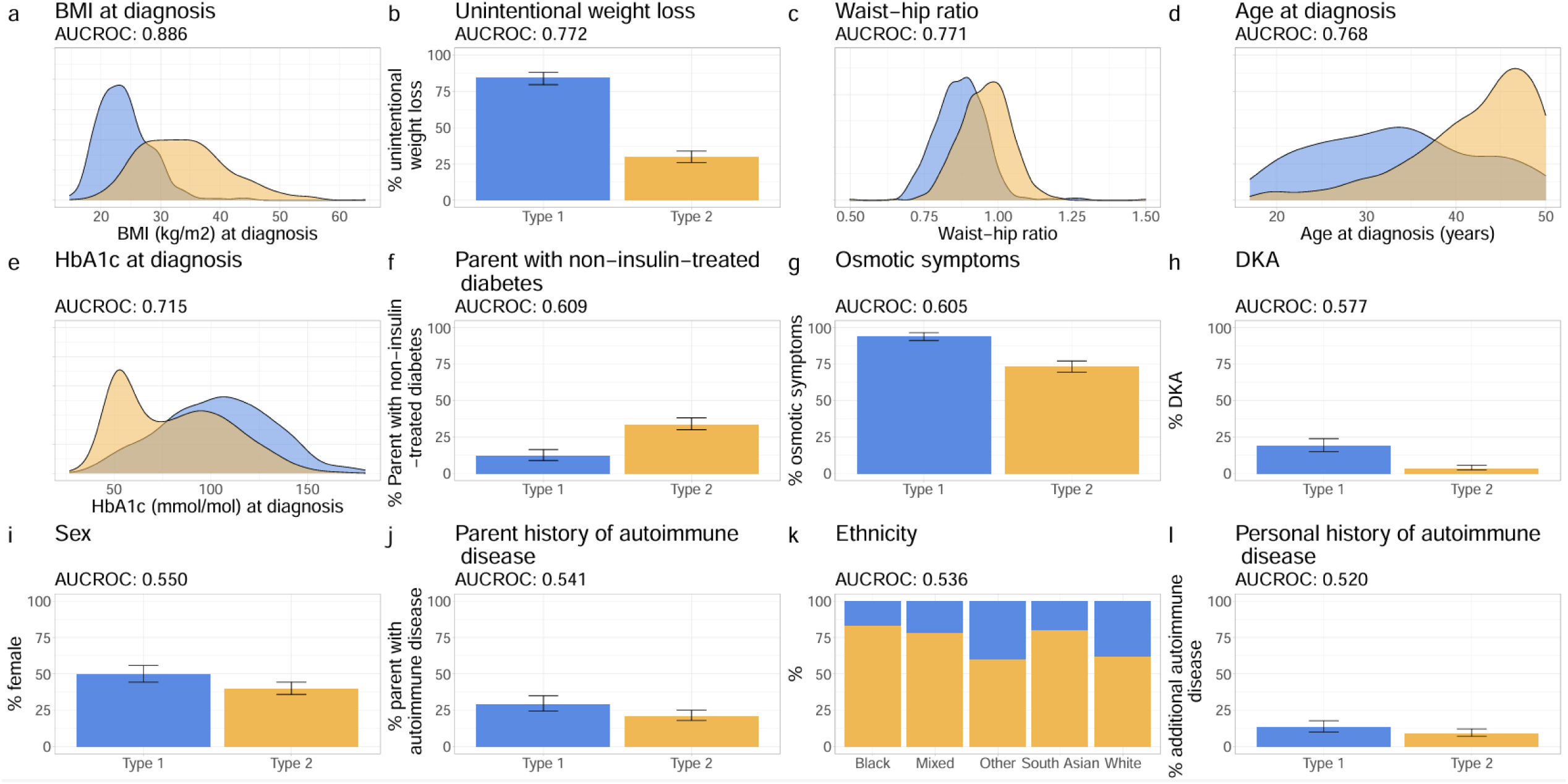
**Distribution of routine clinical features differentiating type 1 and 2 diabetes at diagnosis in participants aged 18-50 (StartRight)**, ranked by area under the Receiver Operating Characteristic (AUCROC). Complete case (n=809; type 1 = 292) univariate analysis of included features that add independently to age-and BMI-at-diagnosis (see Supplementary Table 5 for multivariable analysis). Diabetes subtype defined by ≥3-year C-peptide measurement and insulin treatment (see methods). Note presentation glucose (which has similar performance to presentation HbA1c) is not included due to high levels of missing data.

In 525 participants with both presentation glucose and HbA1c available, the performance of these features was similar (AUCROC 0·66 vs 0·66 respectively; Appendix p14). Due to high missingness of glucose-at-diagnosis only presentation HbA1c is shown in subsequent analysis.

In StartRight Prime (diagnosis age >50 years) the discriminative performance of these features was broadly similar (Appendix p15). Use of the restrictive subtype definition did not meaningfully change the association between clinical features and diabetes subtype, with near identical diagnostic performance in both cohorts (Appendix p16).

### Diagnostic models combining routine clinical features have high utility in discriminating between type 1 and 2 diabetes

A model combining age-at-diagnosis, BMI-at-diagnosis, HbA1c-at-diagnosis, sex, DKA, unintentional weight-loss, presence of autoimmune disorder, presence of osmotic symptoms, ethnicity (defined, due to sample size, as either black, south Asian, white, or other/mixed), and parent history of non-insulin-treated diabetes (clinical features only model) had high discriminative ability in StartRight, illustrating substantially greater performance than any single clinical feature alone (AUCROC=0·94 (95% CI: 0·93, 0·96), AUCPR=0·91, Figure 2, Appendix p17). The further inclusion of islet-autoantibodies (clinical features with islet-autoantibodies model) improved the model discrimination to AUCROC=0·97 (0·96, 0·98) (AUCPR=0·95). The addition of a previously reported T1DGRS did not meaningfully improve overall discrimination above islet-autoantibodies (AUCROC=0·97 (95% CI: 0·96, 0·98); AUCPR=0·95, Appendix pp17-18), but did improve the model performance when assessed by the likelihood ratio test and Akaike information criterion (AIC) (p<0·001; Appendix p19), and improved AUCROC above the clinical features only model (clinical features and T1DGRS AUCROC=0·96 (0·94, 0·97); p<0·001, Appendix p18). All models had good calibration in the development dataset (Figure 2, Appendix p18). Internal validation with 1,000 bootstraps showed low levels of optimism and good stability in model predictions (Appendix pp20-22). Model coefficients are shown in Appendix p23.

**Figure 2:**
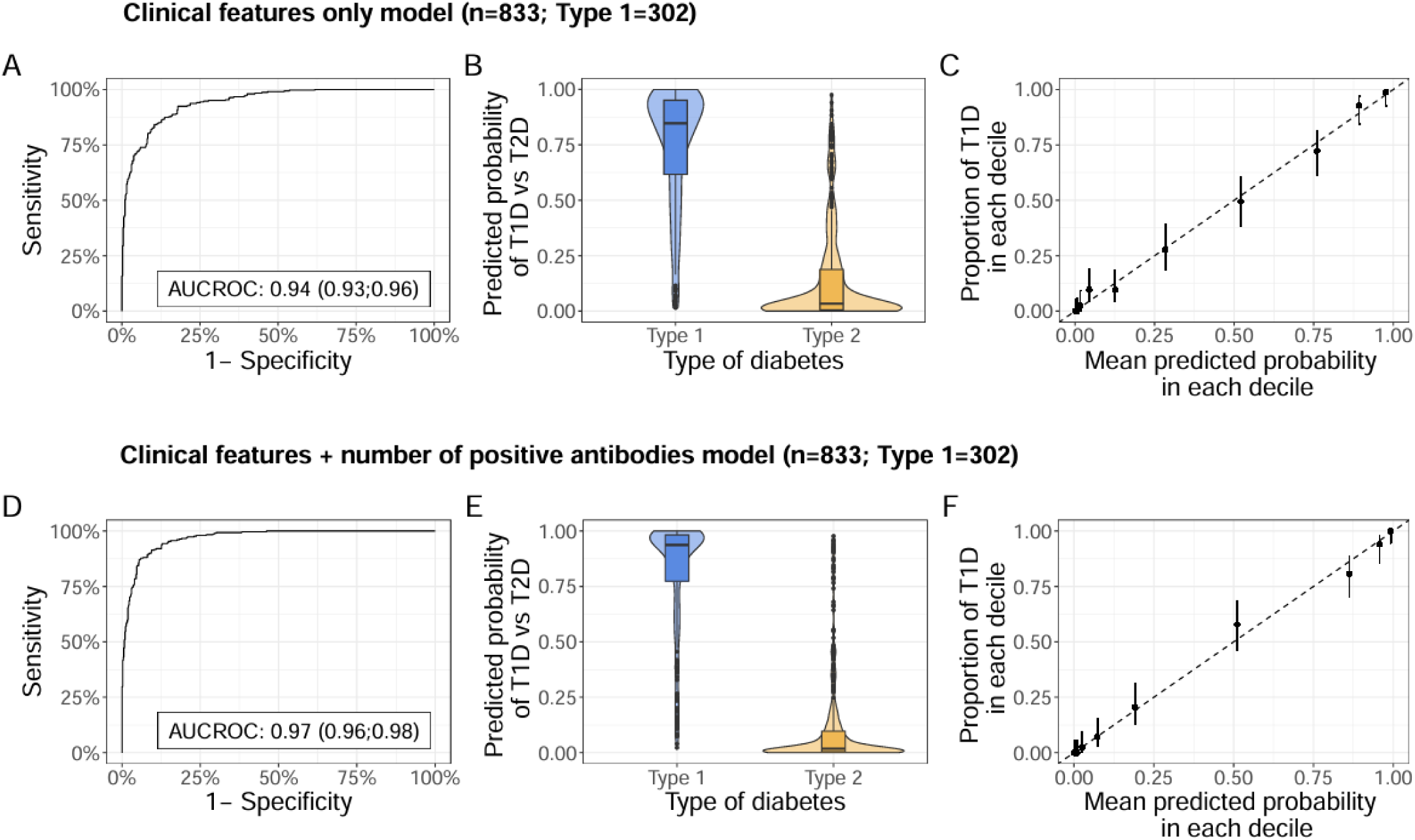
**Model separation and calibration in individuals aged 18-50 (StartRight, development cohort)**, for: clinical features only (age-at-diagnosis, BMI-at-diagnosis, HbA1c-at-diagnosis, sex, DKA, unintentional weight-loss, presence of an autoimmune disorder, presence of osmotic symptoms, and parent history of non-insulin-treated diabetes), and clinical features and islet-autoantibodies (clinical features and number of positive islet-autoantibodies of GAD, IA2 and ZNT8) models. Plots A and D denote the Receiver Operating Characteristic (ROC) Curve for each respective model, with the Area Under the Curve (AUCROC) shown on the plot. Plots B and E, represent the predicted probability of the respective model by primary study outcome. Plots C and F illustrate the calibration of each model, with deciles of model predicted probabilities of type 1 diabetes plotted against the observed proportion of type 1 diabetes defined by primary study outcome.

### Models maintain discrimination in older adults

All models in older adults (StartRight Prime, diagnosis age >50 years) showed similar discriminative performance to those diagnosed ≤50 years (Figure 3). Consistent with the enrichment for insulin treated individuals in StartRight Prime (and therefore over-representation of T1D in this age group), calibration plots demonstrated under-prediction of the probability of T1D in those with intermediate probabilities for the clinical features only model, however this was modest with the inclusion of islet-autoantibodies and/or T1DGRS (Figure 3 and Appendix p18).

**Figure 3:**
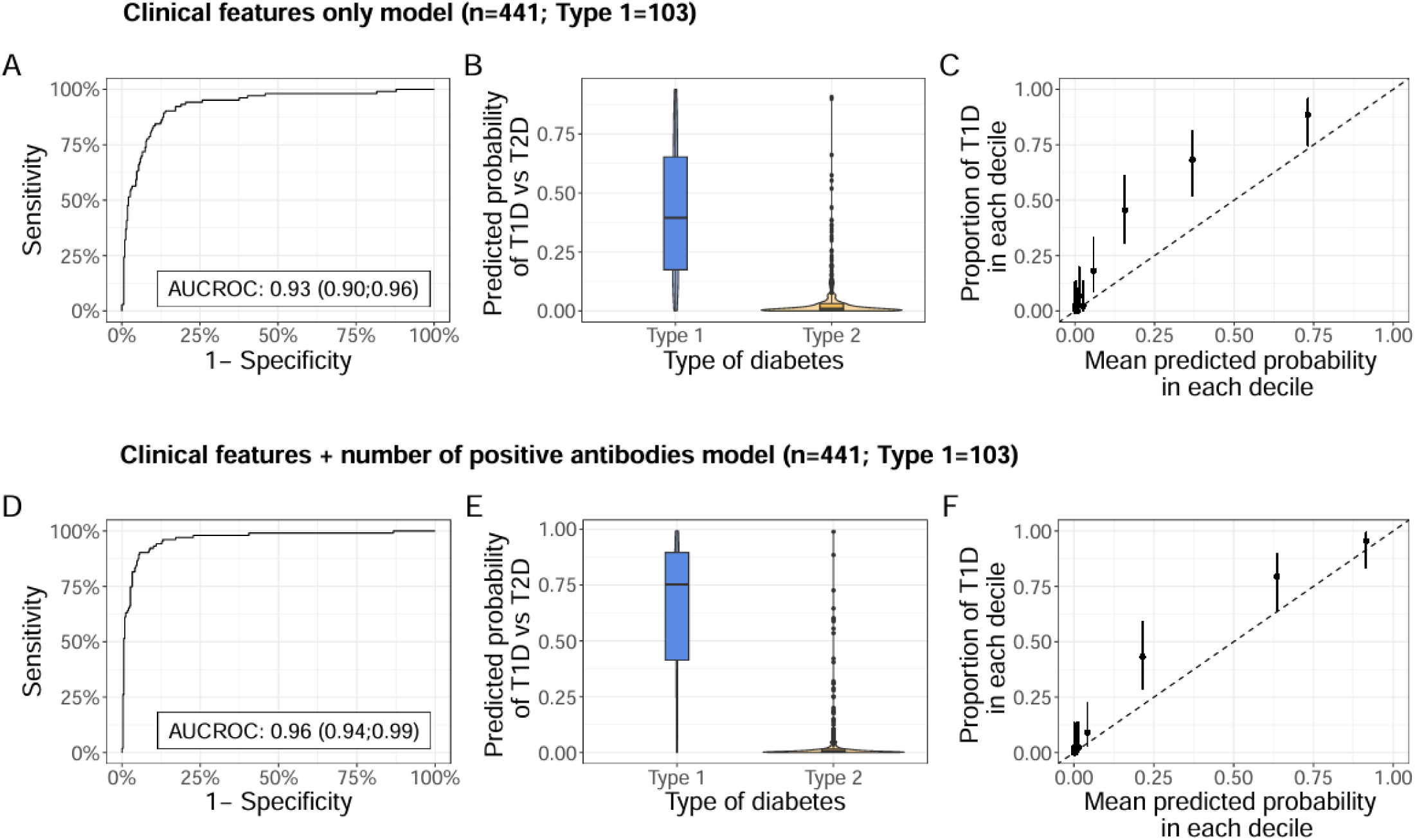
**Model separation and calibration in individuals aged >50 (StartRight Prime, performance assessment cohort)**, for: clinical features only (age-at-diagnosis, BMI-at-diagnosis, HbA1c-at-diagnosis, sex, DKA, unintentional weight-loss, presence of an autoimmune disorder, presence of osmotic symptoms, and parent history of non-insulin-treated diabetes), and clinical features and islet-autoantibodies’(clinical features and number of positive islet-autoantibodies of GAD, IA2 and ZNT8) models. Plots A and D denote the Receiver Operating Characteristic (ROC) Curve for each respective model, with the Area Under the Curve (AUCROC) shown on the plot. Plots B and E, represent the predicted probability of the respective model by primary study outcome. Plots C and F illustrate the calibration of each model, with deciles of model predicted probabilities of type 1 diabetes plotted against the observed proportion of type 1 diabetes defined by primary study outcome.

Use of the restrictive secondary definition of diabetes subtype did not meaningfully change the performance of the clinical features only and clinical features and T1DGRS models in either cohort (Appendix pp25-26).

### Combining features using a diagnostic model improves discrimination over clinical diagnosis or islet-autoantibodies alone

All models maintained high discrimination within those reporting T1D, T2D, or uncertain diabetes subtype at recruitment in both cohorts: StartRight AUCROC 0·84-0·94, StartRight Prime (excluding those reporting T2D due to low T1D frequency (n=5)) AUCROC 0·71-0·98 (Appendix p27-32). Models also maintained high discrimination in those with negative and positive islet-autoantibodies (AUCROC 0·88-0·94 and 0·88-0·91 respectively, Appendix p33-34). When applying a binary outcome (T1D if model probability of T1D ≥50%, T2D if model probability of T1D <50%) all models maintained substantially higher classification accuracy than islet-autoantibodies or T1DGRS alone (Appendix p35).

### Impact of participant ethnicity and sex

In those self-reporting black (n=25; T1D: n=4) and, separately, south Asian (n=40; T1D: n=8) ethnicity in StartRight, models maintained high discrimination (clinical features only model: AUCROC 0·86 (0·65, 1) and 0·95 (0·87, 1) respectively; clinical features with islet-autoantibodies model: AUCROC 0·95 (0·87, 1) and 0·97 (0·93, 1) respectively). While models appeared to maintain calibration, wide confidence intervals prevented robust assessment (Appendix pp36-37). Performance was also high by sex (Appendix pp38-41).

### The routine clinical features model may identify patients unlikely to benefit from islet autoantibody testing

The clinical features only model was strongly associated with islet-autoantibody positivity (StartRight/StartRight Prime: AUCROC 0·84/0.86; Appendix p42). In those with a low probability of T1D (<10%), participants were unlikely to be islet-autoantibody positive, and unlikely to have T1D as defined by the study outcome even in the presence of positive islet-autoantibodies (Appendix p43).

### StartRight Score: A simplified points-based score maintains similar model performance

A simplified scoring table, the StartRight Score, based on model coefficients from the clinical features only model, is shown in Table 2. The score shows high performance in adults diagnosed under and over 50 years: AUCROC=0·94 (0·92; 0·95) and 0·92 (0·89; 0·95) in StartRight and StartRight Prime respectively (Appendix p44). The relationship of the StartRight Score with islet-autoantibody status and outcomes are shown in Appendix pp45-46.

**Table 2:** Scoring table for use at diagnosis for the classification of type 1 diabetes versus type 2 diabetes. Scores derived from rounding beta coefficients of clinical features model to nearest integer. *note this scoring table does not consider other subtypes of diabetes associated with negative islet-autoantibodies, for example monogenic diabetes or secondary diabetes. These statements therefore assume that clinical features do not suggest an alternative diagnosis.

| **StartRight Score** | **Score** |
| --- | --- |
| Age at diagnosis (years): 18-24 | 3 |
| 25-34 years | 2 |
| 35-44 years | 1 |
| ≥45 years | 0 |
| BMI at diagnosis (kg/m<sup>2</sup>): <25 | 5 |
| 25-34 kg/m<sup>2</sup> | 3 |
| ≥35 kg/m<sup>2</sup> | 0 |
| HbA1c at diagnosis (mmol/mol): <58 | 0 |
| ≥58 mmol/mol | 1 |
| Female sex | 1 |
| Presentation osmotic symptoms | 1 |
| Presence of additional autoimmune disease | 1 |
| Presentation unintentional weight-loss | 1 |
| Presentation Ketoacidosis | 1 |
| No parent history of non-insulin-treated diabetes | 1 |
| Ethnicity associated with high type 2 diabetes risk | -1 |
|  | **15** |
| **StartRight Score categories** | **Interpretation** |
| 0-6 | Consistent with type 2 diabetes* (mean probability of type 1 diabetes = 3%). Islet-autoantibody testing unlikely to be indicated in the absence of rapid glycaemic progression. |
| 7-11 | Uncertain subtype, consider islet-autoantibody testing. In those islet-autoantibody negative, the majority of this group (c.80%) will have type 2 diabetes* but type 1 diabetes is not excluded (c.20%)*. Single positive islet-autoantibodies will usually confirm type 1 diabetes (probability >70%), multiple positive islet-autoantibodies confirm type 1 diabetes. |
| 12-15 | Likely type 1 diabetes. High probability of type 1 diabetes regardless of islet-autoantibody result*. |

An online version of the StartRight Score can be found at https://julieanneknupp.shinyapps.io/StartRightScore/. A scoring table incorporating islet-autoantibodies is shown in Appendix p47, which maintained similar performance to the clinical features and islet-autoantibodies model in StartRight and StartRight Prime cohorts (Appendix p48). An online prototype calculator using the full models can be found at https://julieanneknupp.shinyapps.io/T1D_v_T2D_at_diagnosis/).

### In UK population data, the clinical features only model identifies individuals initially treated as type 2 diabetes with outcomes associated with misclassified type 1 diabetes across diverse ethnicities

In data from adult UK primary care records (CPRD) high likelihood of T1D (based on the clinical features only model, and separately, the StartRight Score) was very strongly associated with outcomes suggestive of T1D (insulin within 3 years of diagnosis, post-diagnosis ketoacidosis, subsequent T1D code, and basal-bolus insulin treatment at latest follow-up), across all participants and in those initially treated as T2D (Figure 4, Appendix pp49-53). There was a strong relationship between model probability and T1D associated outcomes across ethnicities (Appendix pp54-61).

**Figure 4:**
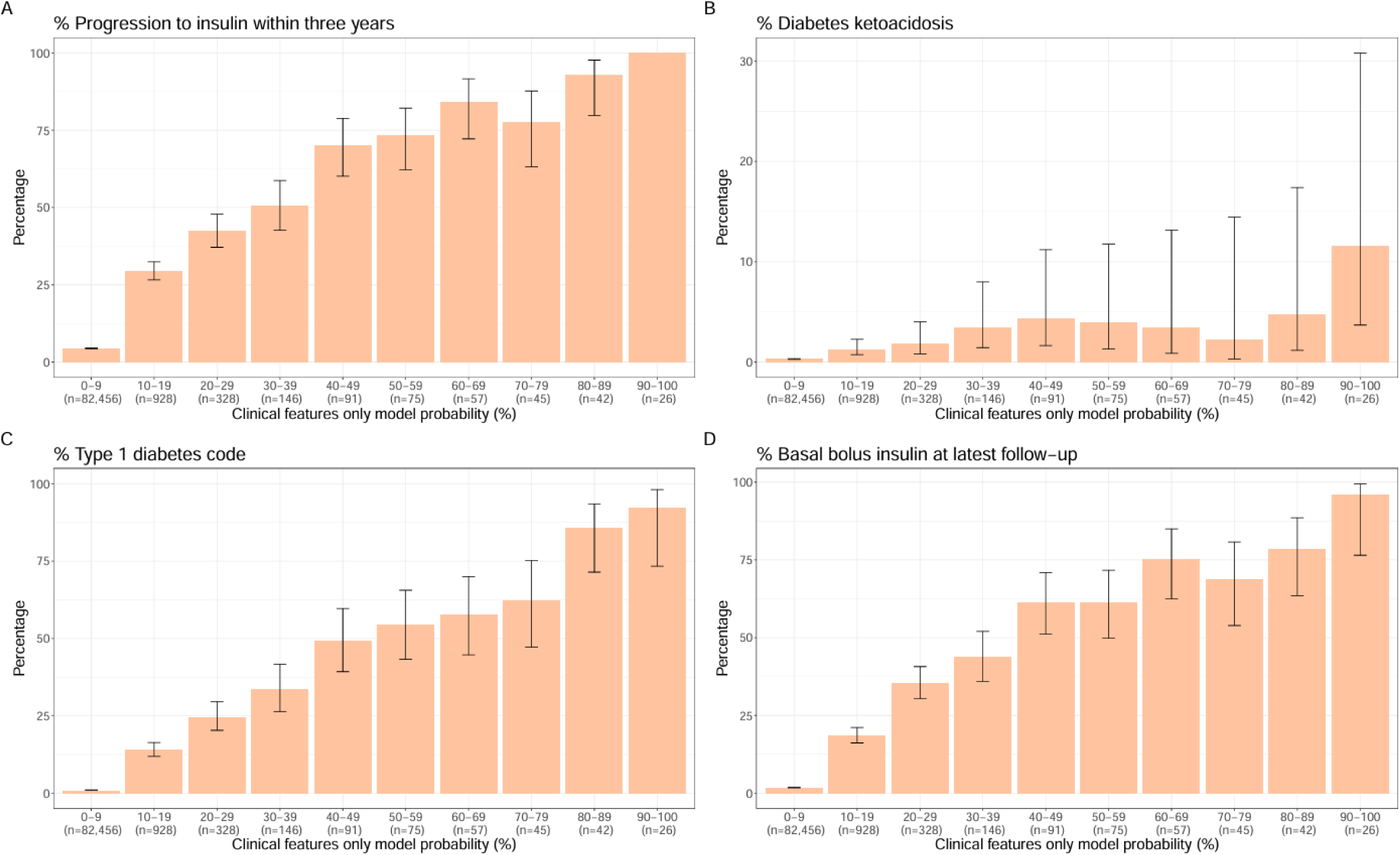
Distribution of early progression to insulin (A), subsequent ketoacidosis (B), a subsequent diagnostic code for type 1 diabetes (C) and subsequent basal bolus insulin (D) by clinical features only model probability in adults newly diagnosed with diabetes, initially treated without insulin (for ≥4 weeks), from UK primary care records (CPRD; n=84,692). Median (IQR) duration of follow up from diabetes diagnosis 4·6 (3·7, 5·4) years. Error bars denote logit-transformed 95% confidence intervals around the proportion. N’s below bars denote the total number of participants within that probability range (denominator of percentage).

### Utility of model and score for prioritisation of islet-autoantibody testing, in comparison to current guidelines

Figure 5 compares the proportion of all adults with incident diabetes in CPRD who would meet criteria for islet-autoantibody testing according to two major clinical guidelines and using a ≥10% clinical features only model cut off and a StartRight Score cut off of ≥7 (closest equivalent to 10% probability). Illustrative sensitivity and specificity for T1D for each strategy (assuming negative islet-autoantibodies or no testing indicated T2D, and positive islet-autoantibodies -where testing is indicated -T1D) is shown in the combined StartRight/StartRight Prime cohorts. Despite substantially less islet-autoantibody testing in population data, diagnostic performance was maintained in the combined StartRight cohorts (Appendix p62).

**Figure 5:**
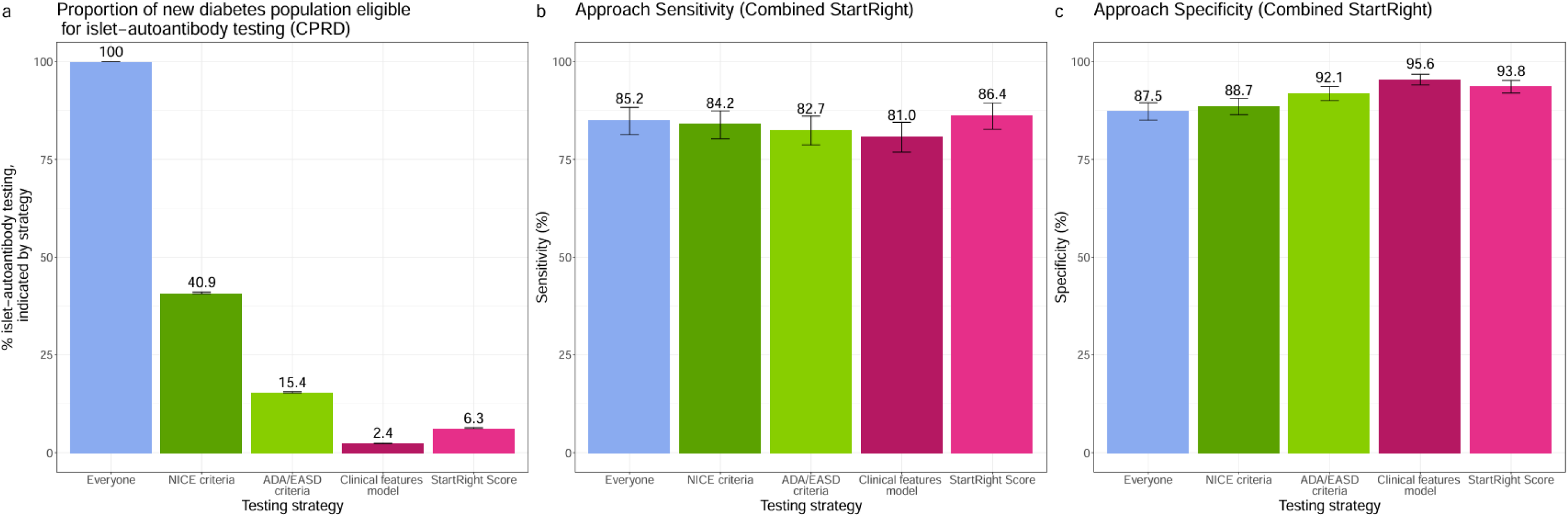
Performance of islet-autoantibody testing strategies in adults with incident diabetes. A) depicts the proportion of adults with incident diabetes in CPRD (UK population data; n=188,232) who meet the criteria for islet-autoantibody testing according to UK NICE guidance, American/European (ADA/EASD) joint guidance, using a ≥10% clinical features only model cut off, and a StartRight Score cut of ≥7 (closest equivalent to 10% on the clinical features only model). Panels B and C respectively illustrate the sensitivity and specificity for type 1 diabetes for each strategy (assuming negative islet-autoantibodies or no testing indicated type 2 diabetes, and positive islet-autoantibodies (where testing is indicated) type 1 diabetes) in the combined StartRight/StartRight Prime cohorts (n=1,274; type 1 = 405). Error bars denote logit-transformed 95% confidence intervals around the proportion. N’s above bars denote the proportion illustrated as a percentage.

## Discussion

This prospective observational study shows that alongside age-at-diagnosis and BMI (the only evidence based discriminative clinical features for T1D and T2D on previous systematic review) eleven additional features are discriminative. Unintentional weight-loss at diagnosis, waist-hip ratio, and presentation glycaemia (glucose or HbA1c) were highly discriminative, but many other features often included in current guidance, such as osmotic symptoms or family history of autoimmunity, were only very weak discriminators. No single clinical feature was able to adequately differentiate between T1D and T2D alone.

We found that combining routine clinical features, with or without islet-autoantibodies, in a prediction model was highly discriminative between T1D and T2D at diagnosis. Models developed in a cohort of adults aged 18-50 (inclusive), at diabetes diagnosis, maintained performance in older adults (diagnosis age >50 years) and strongly predicted future outcomes associates with T1D in UK population data, even in those initially treated as having T2D (without insulin).

Despite the high global prevalence of T1D and T2D there has been limited robust research on what features optimally distinguish these conditions. Systematic review in 2015 concluded that only age-at-diagnosis and BMI had robust evidence for discriminating T1D and T2D at diagnosis (with subsequent rapid insulin requirement also discriminative) and noted a remarkable lack of robust studies.^11^ 2022 systematic review by NICE in the UK^12^ also demonstrated a lack of robust evidence to inform guidance, with research on the use of clinical features to differentiate T1D and T2D, and their combination, highlighted as a priority for patient care.^1^ Previous research has also largely used cross-sectional data and assessed limited clinical features: the availability of extensive features and biomarkers at diabetes diagnosis, and robust classification, based on long-term C-peptide, are key strengths of this research.

To our knowledge, this is the first study to develop a calculator for distinguishing between T1D and T2D at diagnosis in adults. Calculators for classification of prevalent diabetes have been published from cross-sectional data of patients with longstanding diabetes, in European and Chinese populations, using a very limited available features.^26–29^ These calculators may have utility for targeting biomarker testing in prevalent cases to enable reclassification, and for subtyping in research datasets.^28^ We have also previously developed models to assist diagnosis of T1D from T2D in American youth (age ≤19). While these models showed high performance, availability of features at diabetes diagnosis were limited.^30^

Strengths of this research include that this is, to our knowledge, the first study of a mixed adult cohort of T1D and T2D from diagnosis with specific assessment of potentially discriminative features for classification, and/or follow up measurement of insulin secretion (C-peptide). This means that we have, for the first time, been able to robustly assess the performance of a range of routine clinical features, and the utility of islet-autoantibodies, at diabetes diagnosis, alone, and in classification models, against a robust independent definition of subtype based on future C-peptide measurement.^1,31^ We have utilised robust model building procedures and internal validation and assessed performance in an independent dataset, with additional analysis supportive of potential model utility in UK population data.

Limitations of this study include that our older cohort (>50 years) was enriched for T1D, as the high incidence of T2D in this age group make completing a population representative study with sufficient numbers of T1D cases unfeasible. The true prevalence of T1D in newly diagnosed older adults will be much lower, so we cannot assess true model calibration in older adults.^5,32^ Additional analysis was undertaken in the UK population-representative CPRD that supports potential utility of the clinical features only model, however we could not assess models that included islet-autoantibodies or genetic risk in this cohort, as these features were not available. Furthermore, CPRD analysis may underestimate performance as many model features are likely to be poorly recorded (such as symptoms or family history) and we were only able to assess outcomes that imperfectly capture subtype.^33,34^ An additional limitation is that non-routine biomarkers were measured up to 12 months after diagnosis. While this does not meaningly affect interpretation of the islet-autoantibodies (which are stable across this time period in adults,^35^ we are unable to assess the utility of C-peptide and lipids for classification at diagnosis, as these measures may be temporarily altered in the setting of severe hyperglycaemia common at diagnosis, and C-peptide may rapidly decline in the year after diagnosis.^36,37^ While we included ethnicity in our models this was based on small sample sizes and further work to assess classification models in different ethnicities and populations is a key priority. While this and previous work suggests discrimination will be similar by ethnicity,^30,38^ differences in T1D and T2D prevalence by ethnicity and country means validation for the intended population is critical for clinical use.^39,40^

This study provides robust evidence for clinicians and guidelines writers on which clinical features distinguish T1D and T2D at diagnosis, addressing an important evidence gap. Importantly we show that no individual clinical feature is diagnostic of T1D or T2D in isolation, and that many features included in current guidelines as features that should lead to consideration of T1D are only very weakly discriminative, meaning these guidelines may lead to high rates of islet-autoantibody testing in those at low risk, and potential harm from false positive results.^7^ The models and score we have developed offer a simple evidence-based approach for clinicians to assess evidence of T1D at diagnosis using routinely (and therefore freely) available features, to guide initial management, including need for islet-autoantibody measurement. With our combined models, islet-autoantibody (and/or T1DGRS) results can be interpreted considering other discriminative features, with substantially higher accuracy than islet-autoantibody testing alone.

In conclusion, after lower age-at-diagnosis and BMI, unintentional weight-loss, lower waist-hip ratio, and high presentation glycaemia are the most discriminative features for diagnosis of T1D in adults. Models combining routine clinical features, with or without islet-autoantibodies, have high accuracy and could assist clinical classification and prioritisation of classification biomarker testing.

## Data Availability

Data are available from the corresponding author on reasonable request. The UK routine clinical data analysed during the current study are available in the CPRD repository (CPRD; https://cprd.com/research-applications), but restrictions apply to the availability of these data, which were used under license for the current study, and so are not publicly available. For re-using these data, an application must be made directly to CPRD.

## Acknowledgements

StartRight study was funded by the UK National Institute of Health and Social Care Research (Clinician Scientist Fellowship CS-2015-15-018) and Diabetes UK (17/0005624). Genotyping for generation of the type 1 genetic risk score was supported by the European Foundation for the Study of Diabetes (2016 Rising Star Fellowship). Data analysis was additionally supported by Diabetes UK (reference 21/0006328). Data collection and analysis was supported by the NIHR Exeter Clinical Research Facility and NIHR Exeter Biomedical Research Centre (BRC). This article is based partly on data from the CPRD obtained under licence from the UK Medicines and Healthcare products Regulatory Agency. CPRD data are provided by patients and collected by the NHS as part of their care and support. The views expressed are those of the authors and not necessarily those of the NHS, NIHR or the Department of Health and Social Care. We thank all study participants.

## Contributors

AH, ATH, BMS and AGJ designed the study. JK, AH, NJMT, TJMcDonald, BMS and AGJ researched the data. JK analysed the data and drafted the manuscript with assistance from BMS, TJMcKinley and AGJ. DPF derived the type 1 diabetes genetic risk score. KGY prepared CPRD data for analysis. All authors critically revised the manuscript and approved the final version. AGJ is the guarantor of this work and, as such, had full access to all the data in the study and takes responsibility for the integrity of the data and the accuracy of the data analysis. The corresponding author attests that all listed authors meet authorship criteria and that no others meeting the criteria have been omitted.

## Data Sharing

Data are available from the corresponding author on reasonable request.

The UK routine clinical data analysed during the current study are available in the CPRD repository (CPRD; https://cprd.com/research-applications), but restrictions apply to the availability of these data, which were used under license for the current study, and so are not publicly available. For re-using these data, an application must be made directly to CPRD.

