## Appendix for "Using routine clinical features to classify adult-onset diabetes at diagnosis: the StartRight prospective observational study"

### **Appendix Figure 1:**

Table of contents of TRIPOD+AI Reporting Guidelines and corresponding page numbers.

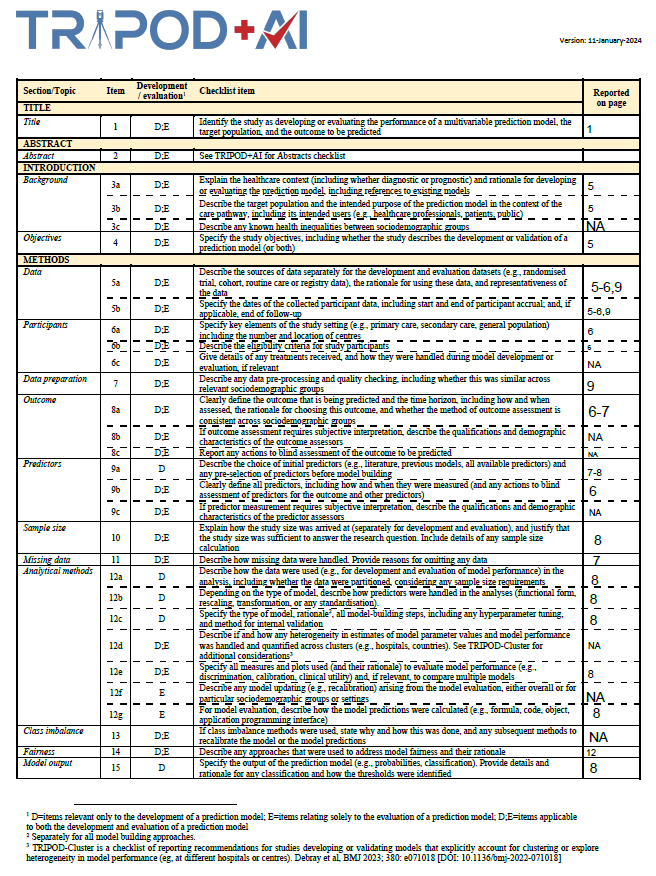

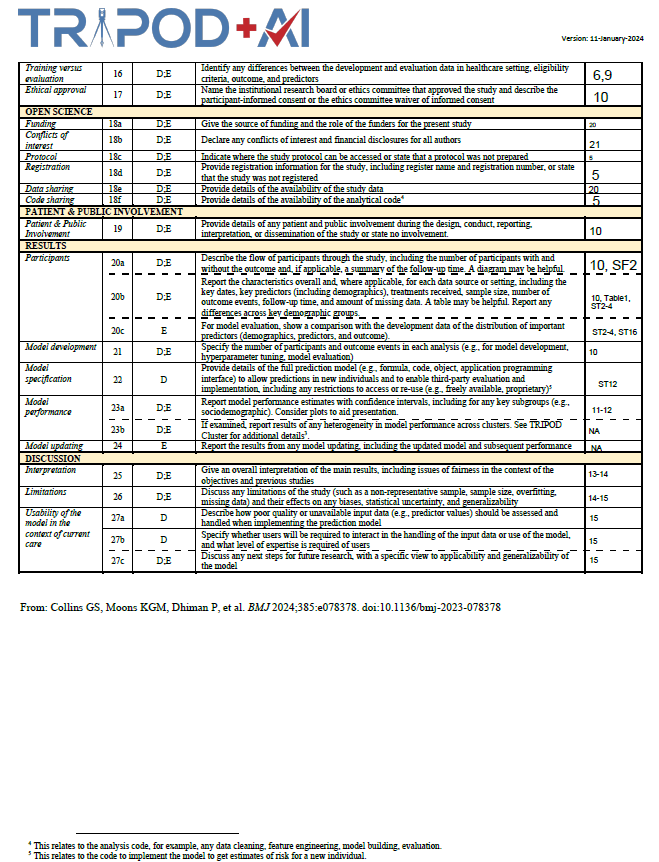

### **Appendix Figure 2:**

Inclusion and exclusion criteria in StartRight (adults diagnosed between 18 and 50 years inclusive) for primary outcome (diabetes type defined by three-year insulin-use and C-peptide).

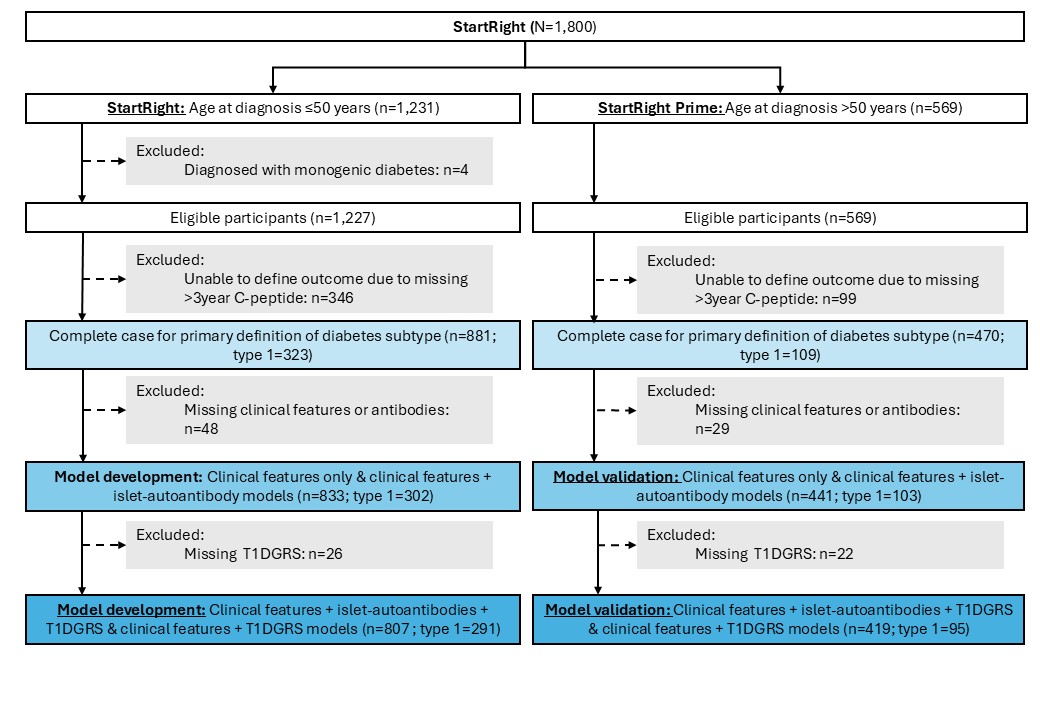

### **Appendix Figure 3:**

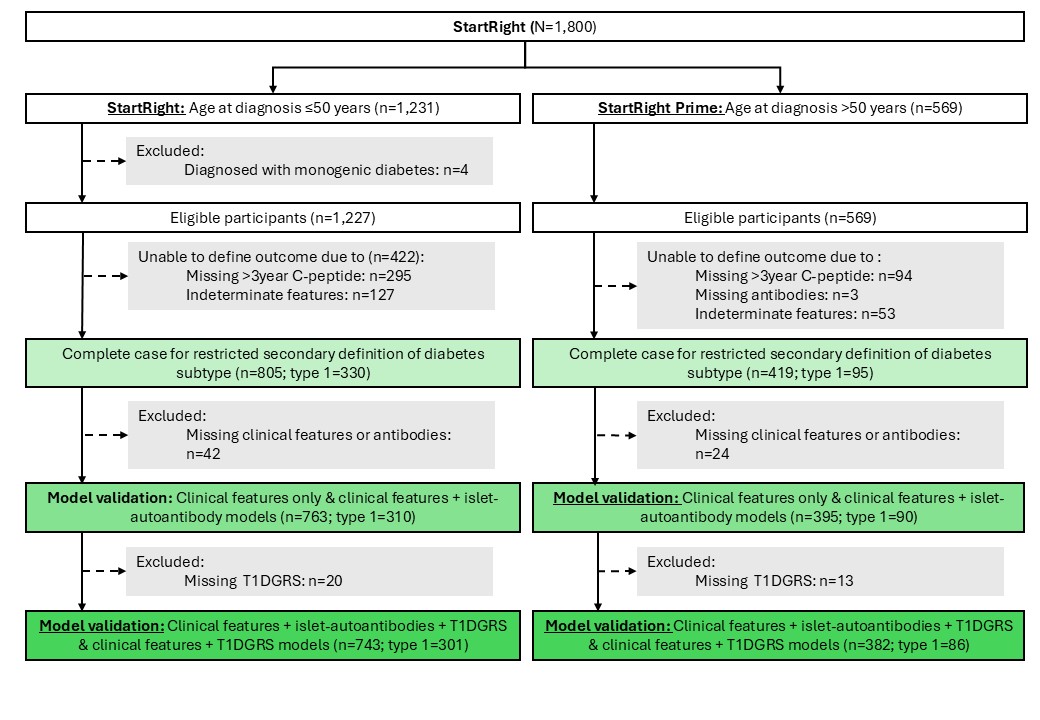
Inclusion and exclusion criteria in StartRight (adults diagnosed between 18 and 50 years) for restricted secondary outcome (diabetes type defined by three-year insulin-use, C-peptide and islet-autoantibodies).

### **Appendix Figure 4:**

CPRD cohort formation and study inclusion criteria (Hopkins et al., 2025). *Model input variables displayed in Appendix Table 1

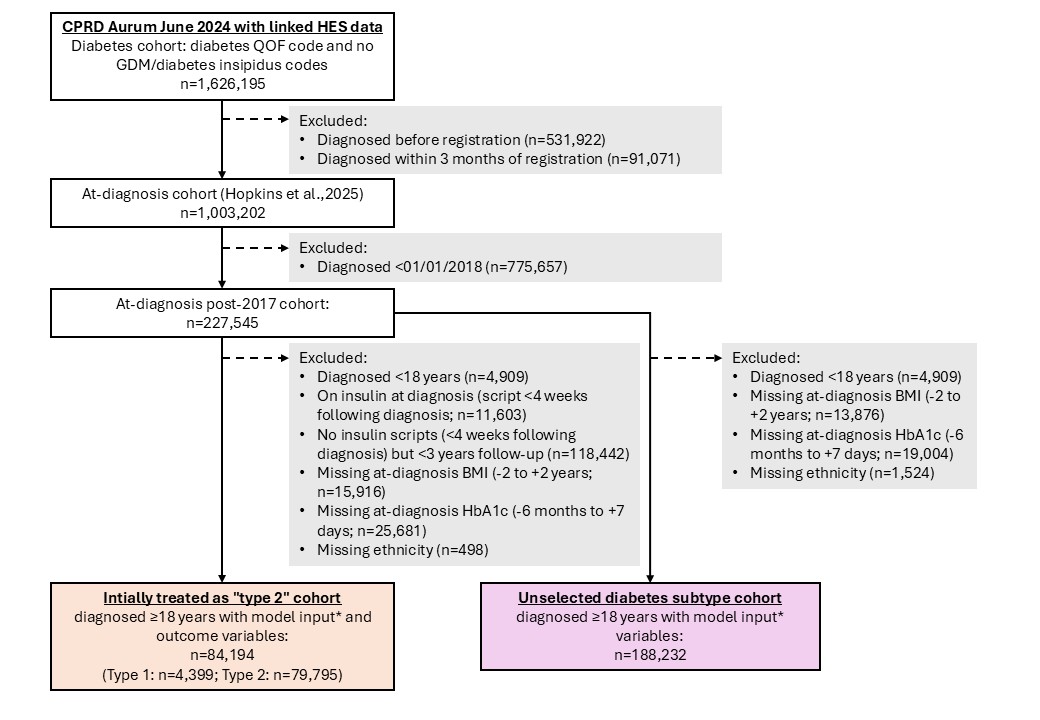

### Appendix Table 1:

Definition of variables in Clinical Practice Datalink (CPRD)) with accompanying hyperlinks to GitHub sources and codelists

| **Variables** | **CPRD definition** |
| --- | --- |
| **Model input variables** | |
| Diabetes diagnosis date | Earliest of [diabetes code](https://github.com/Exeter-Diabetes/CPRD-Codelists/blob/main/Medcodes/Diabetes/exeter_medcodelist_all_diabetes.txt), [code for insulin](https://github.com/Exeter-Diabetes/CPRD-Codelists/blob/main/Prodcodes/Diabetes%20medications/exeter_prodcodelist_insulin.txt)/[other diabetes medications](https://github.com/Exeter-Diabetes/CPRD-Codelists/blob/main/Prodcodes/Diabetes%20medications/exeter_prodcodelist_ohas.txt), [HbA1c](https://github.com/Exeter-Diabetes/CPRD-Codelists/blob/main/Medcodes/Biomarkers/exeter_medcodelist_hba1c.txt) ≥48 mmol/mol in GP records (excluding any codes before DOB/after deregistration/death) |
| BMI at diagnosis | Closest [BMI](https://github.com/Exeter-Diabetes/CPRD-Codelists/blob/main/Medcodes/Biomarkers/exeter_medcodelist_bmi.txt) to diagnosis in window of up to 2 years prior and up to 2 years after diagnosis. (All biomarkers are cleaned by removing low/high values and finding mean of values recorded on the same day as per here: [https://github.com/Exeter-Diabetes/CPRD-Codelists?tab=readme-ov-file#biomarker-algorithms](https://github.com/Exeter-Diabetes/CPRD-Codelists?tab=readme-ov-file)) |
| HbA1c at diagnosis | Closest [HbA1c](https://github.com/Exeter-Diabetes/CPRD-Codelists/blob/main/Medcodes/Biomarkers/exeter_medcodelist_hba1c.txt) to diagnosis in window of up to 6 months prior and up to 7 days after diagnosis. (Cleaned as above.) |
| Parent history of diabetes | [Code for positive or negative diabetes family history](https://github.com/Exeter-Diabetes/CPRD-Codelists/blob/main/Medcodes/Family%20history/exeter_medcodelist_fh_diabetes.txt) in mother/father/parent/unspecified relative/‘family history’ recorded any time (excluding sister/brother/child), excluding if specified as gestational/type 1. Set to 0/No if missing. |
| DKA at diagnosis | Hospital admission with [DKA](https://github.com/Exeter-Diabetes/CPRD-Codelists/blob/main/ICD10/exeter_icd10_dka.txt) as the primary cause within 30 days before or after diagnosis date. |
| Unintentional weight loss | [Code for weight loss](https://github.com/Exeter-Diabetes/CPRD-Codelists/blob/main/Medcodes/Comorbidities/exeter_medcodelist_weight_loss.txt) in GP records within 30 days before or after diagnosis date. Set to 0/No if not recorded. |
| Presence of osmotic symptoms | Code for [polydipsia](https://github.com/Exeter-Diabetes/CPRD-Codelists/blob/main/Medcodes/Comorbidities/exeter_medcodelist_polydipsia.txt) or [polyuria](https://github.com/Exeter-Diabetes/CPRD-Codelists/blob/main/Medcodes/Comorbidities/exeter_medcodelist_urinary_frequency.txt) in GP records within 30 days before or after diagnosis date. Set to 0/No if not recorded. |
| Presence of other autoimmune disorders | Code for coeliac, Grave’s disease, Hashimoto’s disease, Addison’s disease, vitiligo, pernicious anaemia or rheumatoid arthritis in GP records before or on same day as diabetes diagnosis date (excluding codes before DOB) –[codelist files](https://github.com/Exeter-Diabetes/T1DvsT2D_atDiagnosis_adults/tree/main/CPRD) |
| Ethnicity | [5-category ethnicity](https://github.com/Exeter-Diabetes/CPRD-Codelists#ethnicity) from primary care recorded ethnicity used preferentially and supplemented with HES recorded ethnicity where missing as recommended in https://pmc.ncbi.nlm.nih.gov/articles/PMC4245896/. (0=White, 1=South Asian, 2=Black, 3=Other, 4=Mixed, 5=Unknown) |
| **Islet-autoantibody testing strategy variables** | |
| Random glucose | Closest [random glucose](https://eur03.safelinks.protection.outlook.com/?url=https%3A%2F%2Fgithub.com%2FExeter-Diabetes%2FT1DvsT2D_atDiagnosis_adults%2Fblob%2Fmain%2FCPRD%2Frandom_glucose.txt&data=05%7C02%7CJ.Knupp%40exeter.ac.uk%7C4e9b56fa9f02448f3b5b08de64c86814%7C912a5d77fb984eeeaf321334d8f04a53%7C0%7C0%7C639059009052857447%7CUnknown%7CTWFpbGZsb3d8eyJFbXB0eU1hcGkiOnRydWUsIlYiOiIwLjAuMDAwMCIsIlAiOiJXaW4zMiIsIkFOIjoiTWFpbCIsIldUIjoyfQ%3D%3D%7C0%7C%7C%7C&sdata=qE8CJUiWVjYoOoLORNJu1f31pdkQOI4voBjGTu2wElE%3D&reserved=0) to diagnosis in window of up to 6 months prior and up to 7 days after diagnosis. (Cleaned as above.) |
| **Outcomes** |  |
| Insulin treatment in three years | Earliest [code for insulin](https://eur03.safelinks.protection.outlook.com/?url=https%3A%2F%2Fgithub.com%2FExeter-Diabetes%2FCPRD-Codelists%2Fblob%2Fmain%2FProdcodes%2FDiabetes%2520medications%2Fexeter_prodcodelist_insulin.txt&data=05%7C02%7CJ.Knupp%40exeter.ac.uk%7C4e9b56fa9f02448f3b5b08de64c86814%7C912a5d77fb984eeeaf321334d8f04a53%7C0%7C0%7C639059009052901683%7CUnknown%7CTWFpbGZsb3d8eyJFbXB0eU1hcGkiOnRydWUsIlYiOiIwLjAuMDAwMCIsIlAiOiJXaW4zMiIsIkFOIjoiTWFpbCIsIldUIjoyfQ%3D%3D%7C0%7C%7C%7C&sdata=J56Gd3soNo9s9cqWU0CfUTa5SGXiHap9YCfe7H8dBiE%3D&reserved=0) within the 3 years following diabetes diagnosis. |
| Any post-diagnosis Ketoacidosis | Any hospital admission with [DKA](https://eur03.safelinks.protection.outlook.com/?url=https%3A%2F%2Fgithub.com%2FExeter-Diabetes%2FCPRD-Codelists%2Fblob%2Fmain%2FICD10%2Fexeter_icd10_dka.txt&data=05%7C02%7CJ.Knupp%40exeter.ac.uk%7C4e9b56fa9f02448f3b5b08de64c86814%7C912a5d77fb984eeeaf321334d8f04a53%7C0%7C0%7C639059009052922387%7CUnknown%7CTWFpbGZsb3d8eyJFbXB0eU1hcGkiOnRydWUsIlYiOiIwLjAuMDAwMCIsIlAiOiJXaW4zMiIsIkFOIjoiTWFpbCIsIldUIjoyfQ%3D%3D%7C0%7C%7C%7C&sdata=xeT37xaJMty0ACTSVW6UWpJHZS1cNUKxTcjtJDulqpM%3D&reserved=0) as the primary cause more than 30 days after date of diagnosis. |
| Any Type 1 diagnosis code | Any code for Type 1 diabetes (subset of [diabetes codes](https://eur03.safelinks.protection.outlook.com/?url=https%3A%2F%2Fgithub.com%2FExeter-Diabetes%2FCPRD-Codelists%2Fblob%2Fmain%2FMedcodes%2FDiabetes%2Fexeter_medcodelist_all_diabetes.txt&data=05%7C02%7CJ.Knupp%40exeter.ac.uk%7C4e9b56fa9f02448f3b5b08de64c86814%7C912a5d77fb984eeeaf321334d8f04a53%7C0%7C0%7C639059009052943564%7CUnknown%7CTWFpbGZsb3d8eyJFbXB0eU1hcGkiOnRydWUsIlYiOiIwLjAuMDAwMCIsIlAiOiJXaW4zMiIsIkFOIjoiTWFpbCIsIldUIjoyfQ%3D%3D%7C0%7C%7C%7C&sdata=9GPjnA277bERvPbpswYIzXL2a2dWSOmnNwoIbkemNhQ%3D&reserved=0) where category="type 1") |
| Basal bolus insulin treatment at latest follow-up | Any prescription for bolus insulin (subset of [insulin codes](https://eur03.safelinks.protection.outlook.com/?url=https%3A%2F%2Fgithub.com%2FExeter-Diabetes%2FCPRD-Codelists%2Fblob%2Fmain%2FProdcodes%2FDiabetes%2520medications%2Fexeter_prodcodelist_insulin.txt&data=05%7C02%7CJ.Knupp%40exeter.ac.uk%7C4e9b56fa9f02448f3b5b08de64c86814%7C912a5d77fb984eeeaf321334d8f04a53%7C0%7C0%7C639059009052965364%7CUnknown%7CTWFpbGZsb3d8eyJFbXB0eU1hcGkiOnRydWUsIlYiOiIwLjAuMDAwMCIsIlAiOiJXaW4zMiIsIkFOIjoiTWFpbCIsIldUIjoyfQ%3D%3D%7C0%7C%7C%7C&sdata=UcjeaboD1aZGF3XobxEtHfLIs83Px3PnC9UxF%2BQRt7M%3D&reserved=0) where drug_substance="Bolus insulin") within the last year of records (year before the earliest of: deregistration/data collection from GP practice/death) |

### Appendix Table 2:

**Characteristics of included participant (StatRight, age 18-50 cohort),** split by definition of type 1 and type 2 diabetes using three-year insulin-use and C-peptide. Numerical characteristics are described by median [Interquartile range], and categorical characteristics by n (percentage). *Recruitment used where missing diagnosis weight (n=262). **Osmotic symptoms defined by self-report nocturia, polyuria, and/or thirst.

| **Characteristic** |  | **All** | **Type 1** | **Type 2** |
| --- | --- | --- | --- | --- |
| N |  | 833 | 302 | 531 |
| Sex | Female | 361 (43·3%) | 150 (49·7%) | 211 (39·7%) |
|  | Male | 472 (56·7%) | 152 (50·3%) | 320 (60·3%) |
| Ethnicity | White | 744 (89·3%) | 283 (93·7%) | 461 (86·8%) |
|  | South Asian | 40 (4·8%) | 8 (2·7%) | 32 (6·0%) |
|  | Black | 25 (3%) | 4 (1·3%) | 21 (4·0%) |
|  | Mixed | 14 (1·7%) | 3 (1·0%) | 11 (2·1%) |
|  | Other | 10 (1·2%) | 4 (1·3%) | 6 (1·1%) |
| Clinical diagnosis at study recruitment | Type 1 | 349 (41·9%) | 276 (91·4%) | 73 (13·8%) |
|  | Type 2 | 417 (50·1%) | 11 (3·6%) | 406 (76·5%) |
|  | Uncertain | 67 (8·0%) | 15 (5·0%) | 52 (9·8%) |
| Duration of diabetes at recruitment (weeks) | | 17.6 [8.1,33.0] | 14·7 [6·6,31·4] | 18·6 [9·4,33·6] |
| Duration of diabetes at latest follow up (years) | | 4.0 [3.5;4.7] | 3·9 [3·5,4·7] | 4·0 [3·5,4·7] |
| Age at diagnosis (years) | | 41.0 [33.0;46.0] | 33·0 [26·0;40·0] | 44·0 [38·0;47·0] |
| BMI (kg/m^2^) at diagnosis* | | 29.0 [24.2;35.4] | 23·4 [21·0;26·6] | 33·0 [28·0;38·0] |
| Waist-hip ratio | | 0.92 [0.86;0.99] | 0·87 [0·82;0·93] | 0·96 [0·9;1·01] |
| HbA1c at diagnosis (mmol/mol) | | 89.0 [60.0;111.0] | 104·0 [84·0;124·0] | 79·0 [54·0;103·0] |
| ≥3-year C-peptide (pmol/L) | | 803.0 [199.0;636.0] | 199·0 [73·0;251·0] | 1,494·0 [1,004·0;2,190·0] |
| Parent history of non-insulin-treated diabetes | | 216 (25.9%) | 36 (11·9%) | 180 (33·9%) |
| Presentation Ketoacidosis | | 75 (9.0%) | 56 (18·5%) | 19 (3·6%) |
| Unintentional weight-loss | | 412 (49.5%) | 253 (83·8%) | 159 (29·9%) |
| Presence of osmotic symptoms** | | 675 (81.0%) | 285 (94·4%) | 390 (73·5%) |
| Presence of additional autoimmune disorder/s | | 91 (10.9%) | 42 (13·9%) | 49 (9·2%) |
| Parent history of autoimmune disorders | | 203 (24.4%) | 89 (29·5%) | 114 (21·5%) |
| Number of positive islet-autoantibodies (of GAD/IA2/ZNT8) | 0 | 499 (59·9%) | 45 (14·9%) | 454 (85·5%) |
|  | 1 | 149 (17·9%) | 90 (29·8%) | 59 (11·1%) |
|  | 2 | 83 (10·0%) | 72 (23·8%) | 11 (2·1%) |
|  | 3 | 102 (12·2%) | 95 (31·5%) | 7 (1·3%) |

### Appendix Table 3:

**Characteristics of individuals in StartRight Prime**, split by definition of type 1 and type 2 diabetes using three-year insulin-use and C-peptide. Numerical characteristics are described by median [Interquartile range], and categorical characteristics by n (percentage). *Recruitment used where missing diagnosis weight (n=186). **Osmotic symptoms defined by self-report nocturia, polyuria, and/or thirst.

| **Characteristic** |  | **All** | **Type 1** | **Type 2** |
| --- | --- | --- | --- | --- |
| N |  | 470 | 109 | 361 |
| Sex | Female | 190 (40·4%) | 64 (58·7%) | 126 (34·9%) |
|  | Male | 280 (59·6%) | 45 (41·3%) | 235 (65·1%) |
| Ethnicity | White | 450 (95·7%) | 108 (99·1%) | 342 (94·7%) |
|  | South Asian | 7 (1·5%) | 0 (0·0%) | 7 (1·9%) |
|  | Black | 5 (1·1%) | 1 (0·9%) | 4 (1·1%) |
|  | Other | 5 (1·1%) | 0 (0·0%) | 5 (1·4%) |
|  | Mixed | 3 (0·6%) | 0 (0·0%) | 3 (0·8%) |
| Clinical diagnosis at study recruitment | Type 1 | 135 (28·7%) | 96 (88·1%) | 39 (10·8%) |
|  | Type 2 | 294 (62·6%) | 5 (4·6%) | 289 (80·1%) |
|  | Uncertain | 41 (8·7%) | 8 (7·3%) | 33 (9·1%) |
| Duration of diabetes at recruitment (weeks) | | 21.1 [11.0;35.0] | 18·0 [7·9;35·1] | 22·1 [11·4;34·3] |
| Duration of diabetes at latest follow up (years) | | 4.1 [3.7;4.6] | 4·0 [3·6;4·6] | 4·2 [3·8;4·6] |
| Age at diagnosis (years) | | 61.0 [55.0;68.0] | 57·0 [54·0;63·0] | 62·0 [56·0;69·0] |
| BMI (kg/m^2^) at diagnosis* | | 28.0 [25.0;33.5] | 23·2 [21·0;26·8] | 30·0 [26·5;35·0] |
| Waist-hip ratio | | 0.95 [0.89;1.0] | 0·89 [0·83;0·95] | 0·96 [0·92;1·0] |
| HbA1c at diagnosis (mmol/mol) | | 77.0 [51.0;109.0] | 108·0 [88·8;123·0] | 59·0 [50·0;99·0] |
| ≥3-year C-peptide (pmol/L) | | 1,340.0 [501.0;2,118.0] | 199·0 [89·0;286·0] | 1,683·0 [1,178·0;2,452·0] |
| Parent history of non-insulin-treated diabetes | | 95 (20.2%) | 16 (14·7%) | 79 (21·9%) |
| Presentation Ketoacidosis | | 35 (7.5%) | 27 (24·8%) | 8 (2·2%) |
| Unintentional weight-loss | | 184 (39.2%) | 93 (85·3%) | 91 (25·2%) |
| Presence of osmotic symptoms** | | 320 (68.1%) | 100 (91·7%) | 220 (60·9%) |
| Presence of additional autoimmune disorder/s | | 93 (19.8%) | 32 (29·4%) | 61 (16·9%) |
| Parent history of autoimmune disorders | | 84 (17.9%) | 28 (25·7%) | 56 (15·5%) |
| Number of positive islet-autoantibodies (of GAD/IA2/ZNT8) | 0 | 343 (73·0%) | 18 (16·5%) | 325 (90·0%) |
|  | 1 | 58 (12·3%) | 32 (29·4%) | 26 (7·2%) |
|  | 2 | 22 (4·7%) | 20 (18·4%) | 2 (0·6%) |
|  | 3 | 44 (9·4%) | 39 (35·8%) | 5 (1·4%) |

### Appendix Table 4:

**Characteristics of individuals in StartRight Prime utilised for model evaluation**, split by definition of type 1 and type 2 diabetes using three-year insulin-use and C-peptide. Numerical characteristics are described by median [Interquartile range], and categorical characteristics by n (percentage). *Recruitment used where missing diagnosis weight (n=172). **Osmotic symptoms defined by self-report nocturia, polyuria, and/or thirst.

| **Characteristic** |  | **All** | **Type 1** | **Type 2** |
| --- | --- | --- | --- | --- |
| N |  | 441 | 103 | 338 |
| Sex | Female | 179 (40·6%) | 61 (59·2%) | 118 (34·9%) |
|  | Male | 262 (59·4%) | 42 (40·8%) | 220 (65·1%) |
| Ethnicity | White | 423 (95·9%) | 102 (99·0%) | 321 (95·0%) |
|  | South Asian | 6 (1·4%) | 0 (0·0%) | 6 (1·8%) |
|  | Black | 5 (1·1%) | 1 (1·0%) | 4 (1·2%) |
|  | Other | 4 (0·9%) | 0 (0·0%) | 4 (1·2%) |
|  | Mixed | 3 (0·7%) | 0 (0·0%) | 3 (0·9%) |
| Clinical diagnosis at study recruitment | Type 1 | 126 (28·6%) | 91 (88·4%) | 35 (10·4%) |
|  | Type 2 | 277 (62·8%) | 5 (4·9%) | 272 (80·5%) |
|  | Uncertain | 38 (8·6%) | 7 (6·8%) | 31 (9·2%) |
| Duration of diabetes at recruitment (weeks) | | 21.7 [11.3;35.1] | 18·6 [8·7;35·9] | 22·5 [11·6;35·0] |
| Duration of diabetes at latest follow up (years) | | 4.1 [3.7;4.6] | 4·0 [3·6;4·6] | 4·2 [3·8;4·6] |
| Age at diagnosis (years) | | 41.0 [33.0;46.0] | 61·0 [55·0;67·0] | 55·0 [53·0;62·0] |
| BMI (kg/m^2^) at diagnosis* | | 29.0 [24.2;35.4] | 28·7 [25·0;34·0] | 24·5 [22·3;26·7] |
| Waist-hip ratio | | 0.95 [0.89;1 .0] | 0·89 [0·83;0·95] | 0·96 [0·92;1·0] |
| HbA1c at diagnosis (mmol/mol) | | 89.0 [60.0;111.0] | 77·0 [51·0;109·0] | 101·0 [87·3;115·0] |
| ≥3-year C-peptide (pmol/L) | | 1,340.0 [484.0;2,100.0] | 199·0 [87·0;277·0] | 1,666·5 [1179·8;2,452·3] |
| Parent history of non-insulin-treated diabetes | | 93 (21.1%) | 15 (14·6%) | 78 (23·1%) |
| Presentation Ketoacidosis | | 32 (7.3%) | 25 (24·3%) | 7 (2·1%) |
| Unintentional weight-loss | | 178 (40.4%) | 90 (87·4%) | 88 (26·0%) |
| Presence of osmotic symptoms** | | 303 (68.7%) | 95 (92·2%) | 208 (61·5%) |
| Presence of additional autoimmune disorder/s | | 85 (19.3%) | 30 (29·1%) | 55 (16·3%) |
| Parent history of autoimmune disorder | | 82 (18.6%) | 27 (26·2%) | 55 (16·3%) |
| Number of positive islet-autoantibodies (of GAD/IA2/ZNT8) | 0 | 321 (72·8%) | 15 (14·6%) | 306 (90·5%) |
|  | 1 | 55 (12·5%) | 30 (29·1%) | 25 (7·4%) |
|  | 2 | 22 (5·0%) | 20 (19·4%) | 2 (0·6%) |
|  | 3 | 43 (9·8%) | 38 (36·9%) | 5 (1·5%) |

### Appendix Table 5:

Routine clinical features added univariately to a model with age-at-diagnosis and BMI covariates in StartRight, predicting type 1 defined by 3-year insulin-use & C-peptide. Age-at-diagnosis, BMI, and all continuous variables scaled, therefore beta coefficients are standardised. Recruitment BMI is used where missing diagnosis BMI (n=286). LR Test refers to likelihood ratio test p-value against age-at-diagnosis and BMI model in n=878 StartRight participants

| **Clinical features** | **n** | **Non-reference category** | **Beta Coefficient (95% CI)** | **Beta Coefficient**  **p-value** | **LR Test p-value** | **AUCROC (95% CI) (Including age-at-diagnosis and BMI)** |
| --- | --- | --- | --- | --- | --- | --- |
| **Demographic features** |  |  |  |  |  |  |
| Sex | 878 | Male | -0·6 (-1·0; -0·2) | 0·003 | 0·003 | 0·91 (0·89;0·93) |
| Ethnicity | 878 | White | REF | / | 0·002 | 0·91 (0·89; 0·93) |
|  |  | Black | -0·87 (-2·2;0·27) | 0·16 |  |  |
|  |  | Mixed | -1·9 (-3·6; -0·45) | 0·016 |  |  |
|  |  | Other | -0·69 (-2·8; 1·4) | 0·52 |  |  |
|  |  | South Asian | -1·4 (-2·4; -0·44) | 0·006 |  |  |
| Waist circumference (cm) | 859 |  | -0·38 (-0·83;0·10) | 0·089 | 0·088 | 0·91 (0·89;0·93) |
| Waist- Hip ratio | 854 |  | -0·46 (-0·71; -0·20) | <0·001 | <0·001 | 0·92 (0·90;0·94) |
| **Glycaemic features** |  |  |  |  |  |  |
| Glucose at diagnosis (mmol/L) | 543 |  | 0·31 (0·10;0·60) | 0·019 | 0·018 | 0·90 (0·88;0·93) |
| HbA1c at diagnosis (mmol/mol) | 836 |  | 0·43 (0·21;0·66) | <0·001 | <0·001 | 0·92 (0·90;0·94) |
| **Presentation features** |  |  |  |  |  |  |
| DKA | 878 | Yes | 1·3 (0·61;1·9) | <0·001 | <0·001 | 0·91 (0·89;0·93) |
| Osmotic symptoms at presentation | 875 | Yes | 1·7 (1·0;2·4) | <0·001 | <0·001 | 0·92 (0·90;0·94) |
| Unintentional weight-loss | 875 | Yes | 1·7 (1·3;2·2) | <0·001 | <0·001 | 0·93 (0·91;0·94) |
| Additional autoimmune disorder | 878 | Yes | 1·1 (0·48;1·7) | <0·001 | <0·001 | 0·91 (0·89;0·93) |
| **Family history** |  |  |  |  |  |  |
| Parent history of insulin-treated diabetes | 869 | Yes | 0·03 (-0·53;0·58) | 0·91 | 0·91 | 0·91 (0·89;0·93) |
| Parent history of non-insulin-treated diabetes | 878 | Yes | -1·1 (-1·6;-0·64) | <0·001 | <0·001 | 0·91 (0·90;0·93) |
| Parent history of autoimmune disorder | 827 | Yes | 0·83 (0·37;1·3) | <0·001 | <0·001 | 0·91 (0·89;0·93) |
| **Additional features** |  |  |  |  |  |  |
| Acanthosis Nigricans | 874 | Yes | -0·31 (-1·9;0·97) | 0·66 | 0·65 | 0·91 (0·89;0·93) |
| Hypertension (on medication) | 877 | Yes | 0·07 (-0·64;0·74) | 0·84 | 0·84 | 0·91 (0·89;0·93) |

### Appendix Table 6:

**Univariate performance of routine clinical features in differentiating type 1 (T1D) and 2 diabetes (T2D) at diagnosis in participants aged 18-50 (StartRight), ranked by area under the Receiver Operating Characteristic Area Under the Curve (AUCROC**). Features (with exception of age-at-diagnosis and BMI) shown add discriminative ability above age-at-diagnosis and BMI. Complete case analysis (n=809; type 1 = 292). ^†^Optimal threshold calculated by Youden’s Index. ^‡^Based on “optimal threshold”, predictive value for T2D based on absence of the condition stated in optimal threshold for T1D column. *BMI-at-diagnosis or (where missing weight-at-diagnosis, n=256) at recruitment visit. **Optimal threshold in individuals diagnosed 18-50 years. ***Compared to White ethnicity reference group

| **Routine clinical feature** | **AUCROC (95% CI)** | **Optimal Threshold for T1D^†^** | **Sensitivity for T1D (%; 95% CI)^‡^** | **Specificity for T1D (%; 95% CI)^‡^** | **Predictive value for T1D (%; 95% CI)^‡^** | **Predictive value for T2D (%; 95% CI)^‡^** | **Accuracy (%; 95% CI)^‡^** |
| --- | --- | --- | --- | --- | --- | --- | --- |
| BMI (kg/m^2^)* | 0·89 (0·86;0·91) | <28 (kg/m^2^)* | 81·8  (77; 85·9) | 76·8  (73; 80·2) | 66·6  (61·5; 71·3) | 88·2  (84·9; 90·9) | 78·6  (75·7; 81·3) |
| Unintentional weight-loss | 0·77 (0·74; 0·80) | Yes | 84·2  (79·6; 88·0) | 70·2  (66·1; 74·0) | 61·5  (56·6; 66·1) | 88·8  (85·3; 91·5) | 75·3  (72·2; 78·13) |
| Waist-hip ratio | 0·77 (0·74; 0·80) | <0·9 | 63·7  (58·0; 69·0) | 76·4  (72·5; 79·9) | 60·4  (54·8; 65·7) | 78·8  (75·0; 82·2) | 71·8  (68·6; 74·8) |
| Age at diagnosis (years) | 0·77 (0·73;0·80) | <37 years** | 65·4  (59·8; 70·6) | 80·5  (76·8; 83·7) | 65·4  (59·8; 70·6) | 80·5  (76·8; 83·7) | 75  (71·9; 77·9) |
| HbA1c at diagnosis (mmol/mol) | 0·72 (0·68;0·75) | >80 mmol/mol | 81·5  (76·6; 85·6) | 50·7  (46·4; 55) | 48·3  (43·9; 52·7) | 82·9  (78·4; 86·7) | 61·8  (58·4; 65·1) |
| Parent history of non-insulin-treated diabetes | 0·61 (0·58; 0·64) | No | 88·0  (83·8; 91·3) | 33·8  (29·9; 38·0) | 42·9  (39·0; 46·9) | 83·3  (77·7; 87·8) | 53·4  (50·0; 56·8) |
| Presence of osmotic symptoms | 0·61 (0·57; 0·64) | Yes | 94·5  (91·2; 96·6) | 26·5  (22·9; 30·5) | 42·1  (38·3; 45·9) | 89·5  (83·6; 93·5) | 51·1  (47·6; 54·5) |
| DKA | 0·58 (0·54; 0·61) | Yes | 18·8  (14·8; 23·7) | 96·5  (94·5; 97·8) | 75·3  (64·2; 83·8) | 67·8  (64·3; 71·1) | 68·5  (65·2; 71·6) |
| Sex | 0·55 (0·52; 0·58) | Female | 50·0  (44·3; 55·7) | 60·0  (55·7; 64·1) | 41·4  (36·3; 46·6) | 68·0  (63·6; 72·1) | 56·4  (52·9; 59·7) |
| Parent history of autoimmune disorders | 0·54 (0·51; 0·58) | Yes | 29·5  (24·5; 34·9) | 78·7  (75·0; 82·0) | 43·9  (37·1; 50·9) | 66·4  (62·6; 70·0) | 60·9  (57·5; 64·2) |
| Ethnicity (5-factor)*** | 0·54 (0·52;0·56) | / | / | / | / | / | / |
| Presence of other autoimmune disorder | 0·52 (0·49; 0·56) | Yes | 13·4  (9·9; 17·8) | 90·7  (87·9; 92·9) | 44·8  (34·7; 55·4) | 65·0  (61·4; 68·4) | 62·8  (59·4; 66·1) |

### Appendix Table 7:

HbA1c- and glucose-a- diagnosis demonstrate similar discriminatory ability when differentiating type 1 and type 2 diabetes and show no statistically significant difference to each other. Univariate model performance in StartRight (n=525), predicting diabetes subtype defined by primary study outcome (by three-year insulin-use & C-peptide).

| **Clinical features** | **Beta Coefficient (95% CI)** | **Beta Coefficient p-value** | **AUCROC (95% CI)** | **AIC** |
| --- | --- | --- | --- | --- |
| Glucose at diagnosis (mmol/L) | 0·48 (0·30; 0·68) | <0·001 | 0·66 (0·61; 0·71) | 698·42 |
| HbA1c at diagnosis (mmol/mol) | 0·59 (0·41; 0·79) | <0·001 | 0·66 (0·61; 0·70) | 684·27 |

### Appendix Table 8:

**Univariate performance of routine clinical features in differentiating type 1 (T1D) and 2 diabetes (T2D) at diagnosis in participants aged 18-50 (StartRight: n=809; type 1=292**) **& StartRight Prime (n=435; type 1=99), ranked by area under the Receiver Operating Characteristic Area Under the Curve (AUCROC) in StartRight.** Note age-at-diagnosis not shown due to restricted age range. ^†^Optimal threshold calculated by Youden’s Index. ^‡^Based on “optimal threshold”. *BMI-at-diagnosis or (where missing weight-at-diagnosis, StartRight: n=256; StartRight Prime: n=170) at recruitment visit. ** StartRight: Black ethnicity (n=24; type 1 = 4), Mixed ethnicity (n=14, type 1 = 3), Other ethnicity (n=10, type 1 = 4), South Asian ethnicity (n=40, type 1 = 8), White ethnicity (Reference group: n=721, type 1 = 273); StartRight Prime: Black ethnicity (n=5; type 1 = 1), Mixed ethnicity (n=3, type 1 = 0), Other ethnicity (n=5, type 1 = 0), South Asian ethnicity (n=6, type 1 = 0), White ethnicity (Reference group: n=416, type 1 = 98).

|  |  | **StartRight (n=809)** | | | **StartRight Prime (n=435)** | | |
| --- | --- | --- | --- | --- | --- | --- | --- |
| **Routine clinical features** | **Optimal Variable Threshold†** | **AUCROC (95% CI)** | **Sensitivity of T1D**^‡^ **(%; 95% CI)** | **Specificity of T1D**^‡^ **(%; 95% CI)** | **AUCROC (95% CI)** | **Sensitivity of T1D**^‡^ **(%; 95% CI)** | **Specificity of T1D**^‡^ **(%; 95% CI)** |
| BMI (kg/m^2^)* | <28 (kg/m^2^)* | 0·89 (0·86;0·91) | 81·8 (77·0; 85·9) | 76·8 (73·0; 80·2) | 0·81 (0·77;0·86) | 84·9 (76·4; 90·7) | 66·4 (61·2; 71·2) |
| Unintentional weight-loss | Yes | 0·77 (0·74; 0·80) | 84·2 (79·6; 88·0) | 70·2 (66·1; 74·0) | 0·81 (0·77; 0·84) | 87·9 (79·9; 93·0) | 73·5 (68·5; 78·0) |
| Waist-hip ratio | <0·9 | 0·77 (0·74; 0·80) | 63·7 (58·0; 69·0) | 76·4 (72·5; 79·9) | 0·76 (0·70; 0·81) | 71·7 (62·1; 79·7) | 62·5 (57·2; 67·5) |
| HbA1c at diagnosis (mmol/mol) | >80 mmol/mol | 0·72 (0·68;0·75) | 81·5 (76·6; 85·6) | 50·7 (46·4; 55·0) | 0·79 (0·75;0·84) | 82·8 (74·1; 89·1) | 61·6 (56·3; 66·7) |
| Parent history of non-insulin-treated diabetes | No | 0·61 (0·58; 0·64) | 88·0 (83·8; 91·3) | 33·8 (29·9; 38·0) | 0·55 (0·50; 0·60) | 86·9 (78·7; 92·2) | 22·9 (18·7; 27·7) |
| Presence of osmotic symptoms | Yes | 0·61 (0·57; 0·64) | 94·5 (91·2; 96·6) | 26·5 (22·9; 30·5) | 0·65 (0·61; 0·70) | 91·9 (84·7; 95·9) | 38·4 (33·3; 43·7) |
| DKA | Yes | 0·58 (0·54; 0·61) | 18·8 (14·8; 23·7) | 96·5 (94·5; 97·8) | 0·62 (0·57; 0·66) | 25·3 (17·7; 34·7) | 97·9 (95·7; 99·0) |
| Sex | Female | 0·55 (0·52; 0·58) | 50·0 (44·3; 55·7) | 60·0 (55·7; 64·1) | 0·62 (0·58; 0·67) | 59·6 (49·7; 68·8) | 64·9 (59·6; 69·8) |
| Parent history of autoimmune disorders | Yes | 0·54 (0·51; 0·58) | 29·5 (24·5; 34·9) | 78·7 (75·0; 82·0) | 0·56 (0·51; 0·60) | 27·3 (19·4; 36·9) | 84·2 (79·9; 87·7) |
| Ethnicity** | / | 0·54 (0·52;0·56) | / | / | 0·52 (0·51;0·54) | / | / |
| Presence of other autoimmune disorder | Yes | 0·52 (0·49; 0·56) | 13·4 (9·9; 17·8) | 90·7 (87·9; 92·9) | 0·57 (0·52; 0·61) | 30·3 (22·1; 40·0) | 83·3 (79·0; 86·9) |

### Appendix Table 9:

**Univariate performance of routine clinical features in differentiating type 1 (T1D) and 2 diabetes (T2D) at diagnosis in participants aged 18-50 (StartRight: n=742; type 1=300) and Start Right Prime** (**n=389; type 1=88**), **using the restricted secondary definition of diabetes subtype.** Note age-at-diagnosis not shown due to restricted age range. ^†^Optimal threshold calculated by Youden’s Index. ^‡^Based on “optimal threshold”. *BMI-at-diagnosis or (where missing weight at diagnosis, StartRight: n=238; StartRight Prime: n=155) at recruitment visit. ** StartRight: Black ethnicity (n=24; type 1 = 4), Mixed ethnicity (n=10, type 1 = 3), Other ethnicity (n=8, type 1 = 3), South Asian ethnicity (n=34, type 1 = 7), White ethnicity (Reference group: n=666, type 1 = 283); StartRight Prime: Black ethnicity (n=5; type 1 = 1), Mixed ethnicity (n=3, type 1 = 0), Other ethnicity (n=4, type 1 = 0), South Asian ethnicity (n=6, type 1 = 0), White ethnicity (Reference group: n=371, type 1 = 87).

|  |  | **StartRight (n=742)** | | | **StartRight Prime (n=389)** | | |
| --- | --- | --- | --- | --- | --- | --- | --- |
| **Routine clinical features** | **Optimal Variable Threshold†** | **AUCROC (95% CI)** | **Sensitivity of T1D**^‡^ **(%; 95% CI)** | **Specificity of T1D**^‡^ **(%; 95% CI)** | **AUCROC (95% CI)** | **Sensitivity of T1D**^‡^ **(%; 95% CI)** | **Specificity of T1D**^‡^ **(%; 95% CI)** |
| BMI (kg/m^2^) | <28kg/m^2 †^ | 0·88 (0·86; 0·90) | 80·0 (75·1; 84·2) | 79·2 (75·2; 82·7) | 0·85 (0·80; 0·89) | 81·8 (72·4; 88·6) | 68·4 (63·0; 73·4) |
| Unintentional weight-loss | Yes | 0·78 (0·75; 0·81) | 83·3 (78·7; 87·1) | 72·2 (67·8; 76·2) | 0·83 (0·79; 0·86) | 88·6 (80·2; 93·8) | 76·7 (71·6; 81·2) |
| Waist-hip ratio | <0·94 | 0·77 (0·73; 0·80) | 62·0 (56·4; 67·3) | 77·6 (73·5; 81·3) | 0·75 (0·69; 0·81) | 80·1 (75·2; 84·2) | 58·0 (47·4; 67·8) |
| Age at diagnosis (years) | <37 years*** | 0·77 (0·74; 0·81) | 65·0 (59·4; 70·2) | 81·5 (77·6; 84·8) | 0·64 (0·57; 0·70) | NA | NA |
| HbA1c at diagnosis (mmol/mol) | >80 mmol/mol | 0·73 (0·70; 0·77) | 89·3 (85·3; 92·4) | 45·3 (40·7; 49·9) | 0·79 (0·74; 0·83) | 79·6 (69·9; 86·7) | 64·1 (58·5; 69·3) |
| Parent history of non-insulin-treated diabetes | No | 0·62 (0·59; 0·66) | 90·0 (86·1; 92·9) | 34·2 (29·9; 38·7) | 0·55 (0·50; 0·60) | 85·2 (76·2; 91·2) | 24·6 (20·1; 29·8) |
| Presence of osmotic symptoms | Yes | 0·61 (0·58; 0·65) | 94·0 (90·7; 96·2) | 28·5 (24·5; 32·9) | 0·67 (0·62; 0·72) | 93·2 (85·6; 96·9) | 40·9 (35·5; 46·5) |
| DKA | Yes | 0·58 (0·55; 0·62) | 19·7 (15·6; 24·6) | 96·8 (94·7; 98·1) | 0·63 (0·58; 0·67) | 27·3 (19·0; 37·5) | 98·0 (95·6; 99·1) |
| Sex | Female | 0·52 (0·49; 0·56) | 45·7 (40·1; 51·3) | 59·1 (54·4; 63·5) | 0·62 (0·57; 0·66) | 60·2 (49·7; 69·9) | 62·8 (57·2; 68·1) |
| Parent history of other autoimmune disease | Yes | 0·53 (0·50; 0·57) | 27·7 (22·9; 33·0) | 78·7 (74·7; 82·3) | 0·57 (0·52; 0·62) | 29·6 (21·0; 39·9) | 84·4 (79·8; 88·1) |
| Ethnicity** | / | 0·54 (0·52;0·56) | / | / | 0·52 (0·51;0·54) | / | / |
| Presence of other autoimmune disorder | Yes | 0·52 (0·49; 0·56) | 13·0 (9·6; 17·3) | 91·9 (88·9; 94·1) | 0·57 (0·52; 0·62) | 29·6 (21·0; 39·9) | 85·4 (80·9; 88·9) |

### Appendix Figure 5:

**Precision recall plots** for StartRight: A) clinical features only (n=833; type 1=302)) (AUCPR=0·913), B) clinical features + islet-autoantibodies (n=833; type 1=302)) (AUCPR=0·948), C) clinical features, islet-autoantibodies + TIDGRS (n=807; type 1=291)) (AUCPR=0·953) and D) clinical features + T1DGRS (n=807; type 1=291)) (AUCPR=0·933) models; and StartRight Prime: E) clinical features only (n=441; type 1=103) (AUCPR=0·8), F) clinical features + islet-autoantibodies (n=441; type 1=103)) (AUCPR=0·895), G) clinical features, islet-autoantibodies + TIDGRS (n=419; type 1=95) (AUCPR=0·919) and H) clinical features + T1DGRS (n=419; type 1=95) (AUCPR=0·849) models.

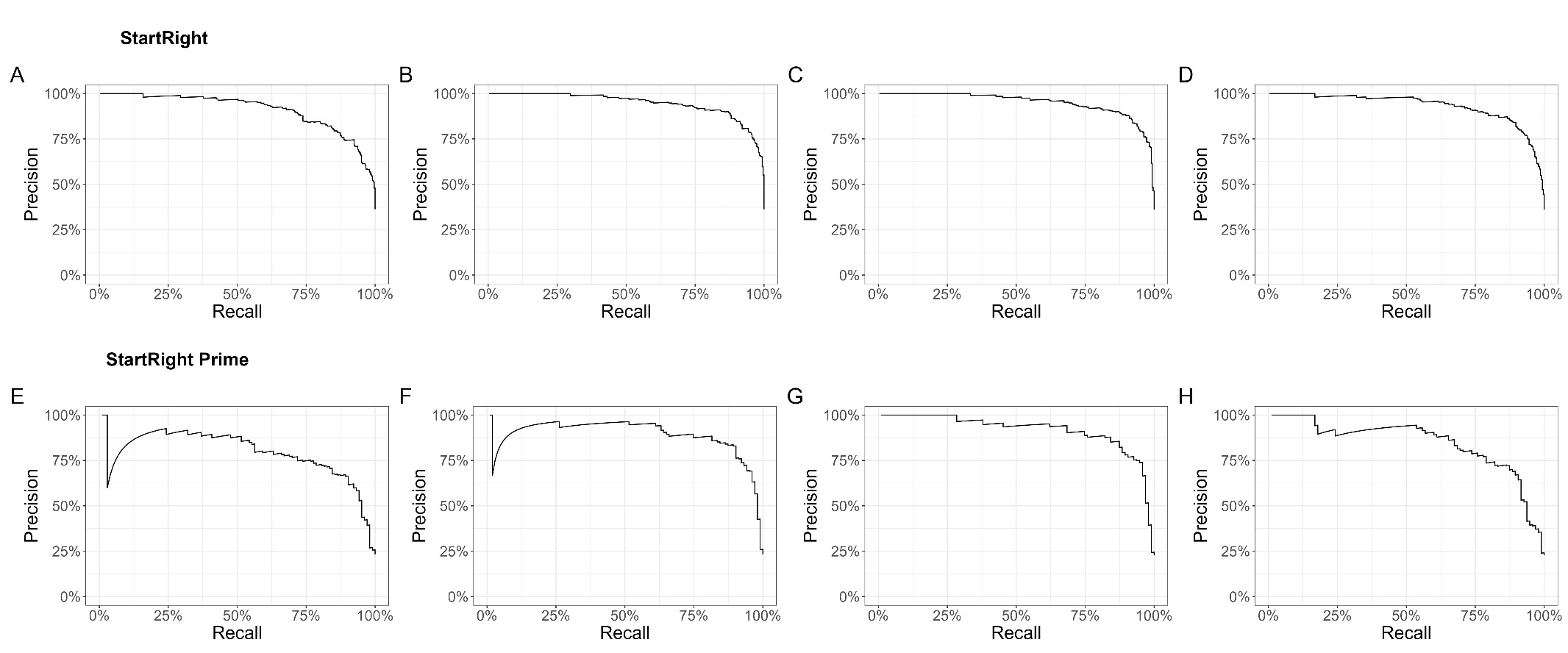

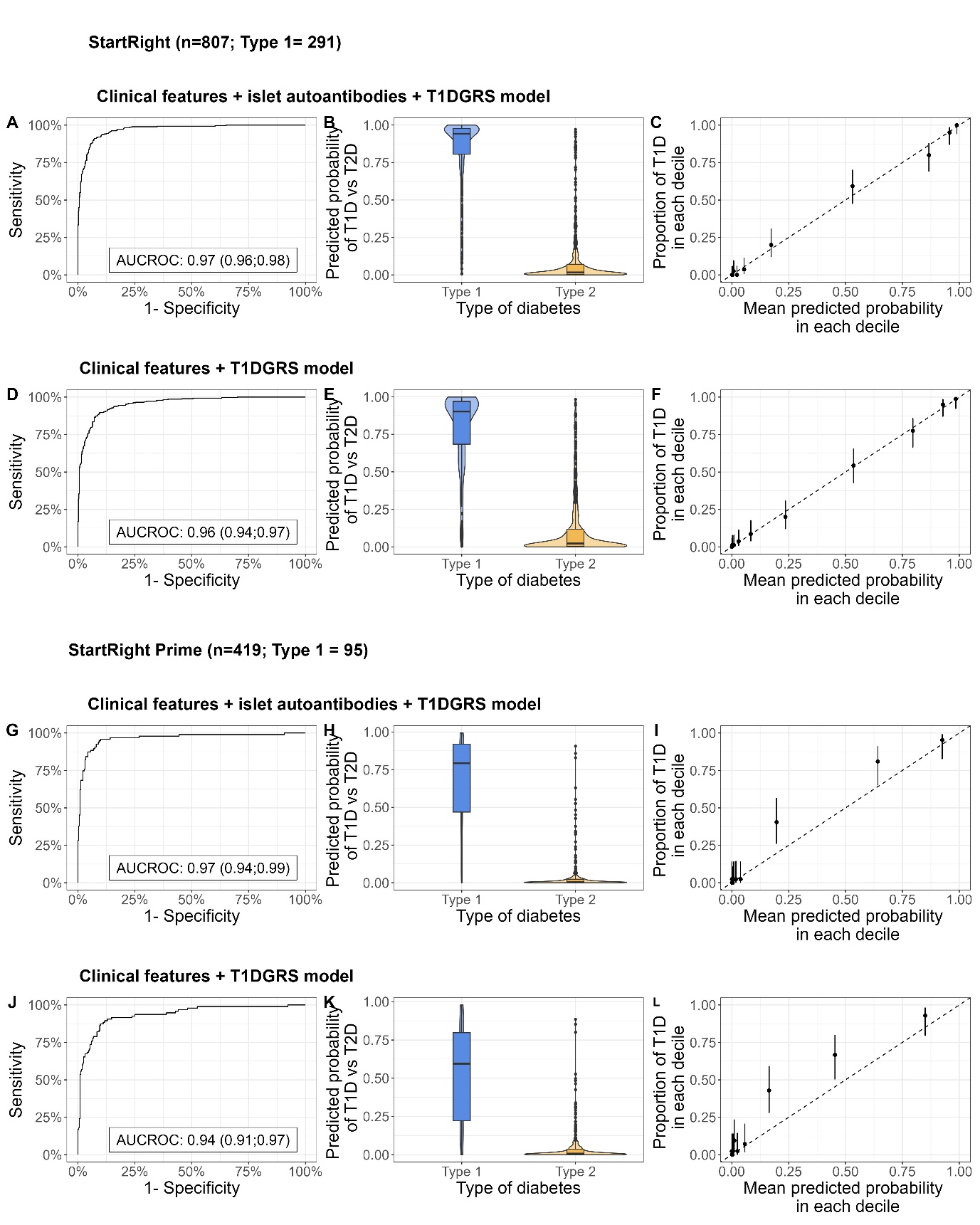

### Appendix Figure 6:

**Discrimination and calibration of diabetes classification models incorporating a type 1 diabetes genetic risk score (T1DGRS)**. Plots A, D, G, and J denote the Receiver Operating Characteristic (ROC) Curve for each respective model in StartRight and StartRight Prime respectively, with the Area Under the Curve (AUC) shown on the plot. Plots B, E, H, and K represent the predicted probability of the respective model by primary study outcome. Plots C, F, I, and L illustrate the calibration of each model, with deciles of model predicted probabilities of type 1 diabetes plotted against the observed proportion of type 1 diabetes defined by primary study outcome.

### Appendix Table 10:

Comparison of models in complete case data in StartRight (n=807), illustrating a comparable AUCROC, AIC and likelihood ratio tests between nested models. *Compared to clinical features only model; **Compared to clinical features and islet-autoantibodies model

| **Model** | **AUCROC (95% CI)** | **AIC** | **Likelihood ratio test*** | **Likelihood ratio test**** |
| --- | --- | --- | --- | --- |
| Clinical features only | 0·944 (0·930;0·959) | 492·4 |  |  |
| Clinical features + islet-autoantibodies | 0·957 (0·944;0·971) | 435·2 | p<0·001 |  |
| Clinical features + T1DGRS | 0·969 (0·959;0·979) | 387·4 | p<0·001 |  |
| Clinical features + islet-autoantibodies + T1DGRS | 0·971 (0·961;0·981) | 372·3 | / | p<0·001 |

### Appendix Table 11:

**Optimism of model performance indices using 1,000 bootstraps** for clinical features only; clinical features and islet-autoantibodies; clinical features, islet-autoantibodies and type 1 diabetes genetic risk score (T1DGRS); and clinical features and T1DGRS models. “Full” denotes the performance index calculated in StartRight, trained on the model developed on the original StartRight; “Bootstrap: training” refers to the mean performance index calculated in a bootstrapped sample of StartRight, based on a model developed on a bootstrapped sample of StartRight, across 1,000 bootstraps; “Bootstrap: test” refers to the mean performance index calculated in the original StartRight, based on a model developed on a bootstrapped sample of StartRight, across 1,000 bootstraps; Optimism refers to the difference between the training and test bootstrapped indices; and “Full: corrected” refers to the difference between the “Full” index and the “Optimism”. Somers’ Dxy rank correlation – correlation between predicted probabilities and outcome. When Dxy = 0, the model is making a random prediction; when Dxy = 1, the predictions are perfectly discriminating. Emax = “calibration error” – maximum absolute difference in predicted and loess calibrated probabilities. D = Discrimination Index = (LR χ2-1)/n. LR χ2 is null deviance – residual deviance. Null deviance is how well outcome is predicted by a model with only intercept, and residual deviance is how well outcome is predicted by model (deviance = discrepancy between observed probabilities and fitted probabilities). So D is how well the model improves prediction of the outcome. Unreliability index = difference in residual deviance between model and calibrated model (i.e. calibrated model is model where use predictions from first model to predict outcome so intercept and slope are 1 and 0). Q = Quality index = D-U. Brier Score = average squared differences between predicted values and outcome. G-index = gini’s mean difference calculated on log-odds scale a measure of the models’ predictive discrimination based only on the fitted values. Larger value = greater discrimination among outcomes.

| **Performance metric** | **Full** | **Bootstrap: training** | **Bootstrap: test** | **Optimism** | **Full: corrected** |
| --- | --- | --- | --- | --- | --- |
| **Clinical features only** | | | | | |
| AUCROC | 0·943 | 0·944 | 0·938 | 0·006 | 0·937 |
| Somers’ Dxy | 0·886 | 0·891 | 0·879 | 0·012 | 0·874 |
| Nagelkere’s R^2^ | 0·706 | 0·717 | 0·695 | 0·021 | 0·685 |
| Calibration Intercept | 0·000 | 0·000 | -0·012 | 0·012 | -0·012 |
| Calibration Slope | 1·000 | 1·000 | 0·934 | 0·066 | 0·934 |
| Emax | 0·000 | 0·000 | 0·017 | 0·017 | 0·017 |
| Discrimination Index (D) | 0·724 | 0·740 | 0·708 | 0·032 | 0·691 |
| Unreliability Index (U) | -0·002 | -0·002 | 0·002 | -0·004 | 0·002 |
| Quality Index (Q) | 0·726 | 0·742 | 0·706 | 0·036 | 0·690 |
| Brier Score | 0·091 | 0·089 | 0·094 | -0·005 | 0·097 |
| g-index | 3·831 | 4·011 | 3·734 | 0·277 | 3·555 |
| g-index on the probability scale | 0·410 | 0·412 | 0·407 | 0·005 | 0·404 |
| **Clinical features + islet-autoantibodies** | | | | | |
| AUCROC | 0·969 | 0·969 | 0·963 | 0·006 | 0·962 |
| Somers’ Dxy | 0·937 | 0·941 | 0·930 | 0·011 | 0·926 |
| Nagelkere’s R^2^ | 0·795 | 0·806 | 0·783 | 0·023 | 0·773 |
| Calibration Intercept | 0·000 | 0·000 | -0·014 | 0·014 | -0·014 |
| Calibration Slope | 1·000 | 1·000 | 0·916 | 0·084 | 0·916 |
| Emax | 0·000 | 0·000 | 0·022 | 0·022 | 0·022 |
| Discrimination Index (D) | 0·868 | 0·886 | 0·847 | 0·039 | 0·829 |
| Unreliability Index (U) | -0·002 | -0·002 | 0·003 | -0·005 | 0·003 |
| Quality Index (Q) | 0·870 | 0·889 | 0·844 | 0·044 | 0·826 |
| Brier Score | 0·067 | 0·064 | 0·070 | -0·006 | 0·074 |
| g-index | 4·458 | 4·760 | 4·339 | 0·421 | 4·037 |
| g-index on the probability scale | 0·433 | 0·434 | 0·430 | 0·005 | 0·428 |
| **Clinical features, islet-autoantibodies + T1DGRS** | | | | | |
| AUCROC | 0·971 | 0·971 | 0·965 | 0·006 | 0·965 |
| Somers’ Dxy | 0·942 | 0·946 | 0·935 | 0·011 | 0·930 |
| Nagelkere’s R^2^ | 0·807 | 0·817 | 0·795 | 0·023 | 0·784 |
| Calibration Intercept | 0·000 | 0·000 | -0·010 | 0·010 | -0·010 |
| Calibration Slope | 1·000 | 1·000 | 0·914 | 0·086 | 0·914 |
| Emax | 0·000 | 0·000 | 0·022 | 0·022 | 0·022 |
| Discrimination Index (D) | 0·887 | 0·906 | 0·866 | 0·041 | 0·846 |
| Unreliability Index (U) | -0·002 | -0·002 | 0·003 | -0·005 | 0·003 |
| Quality Index (Q) | 0·890 | 0·909 | 0·863 | 0·046 | 0·844 |
| Brier Score | 0·062 | 0·059 | 0·065 | -0·006 | 0·069 |
| g-index | 4·508 | 4·844 | 4·402 | 0·442 | 4·066 |
| g-index on the probability scale | 0·434 | 0·436 | 0·431 | 0·005 | 0·429 |
| **Clinical features + T1DGRS** | | | | | |
| AUCROC | 0·957 | 0·959 | 0·952 | 0·006 | 0·951 |
| Somers’ Dxy | 0·915 | 0·919 | 0·909 | 0·010 | 0·905 |
| Nagelkere’s R^2^ | 0·757 | 0·765 | 0·746 | 0·019 | 0·738 |
| Calibration Intercept | 0·000 | 0·000 | -0·009 | 0·009 | -0·009 |
| Calibration Slope | 1·000 | 1·000 | 0·933 | 0·067 | 0·933 |
| Emax | 0·000 | 0·000 | 0·017 | 0·017 | 0·017 |
| Discrimination Index (D) | 0·802 | 0·816 | 0·785 | 0·032 | 0·770 |
| Unreliability Index (U) | -0·002 | -0·002 | 0·002 | -0·004 | 0·002 |
| Quality Index (Q) | 0·804 | 0·819 | 0·783 | 0·036 | 0·768 |
| Brier Score | 0·076 | 0·074 | 0·078 | -0·005 | 0·080 |
| g-index | 4·361 | 4·569 | 4·239 | 0·329 | 4·032 |
| g-index on the probability scale | 0·423 | 0·424 | 0·420 | 0·004 | 0·418 |

### Appendix Figure 7:

**Stability of model predictions using 100 bootstraps**, for A) clinical features only; B) clinical features and islet-autoantibodies; C) clinical features, islet-autoantibodies and T1DGRS; and D) clinical features and T1DGRS models in StartRight (A and B: n=833, type 1=302; C and D: n=807, type 1=291). X axis denotes model probabilities from the developed model in the whole dataset, y axis denotes the model probabilities from 100 bootstrapped samples of the dataset, within which the model is redeveloped. Grey dots denote model probabilities. Dashed lines indicate the 95% Confidence Interval per individual in the whole dataset across all bootstrapped model probabilities.

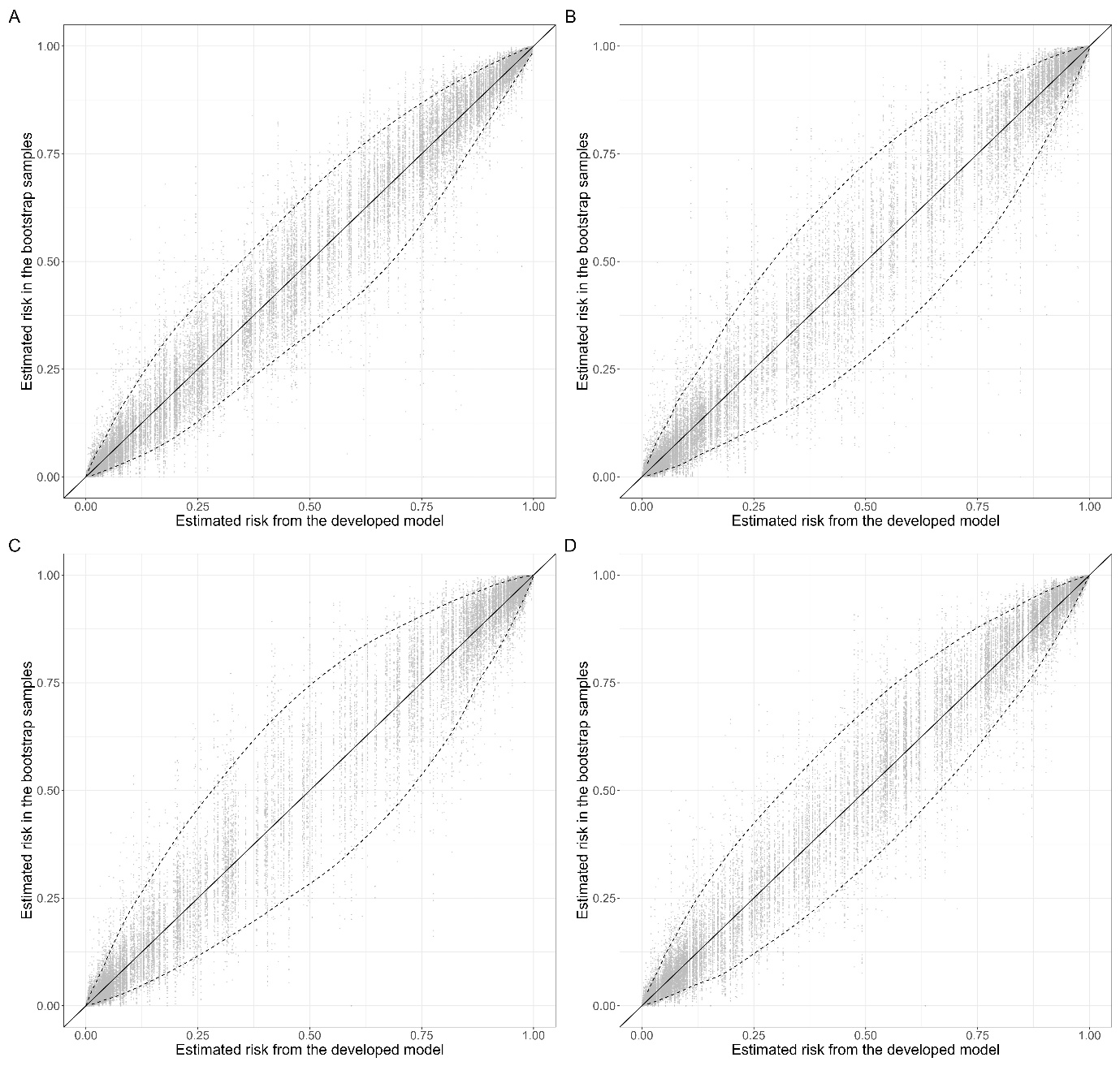

### Appendix Table 12:

**Model beta coefficients and Odds Ratios all four models developed in StartRight.** All numerical features are scaled.

|  | **Clinical features only**  (n=833, type 1=302) | | | **Clinical features + islet-autoantibodies**  **(**n=833, type 1=302) | | | **Clinical features + islet-autoantibodies + T1GRS (**n=807, type 1=291) | | | **Clinical features + T1GRS**  **(**n=807, type 1=291) | | |
| --- | --- | --- | --- | --- | --- | --- | --- | --- | --- | --- | --- | --- |
| **Model Feature** | **B**  **coefficient**  **(95% CI)** | **Odds Ratio (95% CI)** | **p** | **B**  **coefficient (95% CI)** | **Odds Ratio (95% CI)** | **p** | **B coefficient (95% CI)** | **Odds Ratio (95% CI)** | **p** | **B**  **coefficient (95% CI)** | **Odds Ratio (95% CI)** | **p** |
| **Intercept** | -2·29 (-3·12, -1·51) | 0·10 (0·04, 0·22) | <0·001 | -3·51 (-4·58, -2·53) | 0·03 (0·01, 0·08) | <0·001 | -3·36 (-4·46, -2·34) | 0·03 (0·01, 0·10) | <0·001 | -2·51 (-3·43, -1·65) | 0·08 (0·03, 0·19) | <0·001 |
| **Age-at-diagnosis (years)** | -0·90 (-1·15, -0·66) | 0·41 (0·32, 0·51) | <0·001 | -0·73 (-1·02, -0·45) | 0·48 (0·36, 0·64) | <0·001 | -0·62 (-0·92, -0·33) | 0·54 (0·40, 0·72) | <0·001 | -0·76 (-1·03, -0·5) | 0·47 (0·36, 0·61) | <0·001 |
| **BMI-at-diagnosis (kg/m2)** | -2·07 (-2·51, -1·68) | 0·13 (0·08, 0·19) | <0·001 | -1·79 (-2·27, -1·36) | 0·17 (0·10, 0·26) | <0·001 | -1·85 (-2·38, -1·37) | 0·16 (0·09, 0·25) | <0·001 | -2·14 (-2·63, -1·69) | 0·12 (0·07, 0·18) | <0·001 |
| **HbA1c-at-diagnosis (mmol/mol)** | 0·11 (-0·17, 0·38) | 1·11 (0·84, 1·47) | 0·44 | 0·10 (-0·23, 0·43) | 1·11 (0·80, 1·53) | 0·54 | 0·03 (-0·3, 0·37) | 1·03 (0·74, 1·45) | 0·85 | 0·04 (-0·26, 0·35) | 1·04 (0·77, 1·41) | 0·78 |
| **Male sex** | -0·92 (-1·42, -0·43) | 0·40 (0·24, 0·65) | <0·001 | -0·84 (-1·42, -0·27) | 0·43 (0·24, 0·77) | 0·004 | -0·82 (-1·44, -0·22) | 0·44 (0·24, 0·80) | 0·008 | -0·81 (-1·35, -0·28) | 0·45 (0·26, 0·76) | 0·003 |
| **Parent history of non-insulin-treated diabetes** | -1·08 (-1·65, -0·52) | 0·34 (0·19, 0·59) | <0·001 | -0·70 (-1·34, -0·07) | 0·50 (0·26, 0·94) | 0·032 | -0·69 (-1·38, -0·02) | 0·50 (0·25, 0·98) | 0·044 | -1·17 (-1·81, -0·55) | 0·31 (0·16, 0·58) | <0·001 |
| **Presence of DKA** | 0·78 (0·02, 1·59) | 2·18 (1·02, 4·91) | 0·051 | 0·70 (-0·18, 1·6) | 2·02 (0·84, 4·94) | 0·12 | 0·48 (-0·44, 1·43) | 1·62 (0·64, 4·19) | 0·31 | 0·68 (-0·17, 1·58) | 1·97 (0·84, 4·84) | 0·13 |
| **Unintentional weight-loss** | 1·44 (0·88, 2·01) | 4·20 (2·40, 7·48) | <0·001 | 1·31 (0·67, 1·98) | 3·71 (1·95, 7·21) | <0·001 | 1·04 (0·36, 1·74) | 2·83 (1·43, 5·68) | 0·003 | 1·14 (0·53, 1·77) | 3·12 (1·69, 5·85) | <0·001 |
| **Presence of osmotic symptoms** | 1·02 (0·24, 1·84) | 2·78 (1·27, 6·32) | 0·012 | 0·97 (0·03, 1·98) | 2·65 (1·03, 7·24) | 0·050 | 1·03 (0·03, 2·08) | 2·79 (1·04, 7·97) | 0·048 | 1·10 (0·25, 2·0) | 3·01 (1·28, 7·41) | 0·014 |
| **Black ethnicity** | -1·41 (-2·92, -0·08) | 0·24 (0·05, 0·93) | 0·049 | -0·86 (-2·56, 0·61) | 0·43 (0·08, 1·84) | 0·28 | -0·05 (-1·84, 1·48) | 0·95 (0·16, 4·38) | 0·95 | -0·15 (-1·81, 1·29) | 0·86 (0·16, 3·65) | 0·85 |
| **Other/Mixed ethnicity** | -1·43 (-2·9, -0·06) | 0·24 (0·06, 0·94) | 0·049 | -0·65 (-2·39, 0·84) | 0·52 (0·09, 2·32) | 0·43 | -1·28 (-3·14, 0·39) | 0·28 (0·04, 1·48) | 0·16 | -1·81 (-3·33, -0·38) | 0·16 (0·04, 0·68) | 0·016 |
| **South Asian ethnicity** | -0·84 (-2·18, 0·42) | 0·43 (0·11, 1·51) | 0·21 | -0·39 (-1·81, 0·91) | 0·67 (0·16, 2·47) | 0·57 | -0·54 (-2·12, 0·90) | 0·58 (0·12, 2·45) | 0·48 | -0·83 (-2·41, 0·64) | 0·43 (0·09, 1·89) | 0·29 |
| **Presence of other autoimmune disease** | 0·87 (0·19, 1·58) | 2·40 (1·20, 4·86) | 0·014 | 0·73 (-0·10, 1·56) | 2·07 (0·90, 4·75) | 0·085 | 0·77 (-0·09, 1·63) | 2·15 (0·92, 5·10) | 0·079 | 1·03 (0·28, 1·79) | 2·80 (1·33, 6·0) | 0·007 |
| **Single positive islet-autoantibody** |  |  |  | 2·14 (1·52, 2·77) | 8·47 (4·58, 16·04) | <0·001 | 1·81 (1·14, 2·49) | 6·09 (3·12, 12·09) | <0·001 |  |  |  |
| **Two positive islet-autoantibodies** |  |  |  | 2·76 (1·93, 3·68) | 15·88 (6·90, 39·73) | <0·001 | 2·25 (1·38, 3·20) | 9·51 (3·98, 24·54) | <0·001 |  |  |  |
| **Three positive islet-autoantibodies** |  |  |  | 3·76 (2·82, 4·85) | 43·06 (16·72, 128·23) | <0·001 | 3·17 (2·22, 4·26) | 23·83 (9·22, 71·04) | <0·001 |  |  |  |
| **T1DGRS** |  |  |  |  |  |  | 0·7 (0·36, 1·05) | 2·01 (1·43, 2·87) | <0·001 | 1·11 (0·81, 1·44) | 3·04 (2·24, 4·21) | <0·001 |

### Appendix Table 13:

**Model discrimination and accuracy in identifying type 1 diabetes; defined by restricted secondary diabetes subtype outcome (three-year insulin-use, C-peptide and islet-autoantibodies) using a 50% cut-off; compared with, islet-autoantibodies and type 1 diabetes genetic risk score alone.** *Threshold is defined by Youden’s Index.

| **Model** | **AUCROC (95% CI)** | **Threshold** | **Sensitivity (95% CI)** | **Specificity (95% CI)** | **Predictive value T1D (95% CI)** | **Predictive value T2D (95% CI)** | **Accuracy (95% CI)** |
| --- | --- | --- | --- | --- | --- | --- | --- |
| **StartRight** |  |  |  |  |  |  |  |
| Clinical features only  (n=763 [T1D N=310 (40·6%)]) | 0·95 (0·93, 0·96) | 50% | 80·5% (75·6; 84·6) | 91·0% (88·2; 93·1) | 83·5% (78·8; 87·3) | 89·1% (86·2; 91·5) | 87·2% (84·7; 89·3) |
| Clinical features and islet-autoantibodies  (n=763 [T1D N=310 (40·6%)]) | 0·99 (0·99, 1·00) | 50% | 86·8% (82·4; 90·1) | 94·5% (92·3; 96·2) | 90·0% (86·0; 93·0) | 92·6% (90·1; 94·5) | 91·7% (89·6; 93·4) |
| Clinical features, islet-autoantibodies and T1DGRS  (n=743 [T1D N=301(40·5%)]) | 0·99 (0·99, 1·00) | 50% | 87·6% (83·3; 90·9) | 94·0% (91·6; 95·7) | 89·2% (85·0; 92·3) | 93·1% (90·6; 95·0) | 91·7% (89·6; 93·4) |
| Clinical features and T1DGRS  (n=743 [T1D N=301(40·5%)]) | 0·97 (0·96, 0·98) | 50% | 87·3% (82·9; 90·7) | 92·3% (89·6; 94·3) | 86·4% (82·0; 89·9) | 92·8% (90·2; 94·7) | 90·5% (88·2; 92·3) |
| T1DGRS  (n=743 [T1D N=301 (40·5%)]) | 0·84 (0·81; 0·87) | 0·628* | 79·7% (74·8; 83·9) | 76·5% (72·3; 80·2) | 69·8% (64·7; 74·4) | 84·7% (80·8; 87·9) | 77·8% (74·7; 80·6) |
| **StartRight Prime** |  |  |  |  |  |  |  |
| Clinical features only  (n= 395 [T1D N=90 (22·8%)]) | 0·94 (0·92, 0·97) | 50% | 80·5% (75·6; 84·6) | 91·0% (88·2; 93·1) | 83·5% (78·8; 87·3) | 89·1% (86·2; 91·5) | 87·2% (84·7; 89·3) |
| Clinical features and islet-autoantibodies  (n= 395 [T1D N=90 (22·8%)]) | 0·99 (0·99, 1·00) | 50% | 86·8% (82·4; 90·1) | 94·5% (92·3; 96·2) | 90·0% (86·0; 93·0) | 92·6% (90·1; 94·5) | 91·7% (89·6; 93·4) |
| Clinical features, islet-autoantibodies and T1DGRS  (n= 382 [T1D N= 86 (22·5%)]) | 0·99 (0·98, 1·00) | 50% | 87·6% (83·3; 90·9) | 94·0% (91·6; 95·7) | 89·2% (85·0; 92·3) | 93·1% (90·6; 95·0) | 91·7% (89·6; 93·4) |
| Clinical features and T1DGRS  (n= 382 [T1D N= 86 (22·5%)]) | 0·96 (0·93, 0·98) | 50% | 87·3% (82·9; 90·7) | 92·3% (89·6; 94·3) | 86·4% (82·0; 89·9) | 92·8% (90·2; 94·7) | 90·5% (88·2; 92·3) |
| T1DGRS  (n= 382 [T1D N= 86 (22·5%)]) | 0·80 (0·75; 0·85) | 0·66* | 72·1% (61·7; 80·5) | 76·7% (71·5; 81·2) | 47·3% (38·9; 55·8) | 90·4% (86·1; 93·5) | 75·7% (71·1; 79·7) |

### Appendix Figure 8:

**Model performance of clinical features only and clinical features and T1DGRS models to predict type 1 (verses type 2) diabetes defined by study secondary (restricted) subtype definition (**combining three-year insulin-use, islet-autoantibodies and C-peptide, see methods), in StartRight and StartRight Prime. Plots A, D, G, and J denote the Receiver Operating Characteristic (ROC) Curve for each respective model in StartRight and StartRight Prime respectively, with the Area Under the Curve (AUC) shown on the plot (AUCROC (95%CI)). Plots B, E, H, and K represent the predicted probability of the respective model by restricted secondary study outcome. Plots C, F, I, and L illustrate the calibration of each model, with deciles of model predicted probabilities of type 1 diabetes plotted against the observed proportion of type 1 diabetes defined by restricted secondary study outcome.

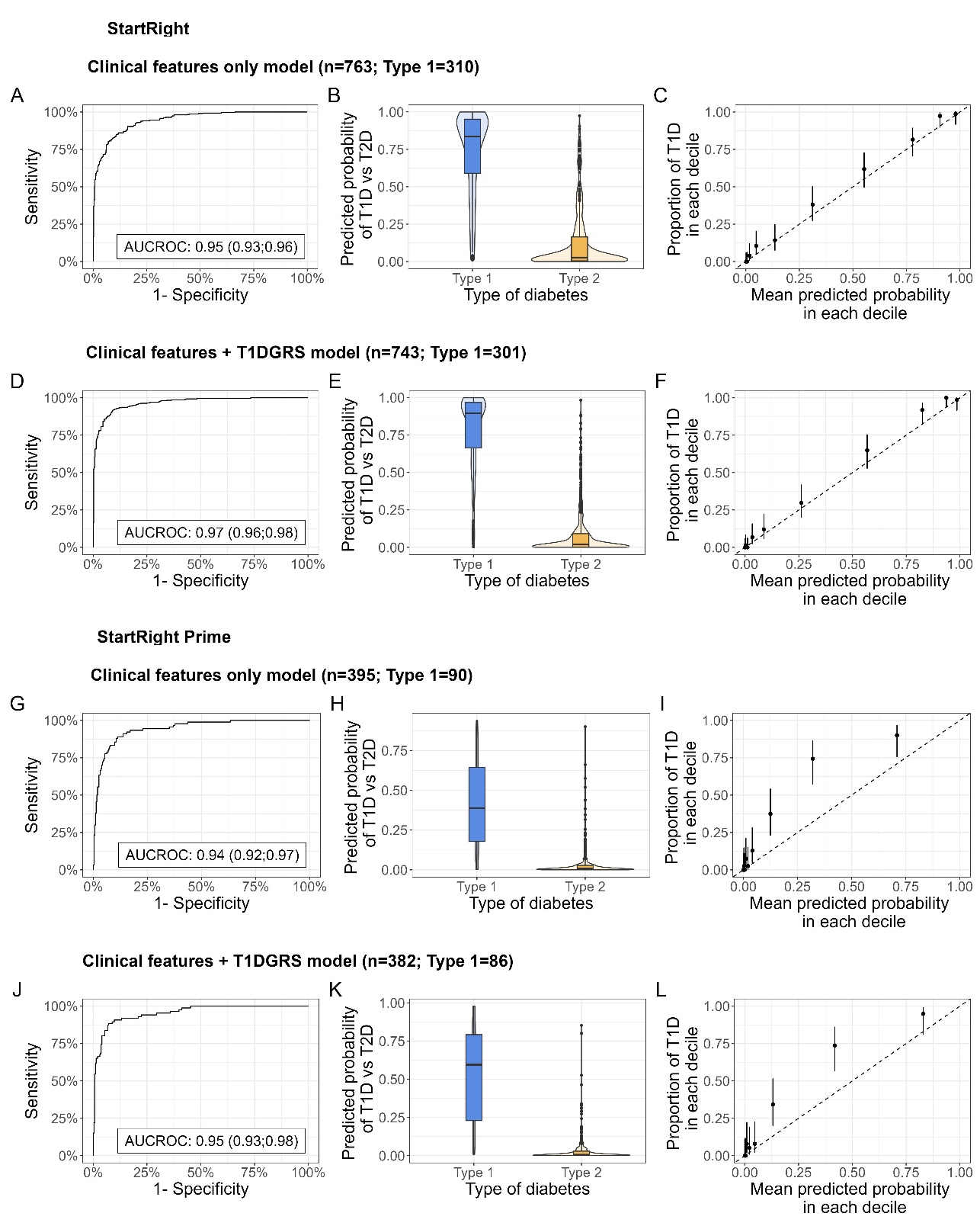

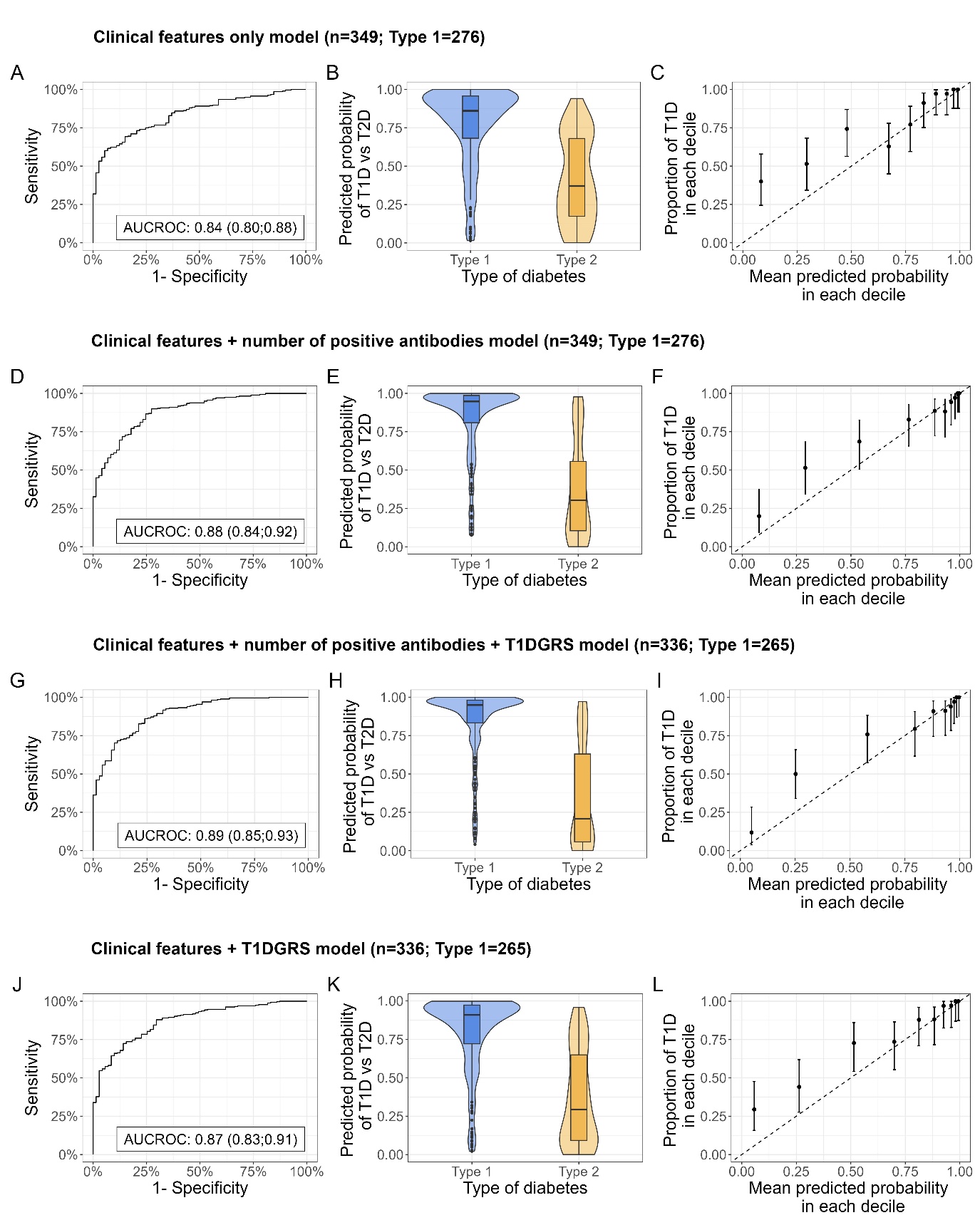

### Appendix Figure 9:

**Model separation and calibration in individuals clinically diagnosed with type 1 diabetes**, in StartRight, for clinical features only, clinical features and islet-autoantibodies, clinical features, islet-autoantibodies and T1DGRS, and clinical features and T1DGRS models. Plots A, D, G, and J denote the Receiver Operating Characteristic (ROC) Curve for each respective model in StartRight, with the Area Under the Curve (AUC) shown on the plot (AUCROC (95%CI). Plots B, E, H, and K represent the predicted probability of the respective model by primary study outcome. Plots C, F, I, and L illustrate the calibration of each model, with deciles of model predicted probabilities of type 1 diabetes plotted against the observed proportion of type 1 diabetes defined by primary study outcome.

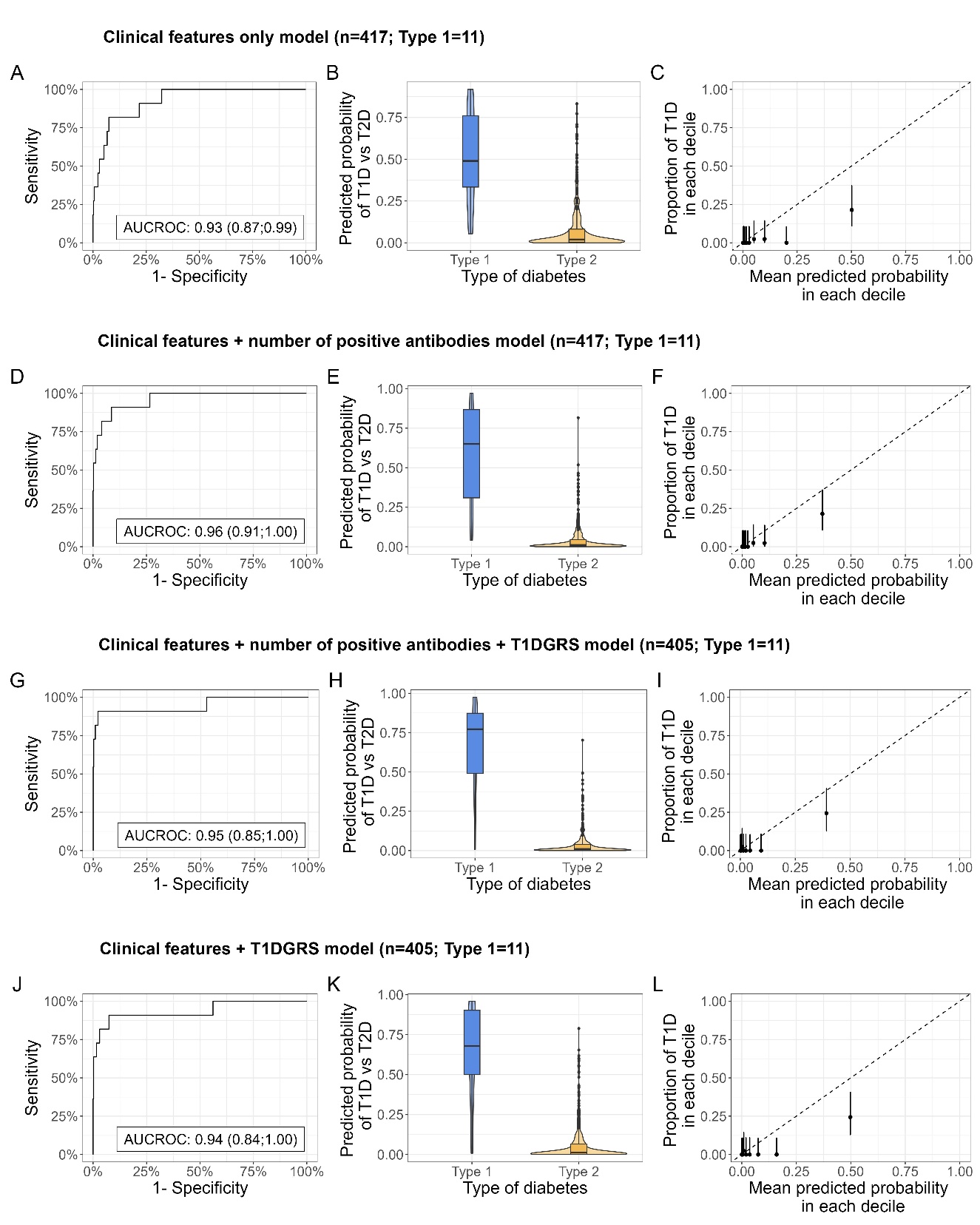

### Appendix Figure 10:

**Model separation and calibration in individuals clinically diagnosed with type 2 diabetes**, in StartRight, for clinical features only, clinical features and islet-autoantibodies, clinical features, islet-autoantibodies and T1DGRS, and clinical features and T1DGRS models. Plots A, D, G, and J denote the Receiver Operating Characteristic (ROC) Curve for each respective model in StartRight, with the Area Under the Curve (AUC) shown on the plot (AUCROC (95%CI)). Plots B, E, H, and K represent the predicted probability of the respective model by primary study outcome. Plots C, F, I, and L illustrate the calibration of each model, with deciles of model predicted probabilities of type 1 diabetes plotted against the observed proportion of type 1 diabetes defined by primary study outcome.

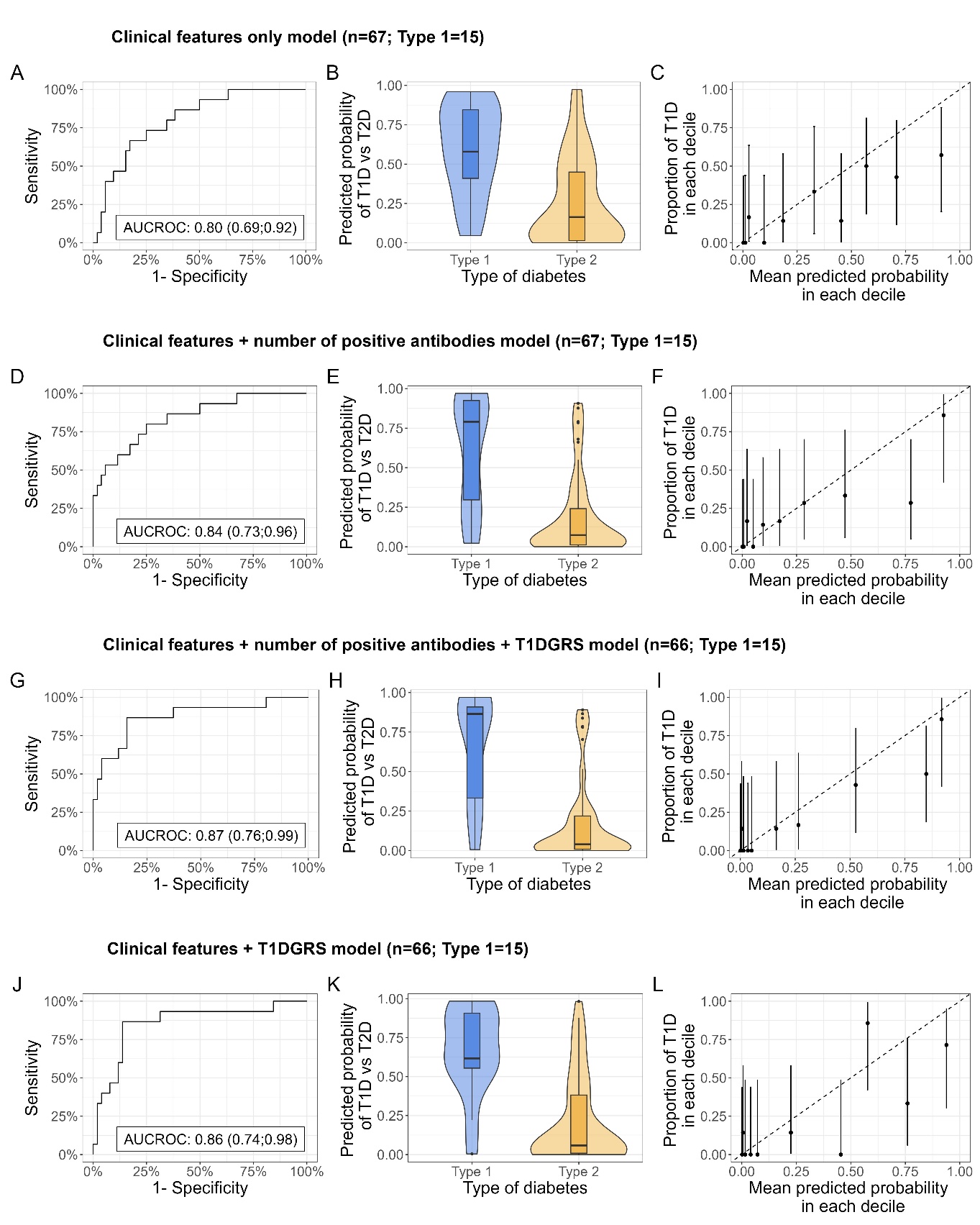

### Appendix Figure 11:

**Model separation and calibration in individuals with uncertain clinical diagnosis**, in StartRight, for clinical features only, clinical features and islet-autoantibodies, clinical features, islet-autoantibodies and T1DGRS, and clinical features and T1DGRS models. Plots A, D, G, and J denote the Receiver Operating Characteristic (ROC) Curve for each respective model in StartRight, with the Area Under the Curve (AUC) shown on the plot (AUCROC (95%CI)). Plots B, E, H, and K represent the predicted probability of the respective model by primary study outcome. Plots C, F, I, and L illustrate the calibration of each model, with deciles of model predicted probabilities of type 1 diabetes plotted against the observed proportion of type 1 diabetes defined by primary study outcome.

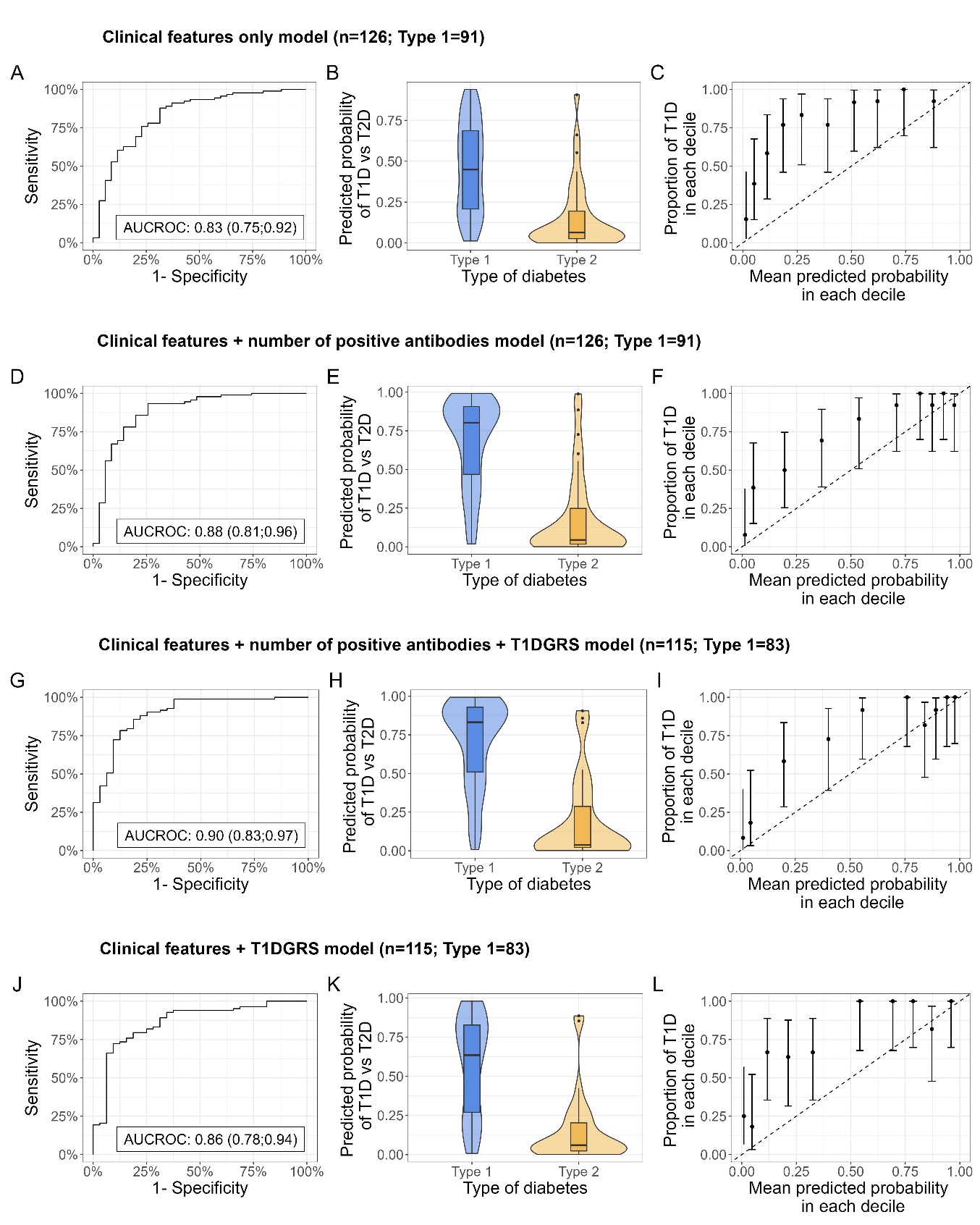

### Appendix Figure 12:

**Model separation and calibration in individuals clinically diagnosed with type 1 diabetes**, in StartRight Prime, for clinical features only, clinical features and islet-autoantibodies, clinical features, islet-autoantibodies and T1DGRS, and clinical features and T1DGRS models. Plots A, D, G, and J denote the Receiver Operating Characteristic (ROC) Curve for each respective model in StartRight Prime, with the Area Under the Curve (AUC) shown on the plot (AUCROC (95%CI)). Plots B, E, H, and K represent the predicted probability of the respective model by primary study outcome. Plots C, F, I, and L illustrate the calibration of each model, with deciles of model predicted probabilities of type 1 diabetes plotted against the observed proportion of type 1 diabetes defined by primary study outcome.

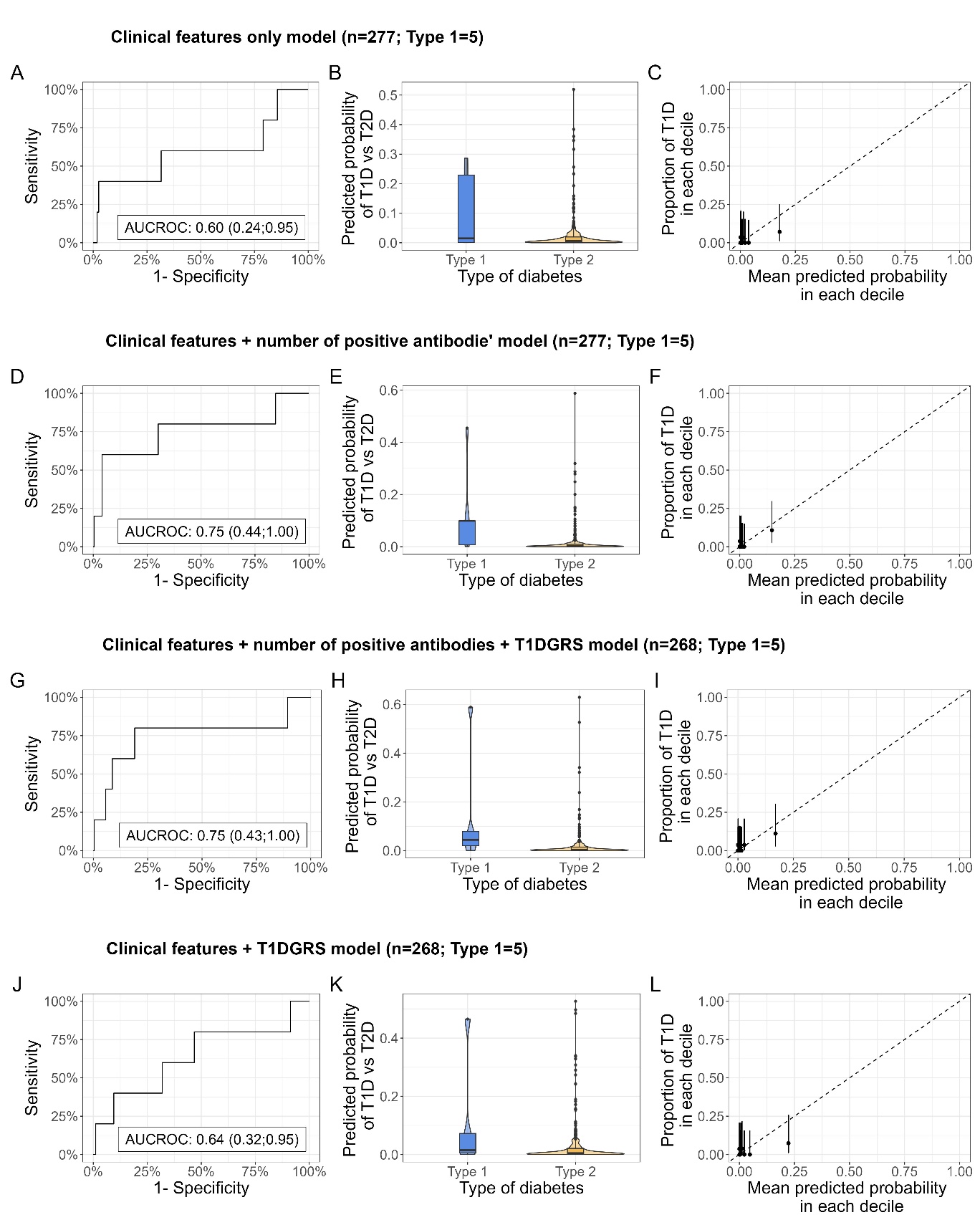

### Appendix Figure 13:

**Model separation and calibration in individuals clinically diagnosed with type 2 diabetes**, in StartRight Prime, for clinical features only, clinical features and islet-autoantibodies, clinical features, islet-autoantibodies and T1DGRS, and clinical features and T1DGRS models. Plots A, D, G, and J denote the Receiver Operating Characteristic (ROC) Curve for each respective model in StartRight Prime, with the Area Under the Curve (AUC) shown on the plot (AUCROC (95% CI)). Plots B, E, H, and K represent the predicted probability of the respective model by primary study outcome. Plots C, F, I, and L illustrate the calibration of each model, with deciles of model predicted probabilities of type 1 diabetes plotted against the observed proportion of type 1 diabetes defined by primary study outcome.

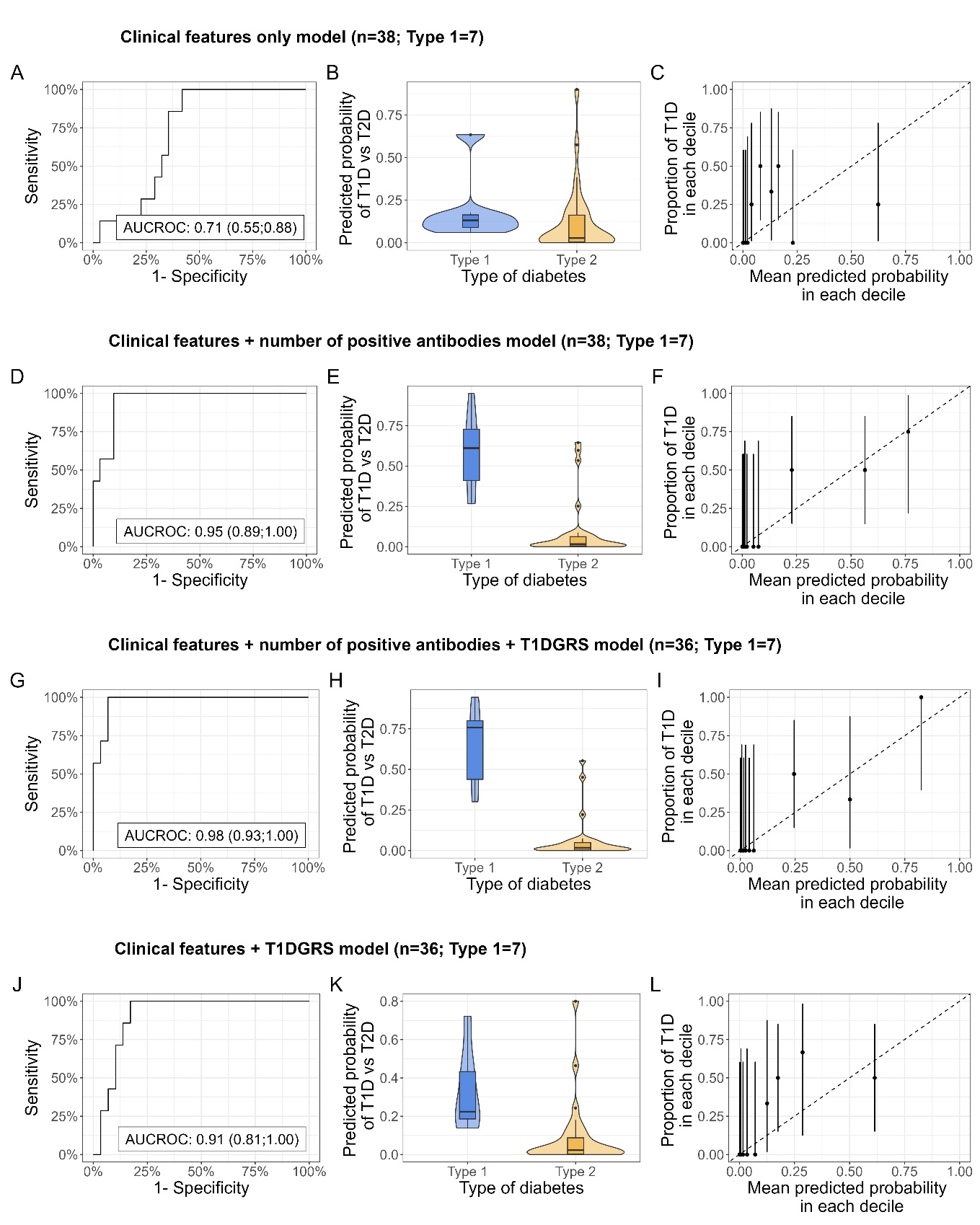

### Appendix Figure 14:

**Model separation and calibration in individuals with uncertain clinical diagnosis**, in StartRight Prime, for clinical features only, clinical features and islet-autoantibodies, clinical features, islet-autoantibodies and T1DGRS, and clinical features and T1DGRS models. Plots A, D, G, and J denote the Receiver Operating Characteristic (ROC) Curve for each respective model in StartRight Prime, with the Area Under the Curve (AUC) shown on the plot (AUCROC (95% CI)). Plots B, E, H, and K represent the predicted probability of the respective model by primary study outcome. Plots C, F, I, and L illustrate the calibration of each model, with deciles of model predicted probabilities of type 1 diabetes plotted against the observed proportion of type 1 diabetes defined by primary study outcome.

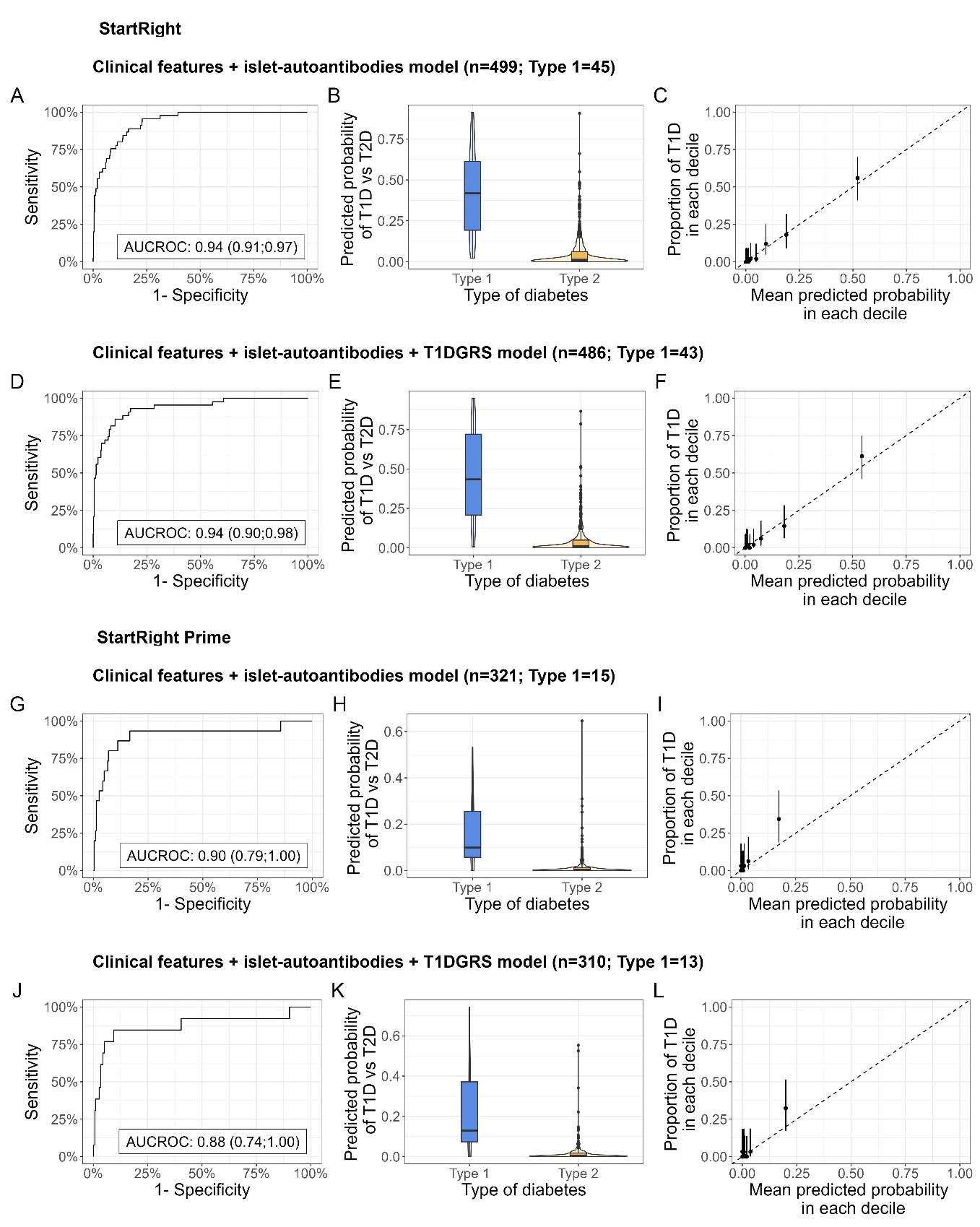

### Appendix Figure 15:

**Model performance of clinical features and islet-autoantibodies and clinical features, islet-autoantibodies and T1DGRS models in individuals that are islet-autoantibody negative**, in StartRight and StartRight Prime. Plots A, D, G, and J denote the Receiver Operating Characteristic (ROC) Curve for each respective model in StartRight and StartRight Prime respectively, with the Area Under the Curve (AUC) shown on the plot (AUCROC (95%CI)). Plots B, E, H, and K represent the predicted probability of the respective model by primary study outcome. Plots C, F, I, and L illustrate the calibration of each model, with deciles of model predicted probabilities of type 1 diabetes plotted against the observed proportion of type 1 diabetes defined by primary study outcome.

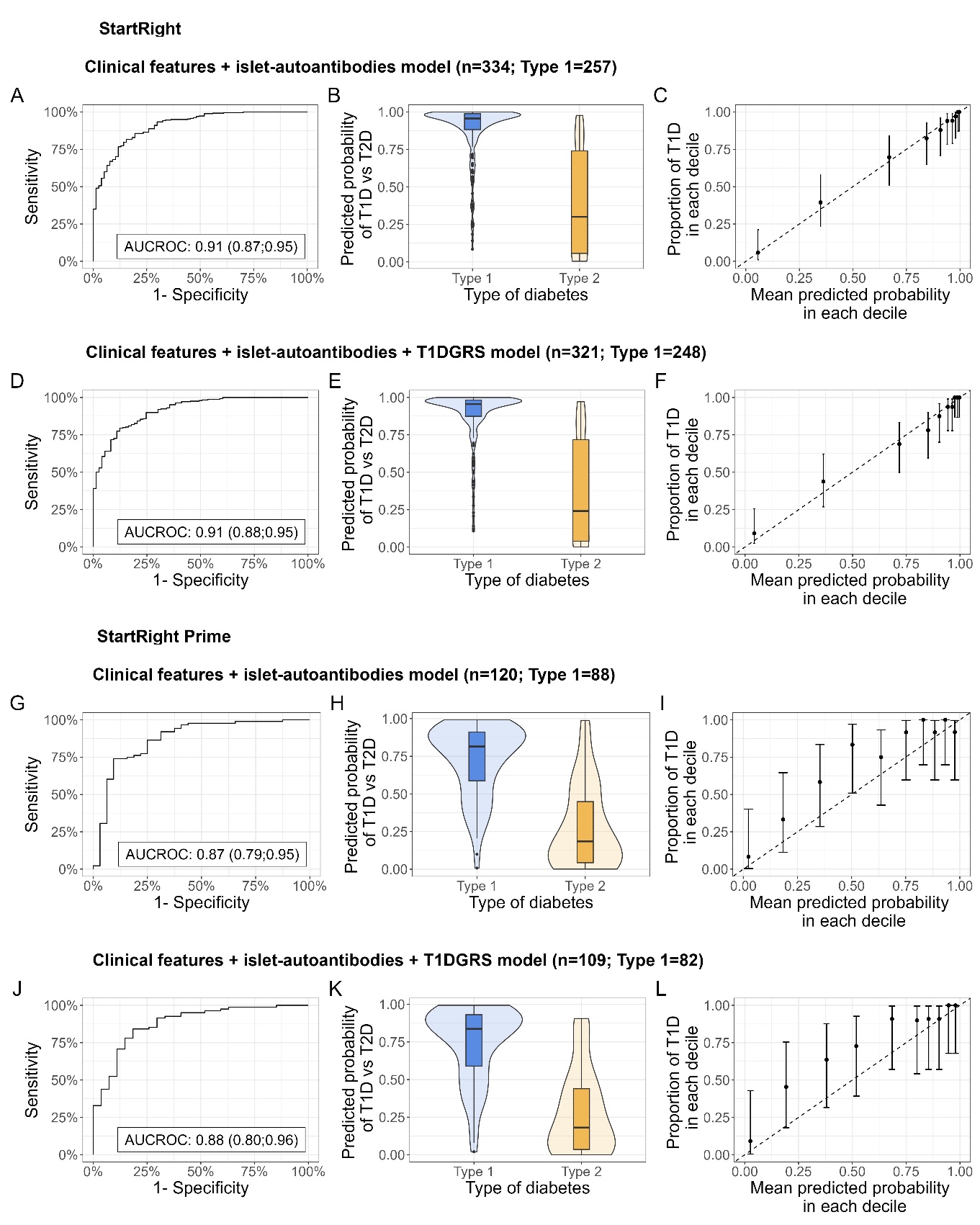

### Appendix Figure 16:

**Model performance of clinical features and islet-autoantibodies and clinical features, islet-autoantibodies and T1DGRS models in individuals that are islet-autoantibody positive**, in StartRight and StartRight Prime. Plots A, D, G, and J denote the Receiver Operating Characteristic (ROC) Curve for each respective model in StartRight and StartRight Prime respectively, with the Area Under the Curve (AUC) shown on the plot (AUCROC (95%CI)). Plots B, E, H, and K represent the predicted probability of the respective model by primary study outcome. Plots C, F, I, and L illustrate the calibration of each model, with deciles of model predicted probabilities of type 1 diabetes plotted against the observed proportion of type 1 diabetes defined by primary study outcome.

### Appendix Table 14:

**Model discrimination and accuracy in identifying type 1 diabetes (T1D) from type 2 diabetes (T2D) (defined by primary study outcome) using a binary (50% probability) cut off; compared with islet-autoantibodies and type 1 diabetes genetic risk score alone (T1DGRS).** *Threshold is defined by Youden’s Index; Sensitivity, specificity, predictive value of T1D and T2D and accuracy are calculated based on this threshold.

| **Model** | **AUCROC (95% CI)** | **Threshold** | **Sensitivity of T1D**  **(95% CI)** | **Specificity of T1D**  **(95% CI)** | **Predictive value T1D (95% CI)** | **Predictive value T2D (95% CI)** | **Accuracy (95% CI)** |
| --- | --- | --- | --- | --- | --- | --- | --- |
| **StartRight** |  |  |  |  |  |  |  |
| Clinical features only  (n=833 [T1D N=302 (36·3%)]) | 0·94 (0·93, 0·96) | 50% | 80·5% (75·6; 84·6) | 91·0% (88·2; 93·1) | 83·5% (78·8; 87·3) | 89·1% (86·2; 91·5) | 87·2% (84·7; 89·3) |
| Clinical features and islet-autoantibodies’  (n=833 [T1D N=302 (36·3%)]) | 0·97 (0·96, 0·90) | 50% | 86·8% (82·4; 90·1) | 94·5% (92·3; 96·2) | 90·0% (86·0; 93·0) | 92·6% (90·1; 94·5) | 91·7% (89·6; 93·4) |
| Clinical features, islet-autoantibodies and T1DGRS  (n=807 [T1D N=291(36·1%)]) | 0·97 (0·96, 0·98) | 50% | 87·6% (83·3; 90·9) | 94·0% (91·6; 95·7) | 89·2% (85·0; 92·3) | 93·1% (90·6; 95·0) | 91·7% (89·6; 93·4) |
| Clinical features and T1DGRS  (n=807 [T1D N=291(36·1%)]) | 0·96 (0·94, 0·97) | 50% | 87·3% (82·9; 90·7) | 92·3% (89·6; 94·3) | 86·4% (82·0; 89·9) | 92·8% (90·2; 94·7) | 90·5% (88·2; 92·3) |
| ≥1 positive islet-autoantibody (of GAD, IA2, ZnT8)  (n=833 [T1D N=302 (36·3%)]) | 0·85 (0·83; 0·88) | Positive | 85·1% (80·6; 88·7) | 85·5% (82·2; 88·3) | 77·0% (72·1; 81·2) | 91·0% (88·1; 93·2) | 85·4% (82·8; 87·6) |
| T1DGRS  (n=807 [T1D N=291 (36·1%)]) | 0·81 (0·78; 0·84) | 0·628* | 79·0% (74·0; 83·3) | 72·5% (68·5; 76·2) | 61·8% (56·8; 66·6) | 86·0% (82·4; 88·9) | 74·8% (71·7; 77·7) |
| **StartRight Prime** |  |  |  |  |  |  |  |
| Clinical features only  (n=441 [T1D N=103 (23·4%)]) | 0·93 (0·90, 0·96) | 50% | 80·5% (75·6; 84·6) | 91·0% (88·2; 93·1) | 83·5% (78·8; 87·3) | 89·1% (86·2; 91·5) | 87·2% (84·7; 89·3) |
| Clinical features and islet-autoantibodies  (n=441 [T1D N=103 (23·4%)]) | 0·97 (0·94, 0·99) | 50% | 86·8% (82·4; 90·1) | 94·5% (92·3; 96·2) | 90·0% (86·0; 93·0) | 92·6% (90·1; 94·5) | 91·7% (89·6; 93·4) |
| Clinical features, islet-autoantibodies and T1DGRS  (n=419 [T1D N=95 (22·7%)]) | 0·97 (0·94, 0·99) | 50% | 87·6% (83·3; 90·9) | 94·0% (91·6; 95·7) | 89·2% (85·0; 92·3) | 93·1% (90·6; 95·0) | 91·7% (89·6; 93·4) |
| Clinical features and T1DGRS  (n=419 [T1D N=95 (22·7%)]) | 0·94 (0·91, 0·97) | 50% | 87·3% (82·9; 90·7) | 92·3% (89·6; 94·3) | 86·4% (82·0; 89·9) | 92·8% (90·2; 94·7) | 90·5% (88·2; 92·3) |
| ≥1 positive islet-autoantibody (of GAD, IA2, ZnT8)  (n=441 [T1D N=103 (23·4%)]) | 0·88 (0·85; 0·91) | Positive | 85·4 % (77·2; 91·0) | 90·5% (86·9; 93·2) | 73·3% (64·7; 80·5) | 95·3% (92·4; 97·2) | 89·3% (86·1; 91·9) |
| T1DGRS  (n=419 [T1D N=95 (22·7%)]) | 0·78 (0·73; 0·83) | 0·66* | 72·6% (62·8; 80·6) | 74·4% (69·4; 78·8) | 45·4% (37·7; 53·4) | 90·3% (86·1; 93·3) | 74·0% (69·6; 78·0) |

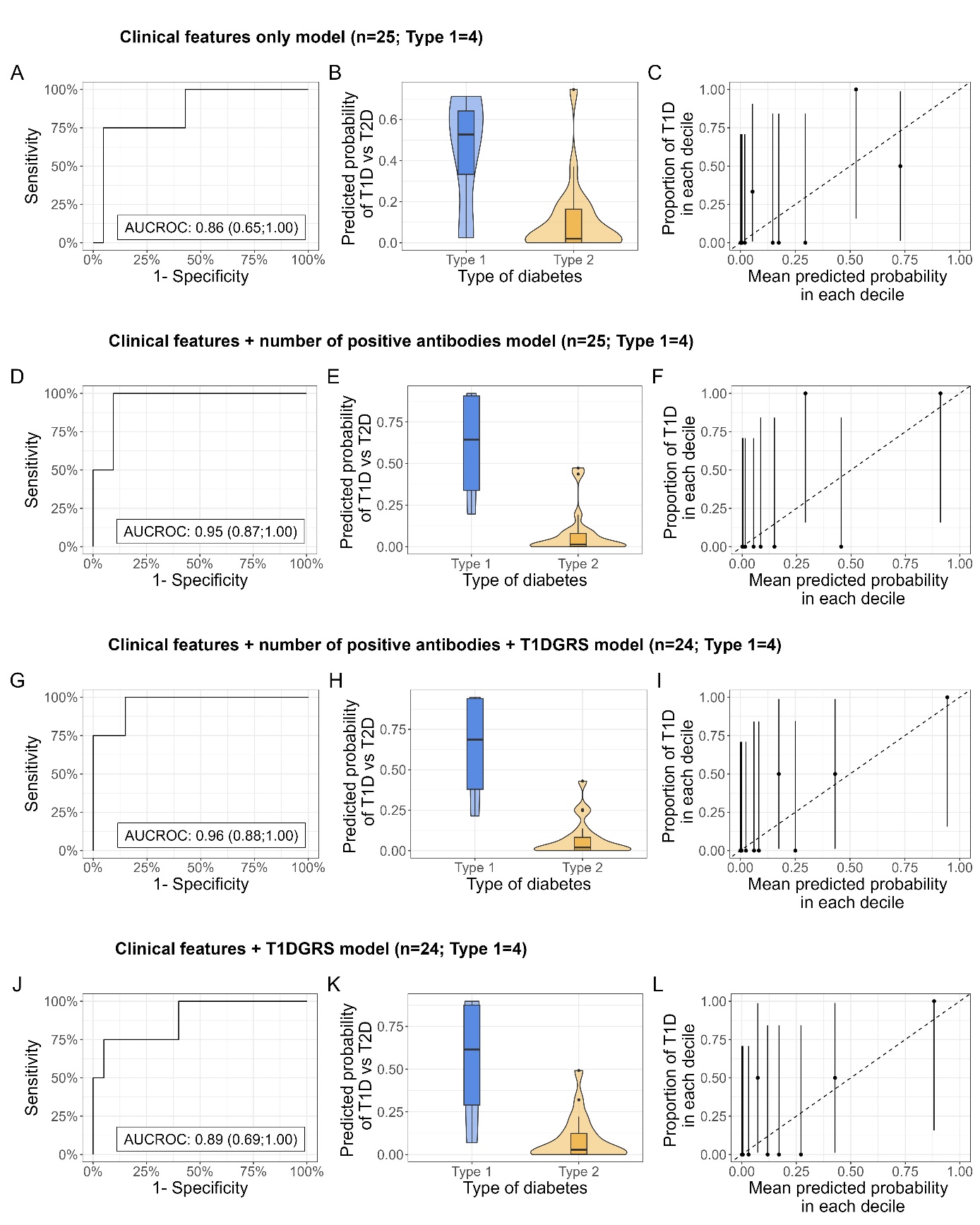

### Appendix Figure 17:

**Model separation and calibration in individuals of self-reported black ancestry**, for clinical features only, clinical features and islet-autoantibodies, clinical features, islet-autoantibodies and T1DGRS, and clinical features and T1DGRS models. Plots A, D, G, and J denote the Receiver Operating Characteristic (ROC) Curve for each respective model in StartRight, with the Area Under the Curve (AUC) shown on the plot (AUCROC (95% CI)). Plots B, E, H, and K represent the predicted probability of the respective model by primary study outcome. Plots C, F, I, and L illustrate the calibration of each model, with deciles of model predicted probabilities of type 1 diabetes plotted against the observed proportion of type 1 diabetes defined by primary study outcome.

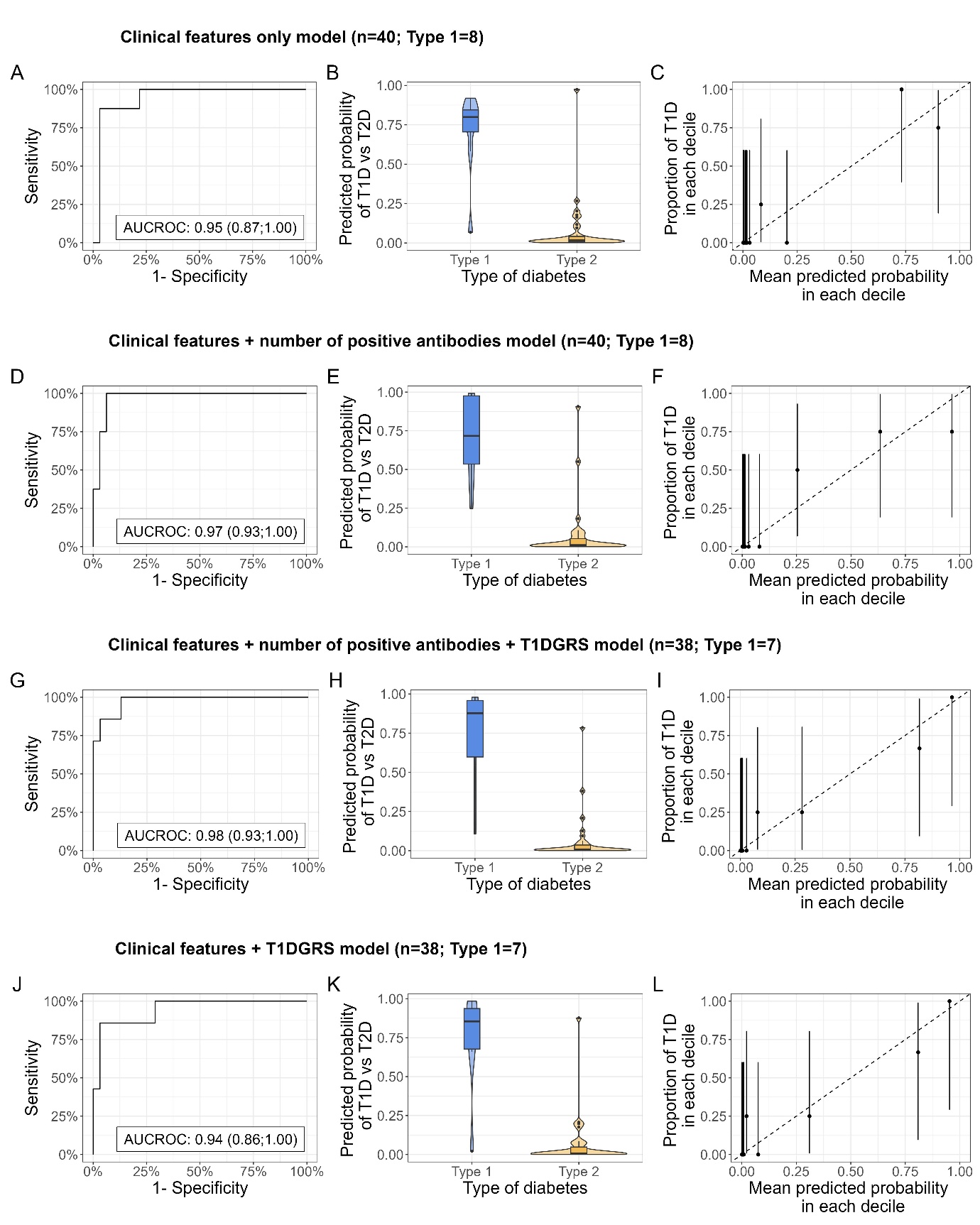

### Appendix Figure 18:

**Model separation and calibration in individuals of self-reported south Asian ancestry**, in StartRight, for clinical features only, clinical features and islet-autoantibodies, clinical features, islet-autoantibodies and T1DGRS, and clinical features and T1DGRS models. Plots A, D, G, and J denote the Receiver Operating Characteristic (ROC) Curve for each respective model in StartRight, with the Area Under the Curve (AUC) shown on the plot (AUCROC (95% CI)). Plots B, E, H, and K represent the predicted probability of the respective model by primary study outcome. Plots C, F, I, and L illustrate the calibration of each model, with deciles of model predicted probabilities of type 1 diabetes plotted against the observed proportion of type 1 diabetes defined by primary study outcome.

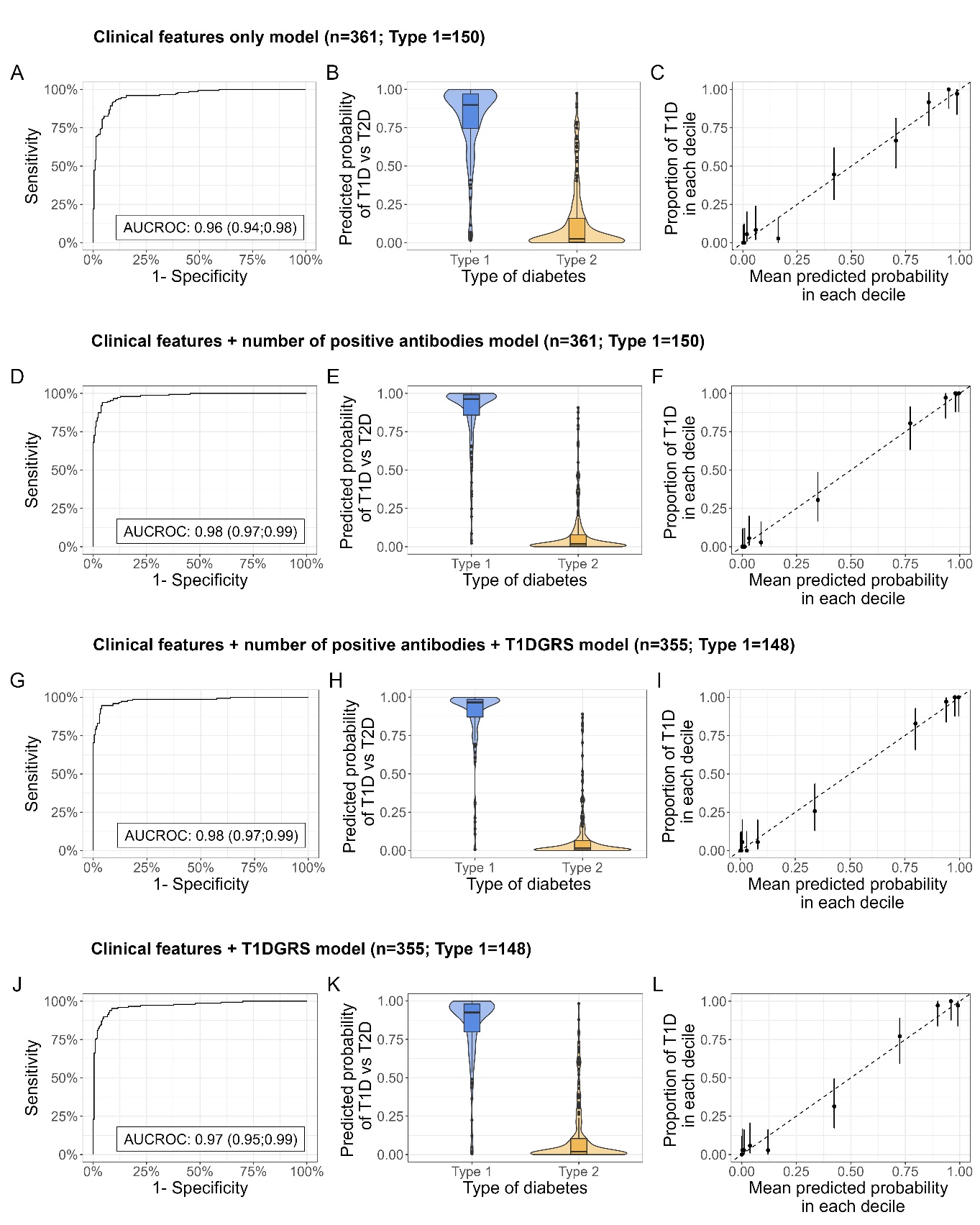

### Appendix Figure 19:

**Model separation and calibration in female individuals**, in StartRight, for clinical features only, clinical features and islet-autoantibodies, clinical features, islet-autoantibodies and T1DGRS, and clinical features and T1DGRS models. Plots A, D, G, and J denote the Receiver Operating Characteristic (ROC) Curve for each respective model in StartRight, with the Area Under the Curve (AUC) shown on the plot (AUCROC (95% CI)). Plots B, E, H, and K represent the predicted probability of the respective model by primary study outcome. Plots C, F, I, and L illustrate the calibration of each model, with deciles of model predicted probabilities of type 1 diabetes plotted against the observed proportion of type 1 diabetes defined by primary study outcome.

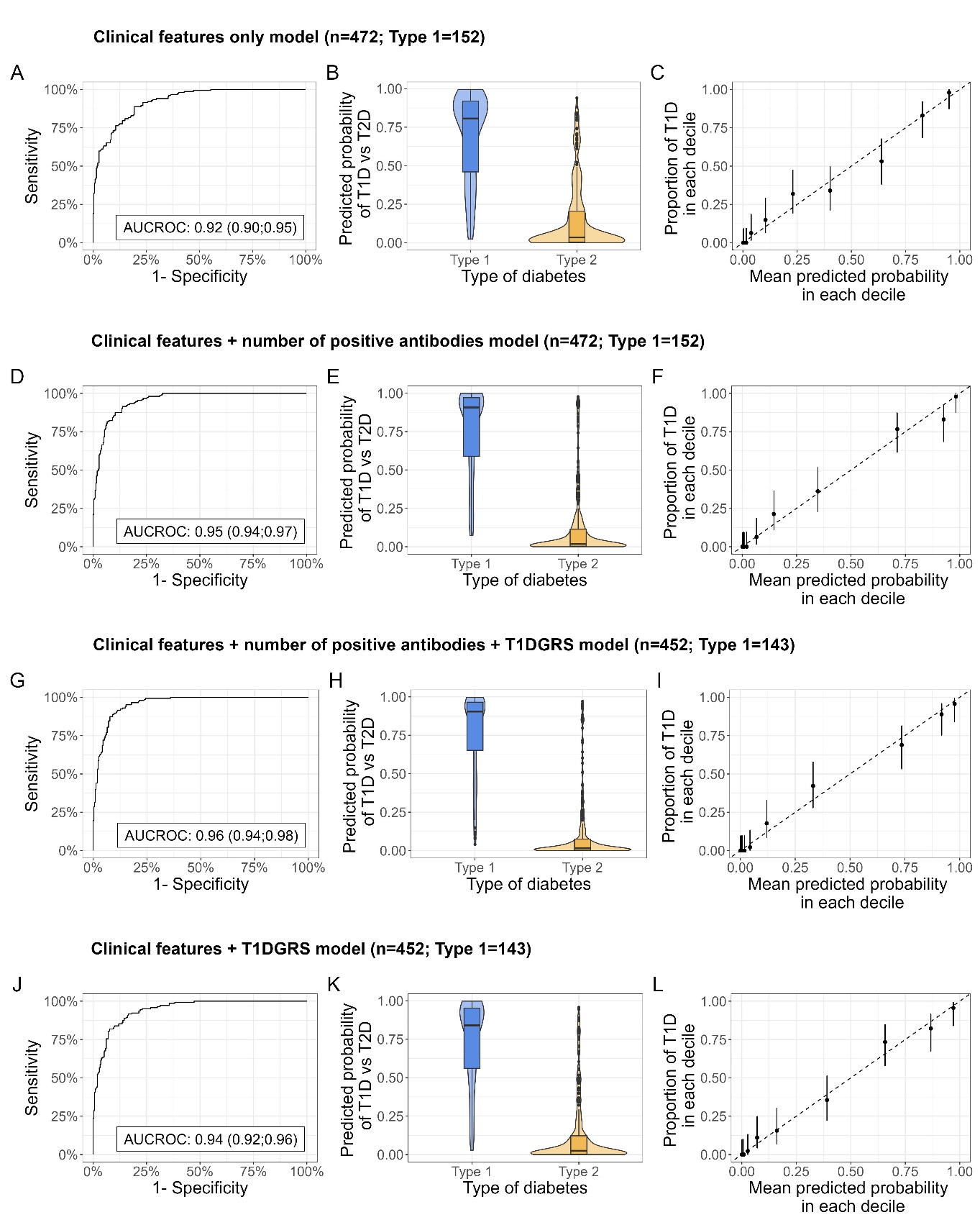

### Appendix Figure 20:

**Model separation and calibration in male individuals**, in StartRight, for clinical features only, clinical features and islet-autoantibodies, clinical features, islet-autoantibodies and T1DGRS, and clinical features and T1DGRS models. Plots A, D, G, and J denote the Receiver Operating Characteristic (ROC) Curve for each respective model in StartRight, with the Area Under the Curve (AUC) shown on the plot (AUCROC (95% CI)). Plots B, E, H, and K represent the predicted probability of the respective model by primary study outcome. Plots C, F, I, and L illustrate the calibration of each model, with deciles of model predicted probabilities of type 1 diabetes plotted against the observed proportion of type 1 diabetes defined by primary study outcome.

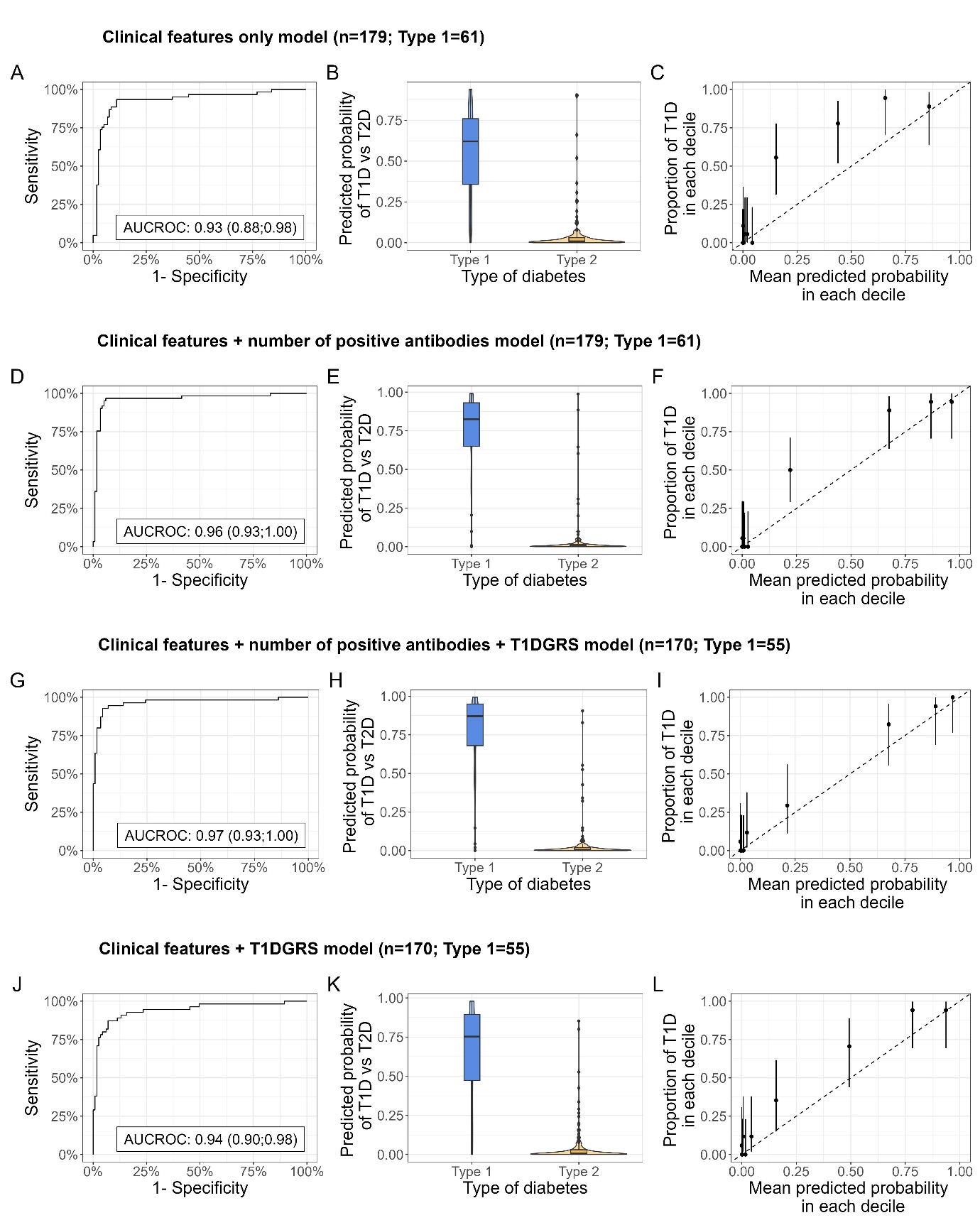

### Appendix Figure 21:

**Model separation and calibration in female individuals**, in StartRight Prime, for clinical features only, clinical features and islet-autoantibodies, clinical features, islet-autoantibodies and T1DGRS, and clinical features and T1DGRS models. Plots A, D, G, and J denote the Receiver Operating Characteristic (ROC) Curve for each respective model in StartRight, with the Area Under the Curve (AUC) shown on the plot (AUCROC (95% CI)). Plots B, E, H, and K represent the predicted probability of the respective model by primary study outcome. Plots C, F, I, and L illustrate the calibration of each model, with deciles of model predicted probabilities of type 1 diabetes plotted against the observed proportion of type 1 diabetes defined by primary study outcome.

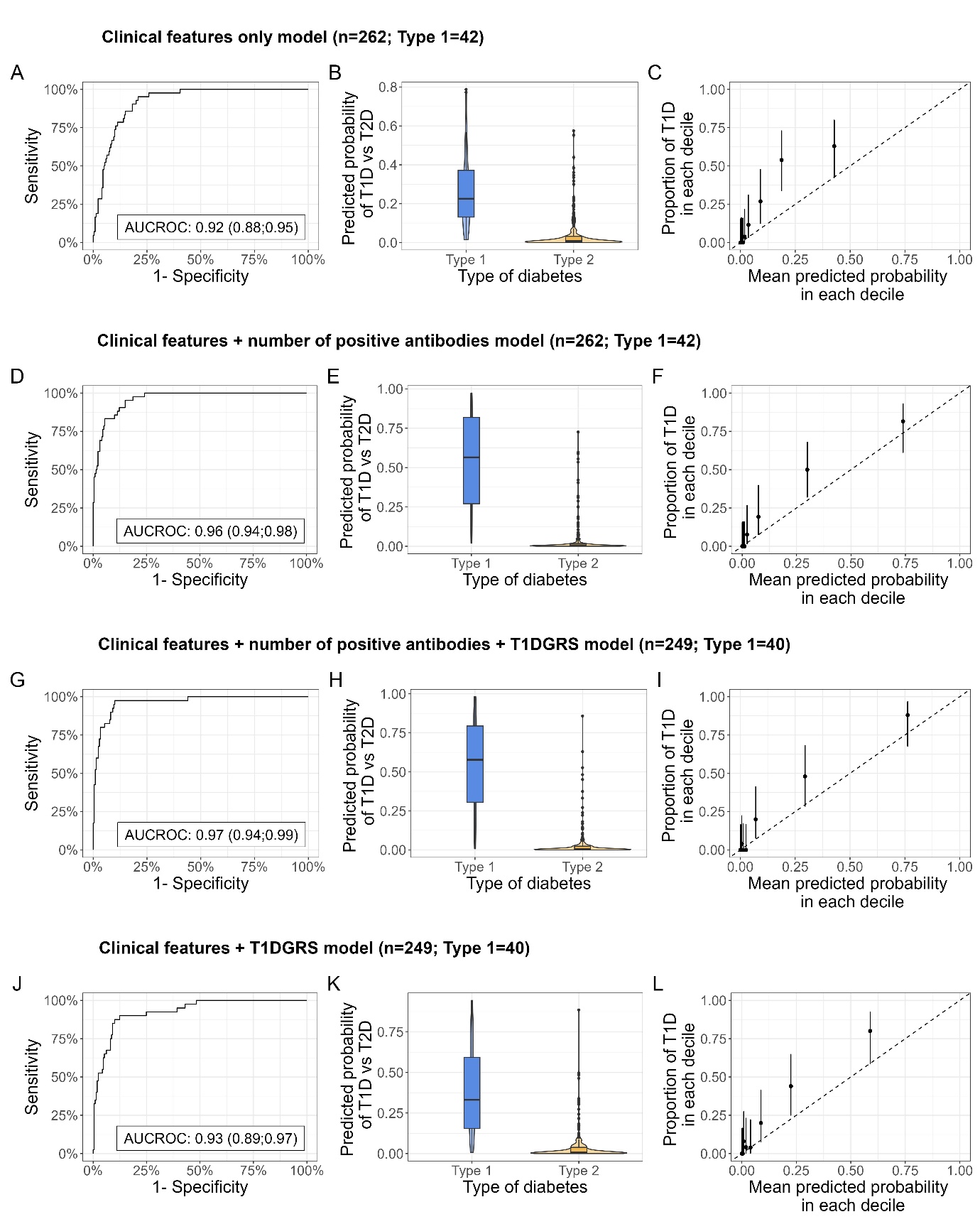

### Appendix Figure 22:

**Model separation and calibration in male individuals**, in StartRight Prime, for clinical features only, clinical features and islet-autoantibodies, clinical features, islet-autoantibodies and T1DGRS, and clinical features and T1DGRS models. Plots A, D, G, and J denote the Receiver Operating Characteristic (ROC) Curve for each respective model in StartRight, with the Area Under the Curve (AUC) shown on the plot (AUCROC (95% CI)). Plots B, E, H, and K represent the predicted probability of the respective model by primary study outcome. Plots C, F, I, and L illustrate the calibration of each model, with deciles of model predicted probabilities of type 1 diabetes plotted against the observed proportion of type 1 diabetes defined by primary study outcome.

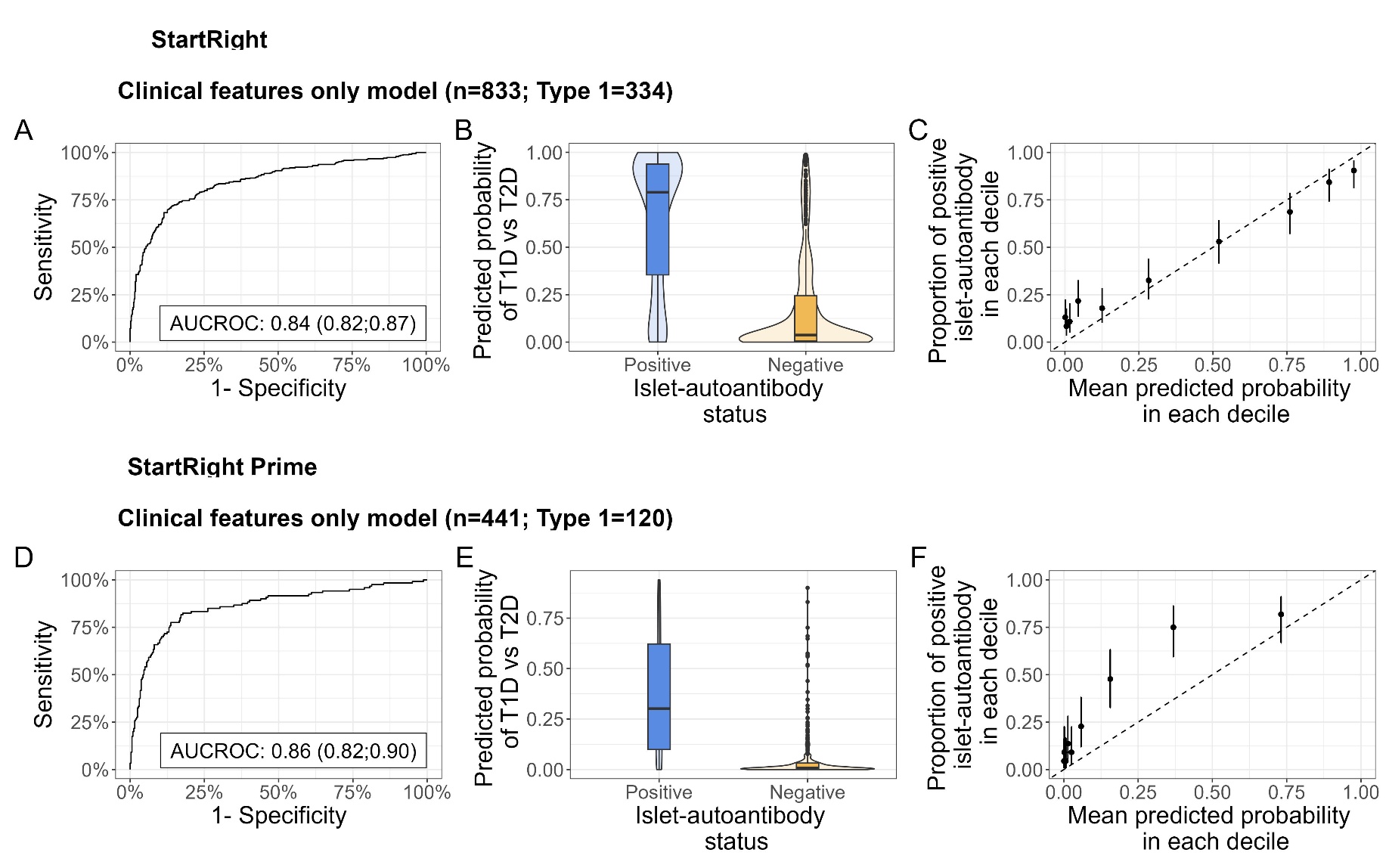

### Appendix Figure 23:

**Model performance of clinical features only model**, **for the prediction of islet-autoantibody positivity** (1+ positive islet-autoantibodies) in StartRight and StartRight Prime. Plots A and D denote the Receiver Operating Characteristic (ROC) Curve for the model in StartRight and StartRight Prime respectively, with the Area Under the Curve (AUC) shown on the plot (AUCROC (95% CI)). Plots B and E represent the predicted probability of the model by islet-autoantibody status. Plots C and F illustrate the calibration of the model, with deciles of model predicted probabilities of type 1 diabetes plotted against the observed proportion of islet-autoantibody positive individuals in each decile.

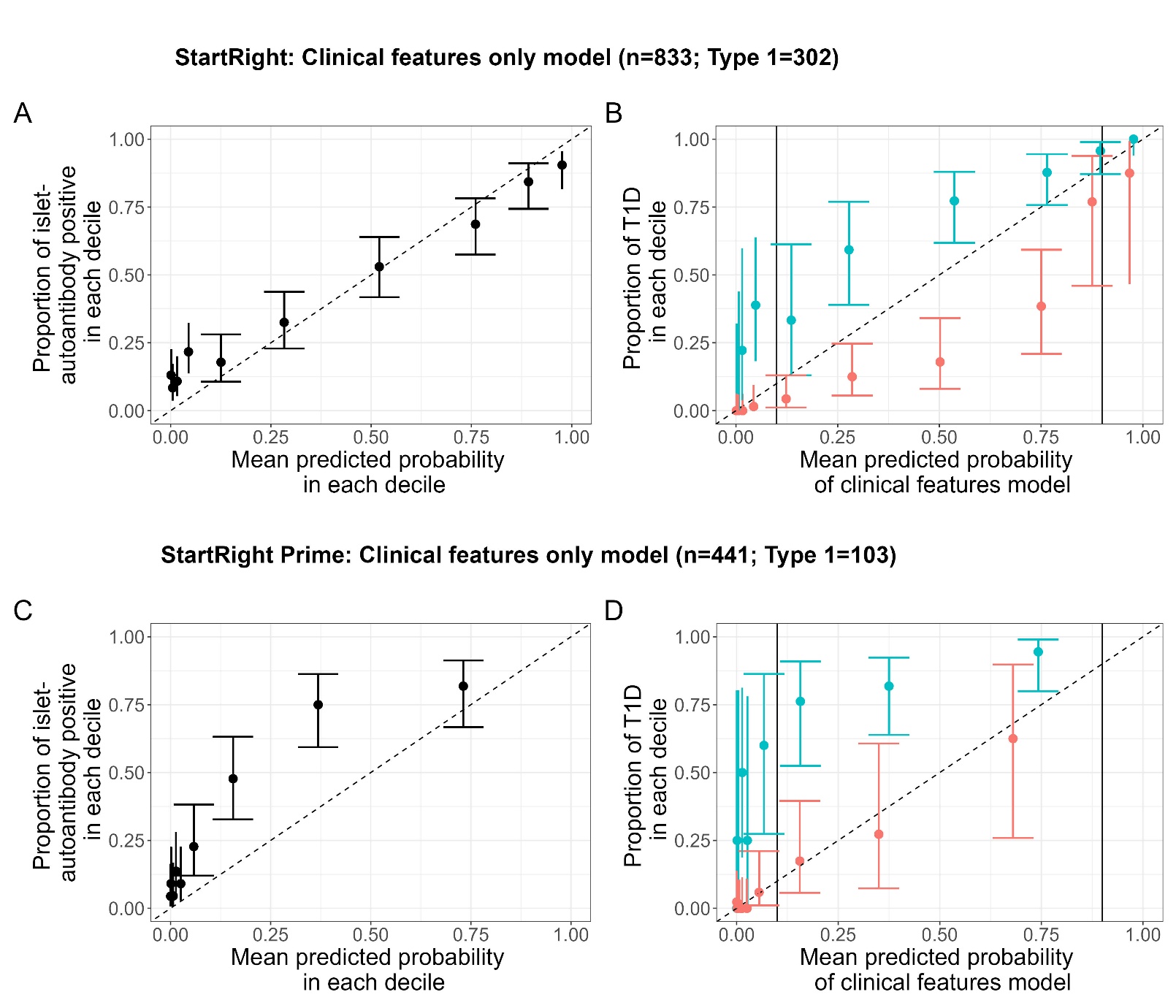

### Appendix Figure 24:

**Clinical features only model can inform when to test islet-autoantibodies**. A and C) Calibration of the clinical features only model when used to predict islet-autoantibody positivity (1+ islet-autoantibodies) in StartRight and StartRight Prime, respectively. Deciles of clinical features only model probabilities of type 1 are plotted against the proportion of islet-autoantibody positive individuals in each decile. B and D) Calibration of the clinical features only model split by islet-autoantibody status (islet-autoantibody positive = blue-turquoise, islet-autoantibody negative = coral-red) in StartRight and StartRight Prime, respectively. Deciles of clinical features only model probabilities of type 1 for each islet-autoantibody status are plotted against the proportion of observed type 1 individuals (defined by 3-year insulin-use and C-peptide) in each decile.

### Appendix Figure 25:

**Separation and calibration of the StartRight Score**, in StartRight and StartRight Prime. Plots A and D denote the Receiver Operating Characteristic (ROC) Curve for the model in StartRight and StartRight Prime respectively, with the Area Under the Curve (AUC) shown on the plot (AUCROC (95% CI)). Plots B and E represent the predicted probability by primary study outcome (diabetes defined by 3-year insulin-use and C-peptide). Plots C and F illustrate the calibration of the model, with deciles of model predicted probabilities of type 1 diabetes plotted against the observed proportion of type 1 diabetes defined by primary study outcome in each score.

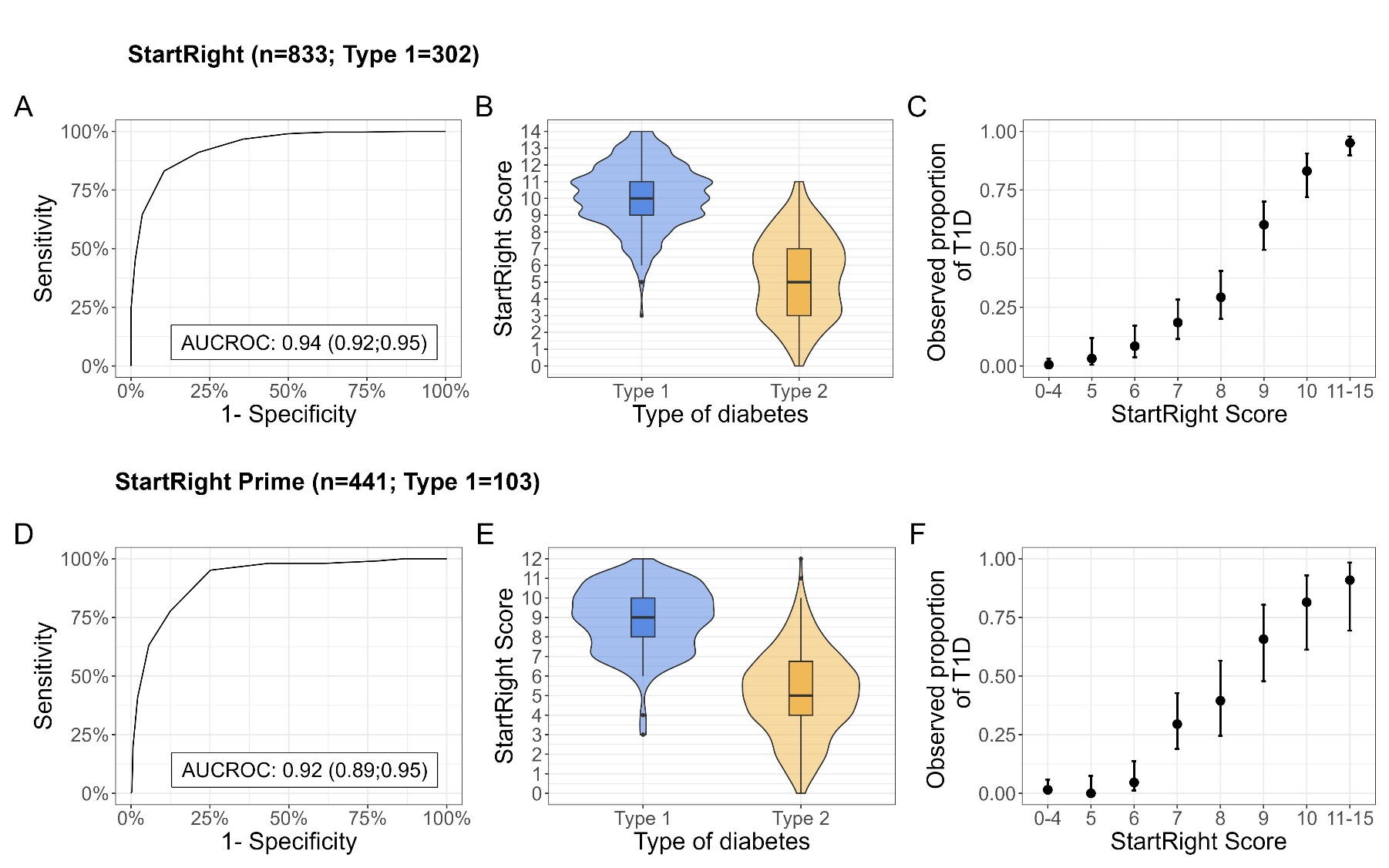

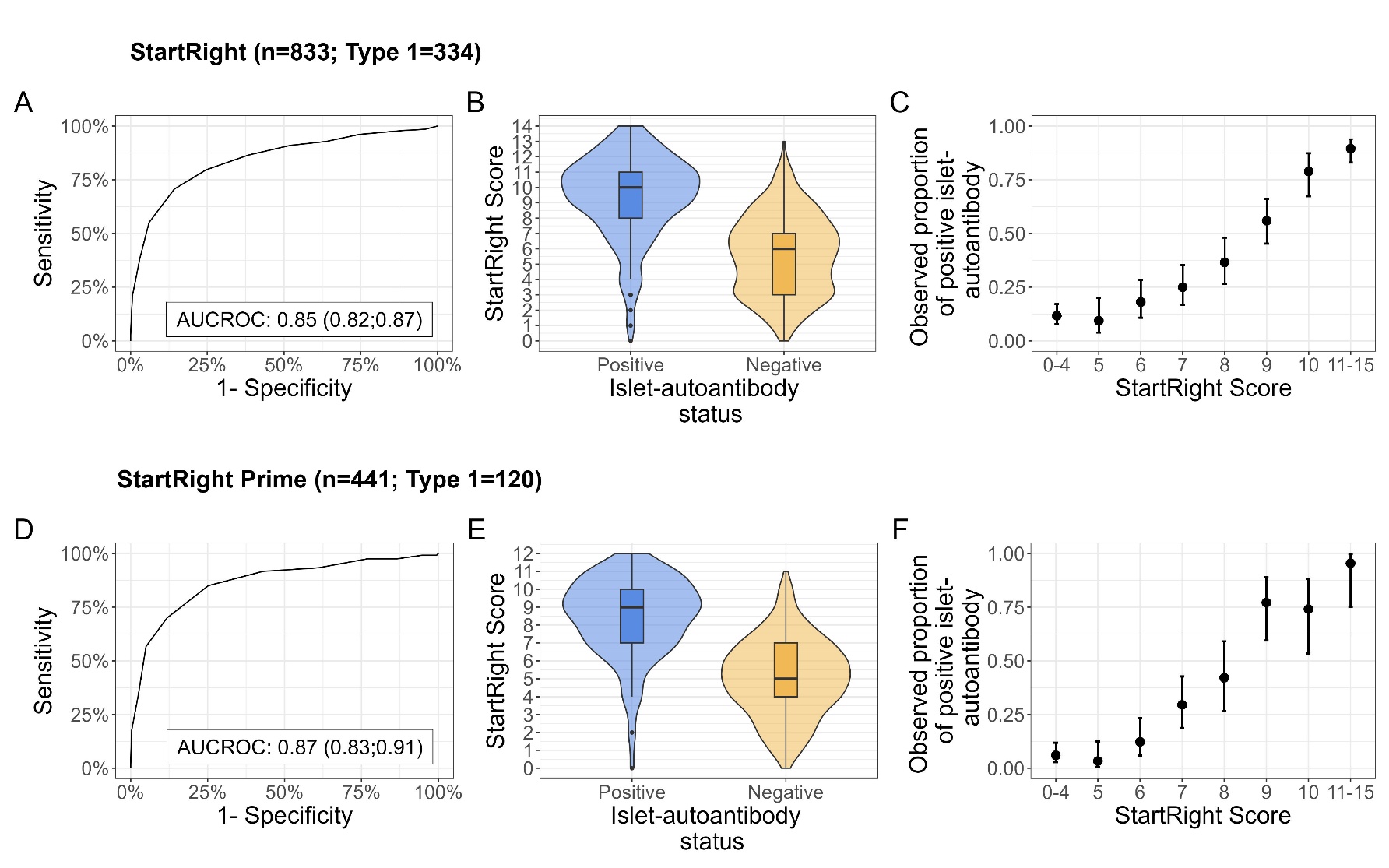

### Appendix Figure 26:

**Predictive performance of StartRight Score**, **for the prediction of islet-autoantibody positivity** (1+ positive islet-autoantibodies) in StartRight and StartRight Prime. Plots A and D denote the Receiver Operating Characteristic (ROC) Curve for the score in StartRight and StartRight Prime respectively, with the Area Under the Curve (AUC) shown on the plot (AUCROC (95% CI)). Plots B and E represent the StartRight Score points distribution by islet-autoantibody status. Plots C and F illustrate the distribution of the score, with StartRight Score points plotted against the observed proportion of islet-autoantibody positive individuals in each point.

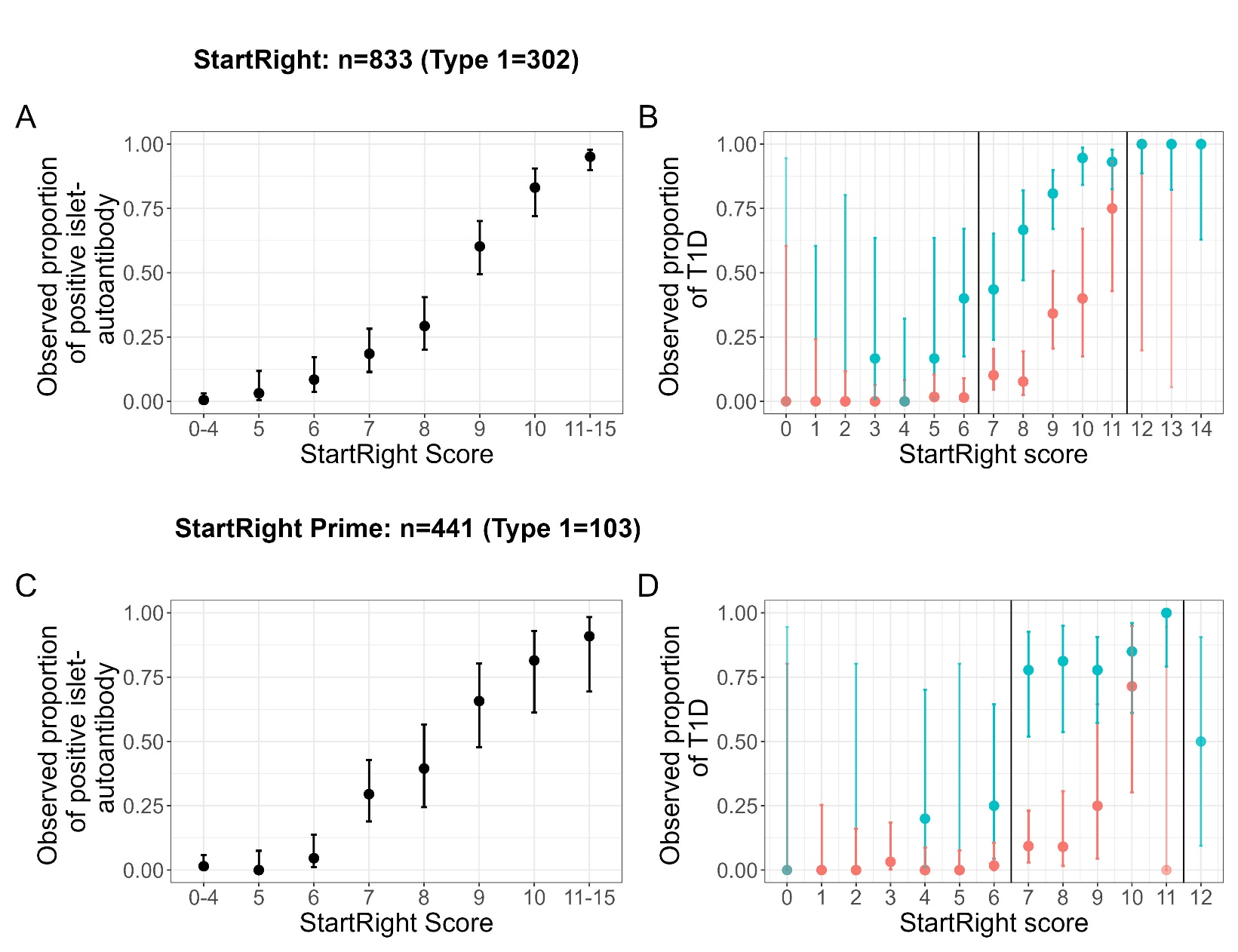

### Appendix Figure 27:

**StartRight Score can inform when to test islet-autoantibodies**. A and C) Distribution of the StartRight Score points when used to predict islet-autoantibody positivity (1+ islet-autoantibodies) in StartRight and StartRight Prime, respectively. StartRight Score points are plotted against the proportion of islet-autoantibody positive individuals in each point. B and D) Distribution of the StartRight Score points split by islet-autoantibody status (islet-autoantibody positive = blue-turquoise, islet-autoantibody negative = coral-red) in StartRight and StartRight Prime, respectively. StartRight Score points for each islet-autoantibody status are plotted against the proportion of observed type 1 individuals (defined by 3-year insulin-use and C-peptide) in each point.

### Appendix Table 15:

**Scoring table for use at diagnosis for the classification of type 1 diabetes versus type 2 diabetes.** Scores derived from rounding beta coefficients of clinical features and islet-autoantibodies model to nearest integer. *Black, South Asian, Other or Mixed ethnicity

| **Type of diabetes at diagnosis Scoring Table** | **StartRight Islet-autoantibodies Score** |
| --- | --- |
| Age at diagnosis (years): 18-34 | 2 |
| 35-44 years | 1 |
| ≥45 years | 0 |
| BMI at diagnosis (kg/m^2^): <25 | 4 |
| 25-34 kg/m^2^ | 2 |
| ≥35 kg/m^2^ | 0 |
| HbA1c at diagnosis (mmol/mol): <58 | 0 |
| ≥58 mmol/mol | 1 |
| Female sex | 1 |
| Presentation osmotic symptoms | 1 |
| Presence of additional autoimmune disease | 1 |
| Presentation unintentional weight-loss | 1 |
| Presentation Ketoacidosis | 1 |
| No parent history of non-insulin-treated diabetes | 1 |
| Ethnicity associated with high type 2 diabetes risk* | -1 |
| Islet-autoantibodies (of GAD, IA2 & ZNT8 testing): all negative | 0 |
| One positive | 2 |
| Two positive | 3 |
| Three positive | 4 |
|  | **17** |

### Appendix Figure 28:

**Separation and calibration of the StartRight Score with islet-autoantibodies**, in StartRight and StartRight Prime. Plots A and D denote the Receiver Operating Characteristic (ROC) Curve for the model in StartRight and StartRight Prime respectively, with the Area Under the Curve (AUC) shown on the plot (AUCROC (95% CI)). Plots B and E represent the predicted probability by primary study outcome (diabetes defined by 3-year insulin-use and C-peptide). Plots C and F illustrate the calibration of the model, with deciles of model predicted probabilities of type 1 diabetes plotted against the observed proportion of type 1 diabetes defined by primary study outcome in each score.

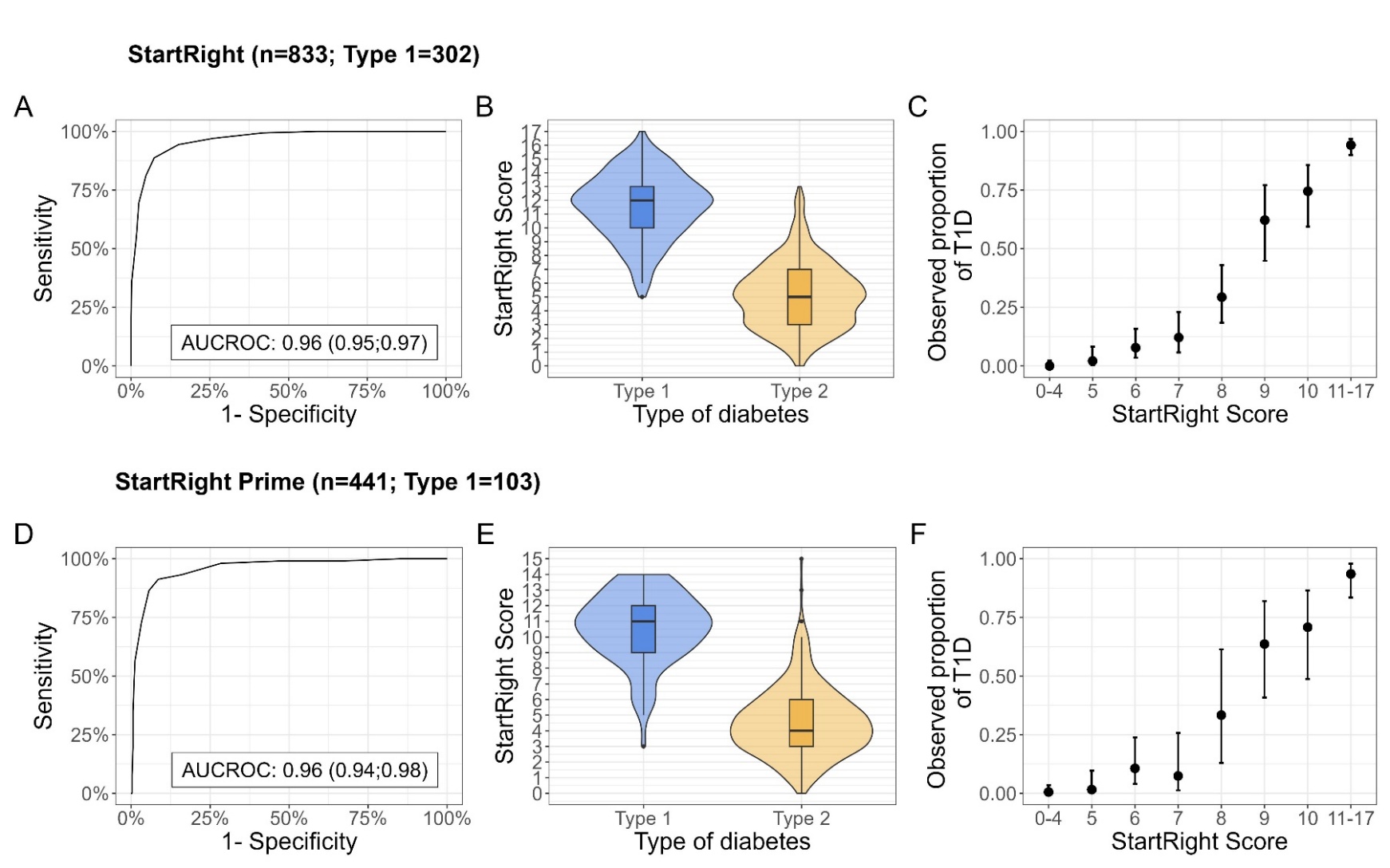

### Appendix Table 16:

CPRD characteristics table. Numerical characteristics are described by median [Interquartile range], and categorical characteristics by n (percentage). Note that (for robust assessment of model performance for early insulin requirement) analysis of those initially treated as type 2 diabetes/without insulin was restricted to participants with ≥3 years follow up duration.

| **Characteristic** | | **Initially treated as type 2 diabetes cohort** | **Unselected diabetes subtype cohort** |
| --- | --- | --- | --- |
| N |  | 84,194 | 188,232 |
| Ethnicity | White | 65,821 (78·2%) | 147,190 (78.2%) |
|  | Black | 49,91 (5·9%) | 11,106 (5.9%) |
|  | South Asian | 10,659 (12·7%) | 23,461 (12.5%) |
|  | Other/Mixed | 2,723 (3·2%) | 6,475 (3.4%) |
| Sex | Female | 35,927 (42·7%) | 78,836 (41.9%) |
|  | Male | 48,267 (57·3%) | 109,396 (58.1%) |
| Age at diagnosis (years) | | 58.7 [50.1;68.7] | 58·81 [49·8;69·0] |
| BMI (kg/m^2^) at diagnosis* | | 32.0 [28.1;36.9.0] | 32·0 [28·0;37·0] |
| HbA1c at diagnosis (mmol/mol) | | 51.0 [48.8;60.0] | 52·0 [49·0;63·0] |
| Follow-up duration (days) | | 1,680 [1,346;1,965] | 985 [512;1,614] |
| Parent history of non-insulin-treated diabetes | | 2,6711 (31.7%) | 60,089 (31·9%) |
| DKA | | 98 (0.1%) | 600 (0·3%) |
| Unintentional weight-loss | | 605 (0.7%) | 1702 (0·9%) |
| Presence of osmotic symptoms | | 8,307 (9.9%) | 17,872 (9·5%) |
| Presence of additional autoimmune disorder/s | | 1,447 (1.7%) | 3,657 (1·9%) |

### Appendix Figure 29:

**Distribution of insulin treatment within three years (A), subsequent ketoacidosis (B), a subsequent diagnostic code for type 1 diabetes (C), and subsequent basal bolus insulin (D) by clinical features only model probability in adults newly diagnosed with diabetes, from UK primary care records** (CPRD: n=188,232). Error bars denote logit-transformed 95% confidence intervals around the proportion.

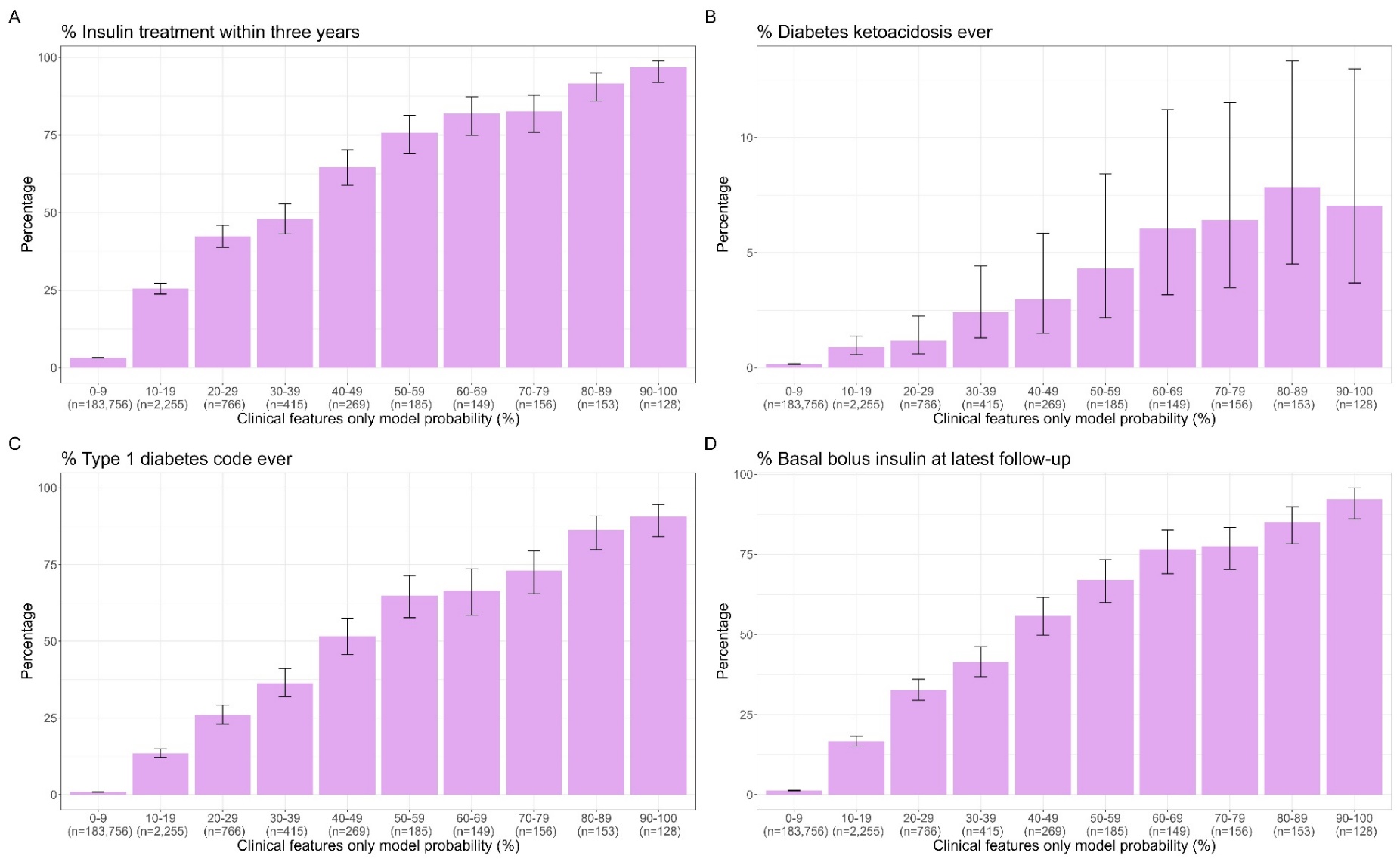

### Appendix Figure 30:

**Distribution of insulin treatment within three years (A), subsequent ketoacidosis (B), a subsequent diagnostic code for type 1 diabetes (C), and subsequent basal bolus insulin (D) by StartRight Score points in adults newly diagnosed with diabetes** from UK primary care records (CPRD: n=188,232). Error bars denote logit-transformed 95% confidence intervals around the proportion.

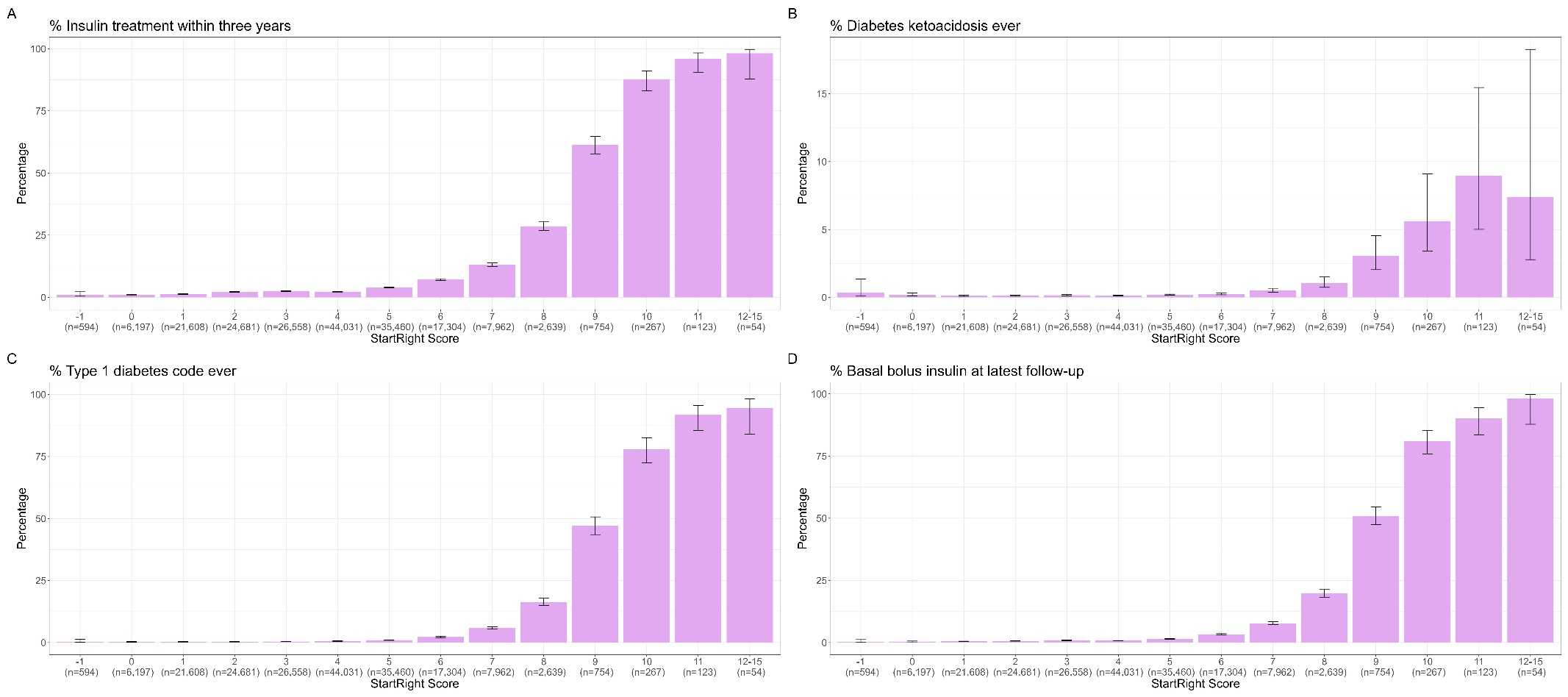

### Appendix Figure 31:

**Model separation and calibration of the clinical features only model in CPRD, predicting progression to insulin in three years in individuals not on insulin at diagnosis**. Plot A denotes the Receiver Operating Characteristic (ROC) Curve for the model, with the Area Under the Curve (AUC) shown on the plot. Panel B represents the predicted probability of the model by whether individuals progressed to insulin in 3 years or not. Panel C illustrates the calibration of the model, with deciles of model predicted probabilities of type 1 diabetes plotted against the observed proportion of individuals that progressed to insulin in 3 years.

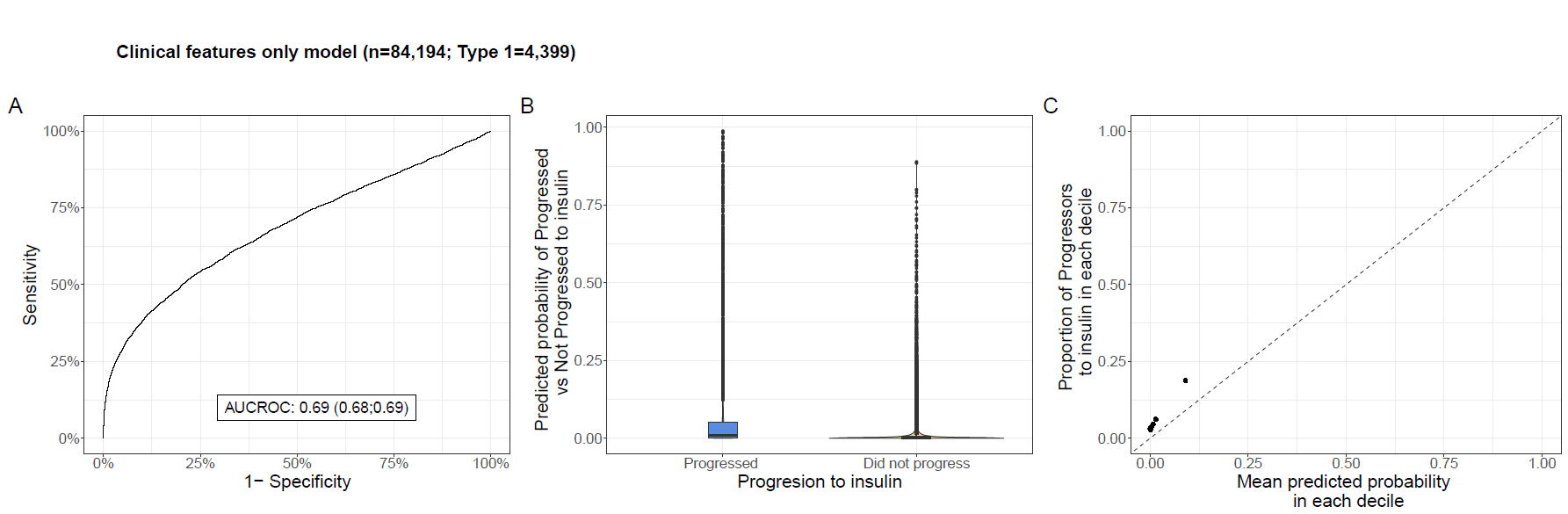

### Appendix Figure 32:

Distribution of early progression to insulin (A), subsequent ketoacidosis (B), a subsequent diagnostic code for type 1 diabetes (C), and subsequent basal bolus insulin (D) by StartRight Score points in adults newly diagnosed with diabetes, initially treated without insulin (for ≥4 weeks), from UK primary care records (CPRD: n=84,692). Error bars denote logit-transformed 95% confidence intervals around the proportion. N’s below bars denote the total number of participants within that probability range (denominator of percentage).

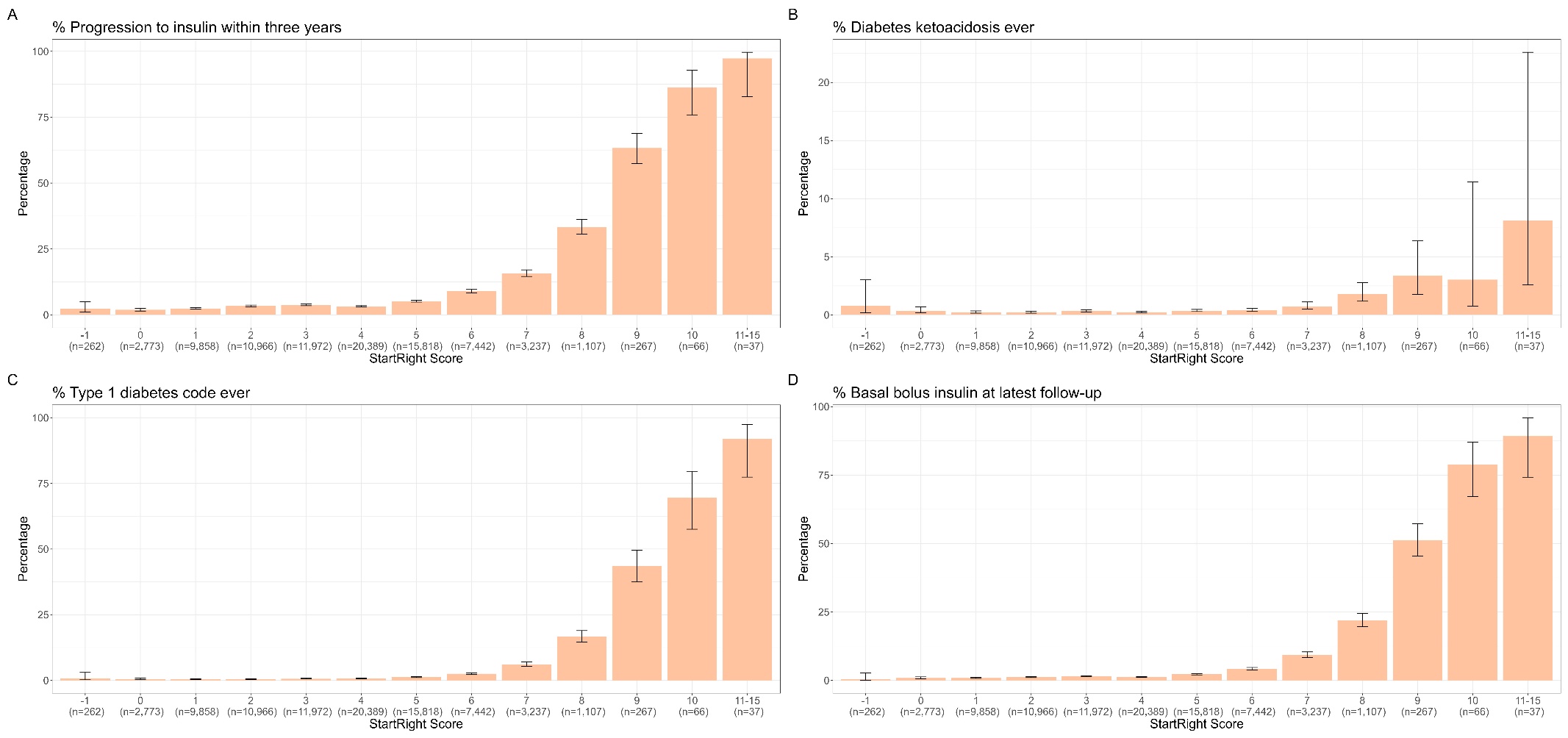

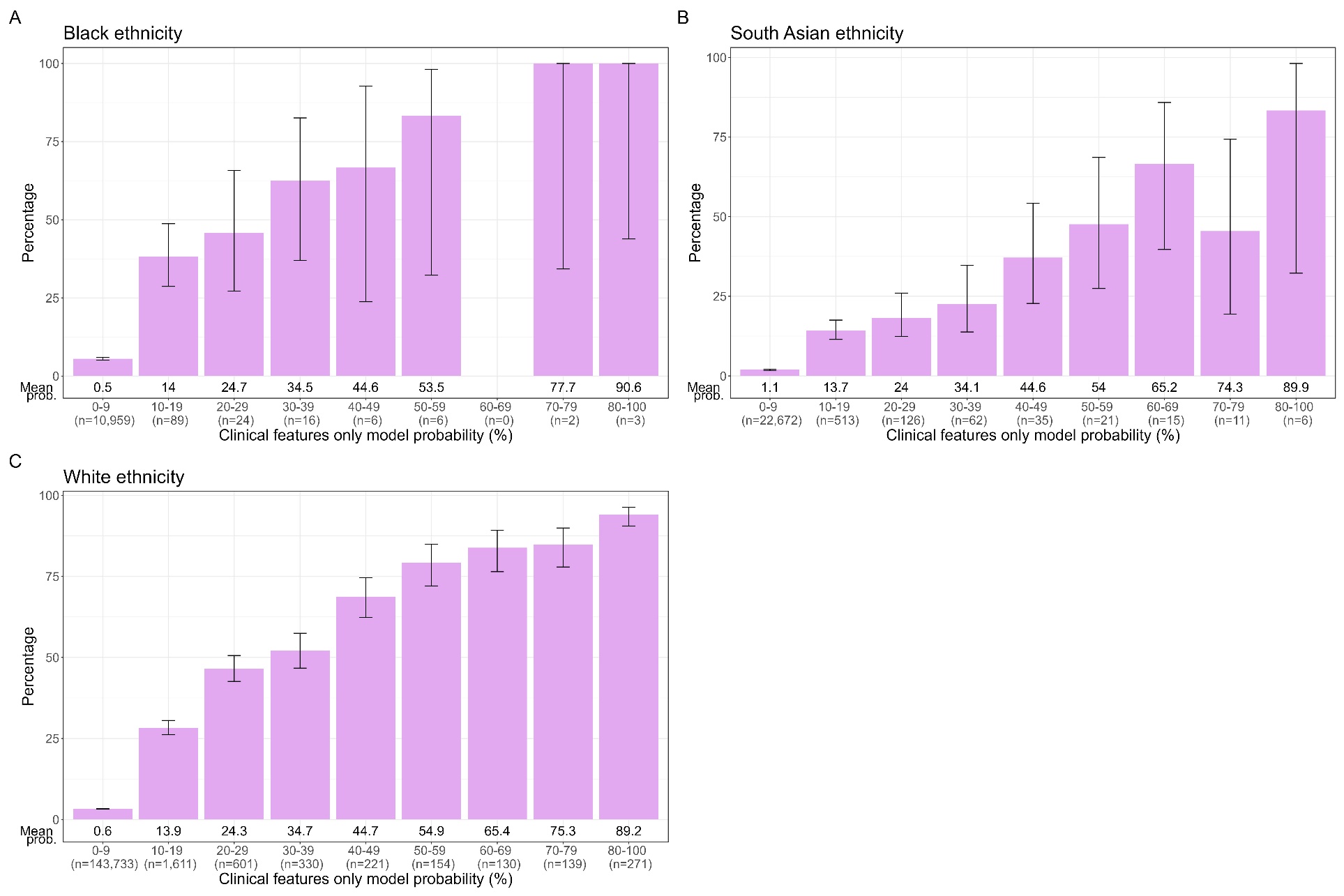

### Appendix Figure 33:

**Distribution of insulin treatment within three years of diagnosis by clinical features only model probability in adults newly diagnosed with diabetes from UK primary care records** (CPRD: n=188,232**),** in individuals of (A) black, (B) south Asian, and (C) white ethnicity. Error bars denote logit-transformed 95% confidence intervals around the proportion; when proportion is 100%, Wilson method is used.

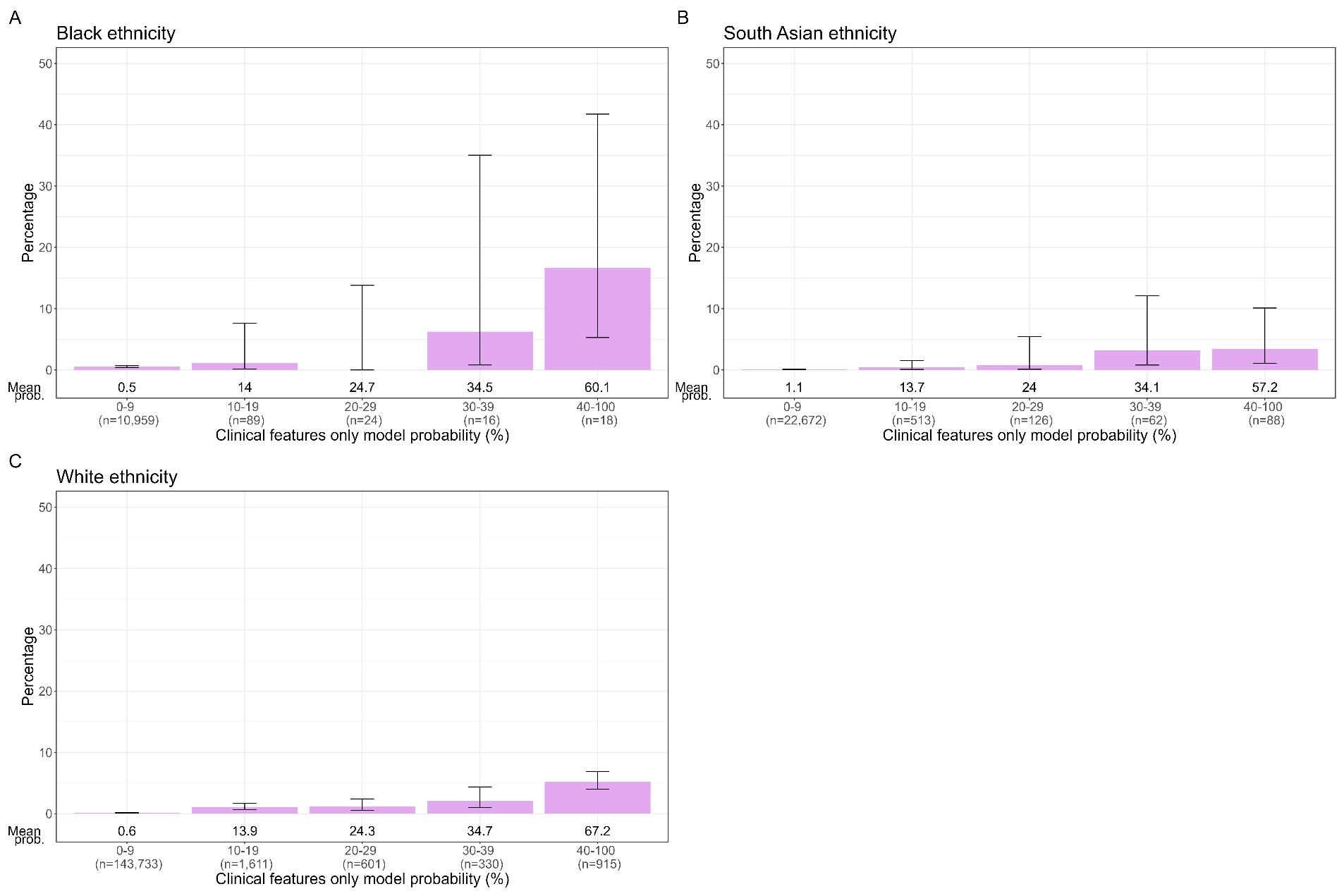

### Appendix Figure 34:

**Distribution of subsequent ketoacidosis by clinical features only model probability in adults newly diagnosed with diabetes from UK primary care records** (CPRD: n=188,232), in individuals of (A) black, (B) south Asian, and (C) white ethnicity. Error bars denote logit-transformed 95% confidence intervals around the proportion; when proportion is 0%, Wilson method is used.

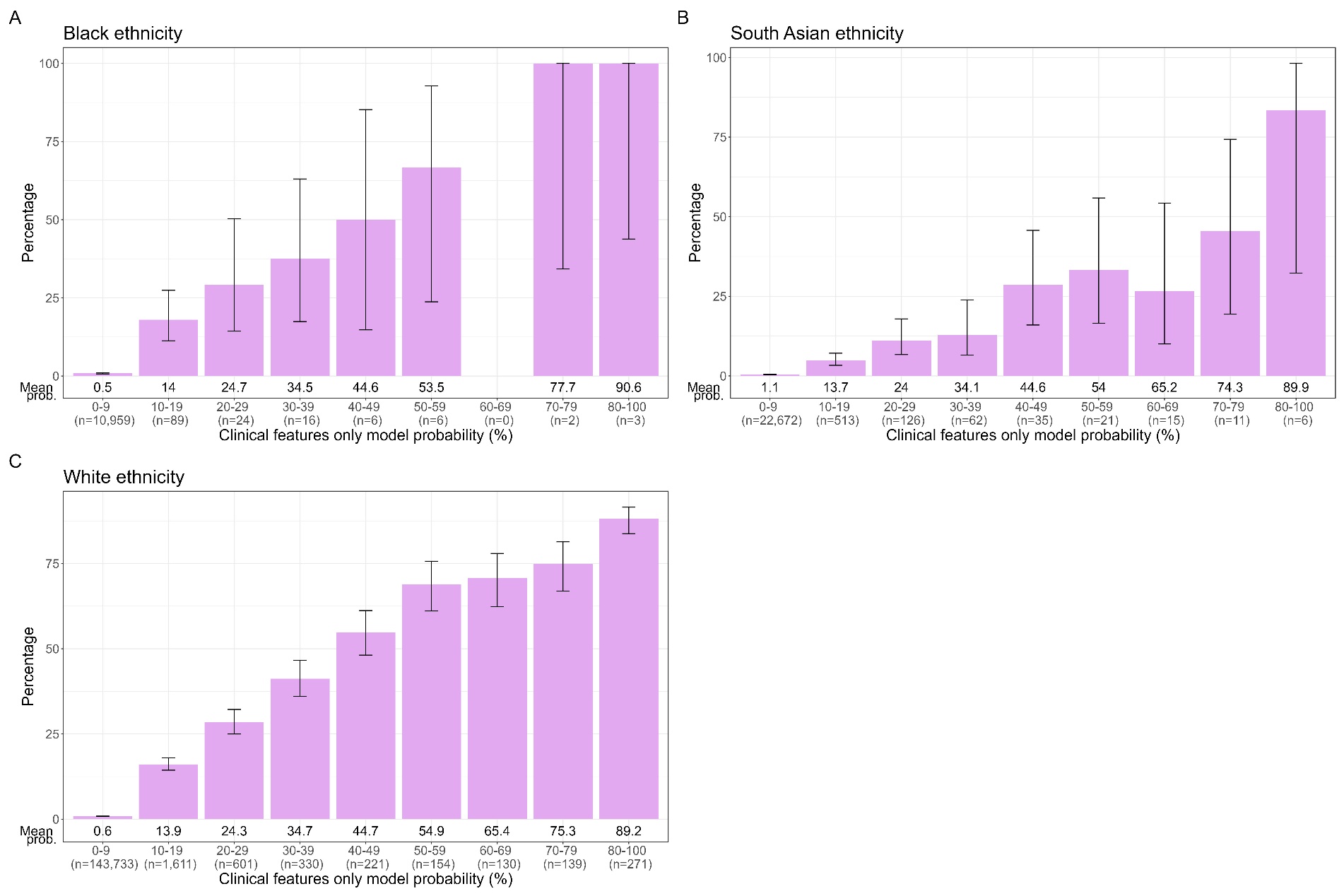

### Appendix Figure 35:

**Distribution of any subsequent type 1 diabetes code by clinical features only model probability in adults newly diagnosed with diabetes from UK primary care records** (CPRD: n=188,232), in individuals of (A) black, (B) south Asian, and (C) white ethnicity. Error bars denote logit-transformed 95% confidence intervals around the proportion; when proportion is 100%, Wilson method is used.

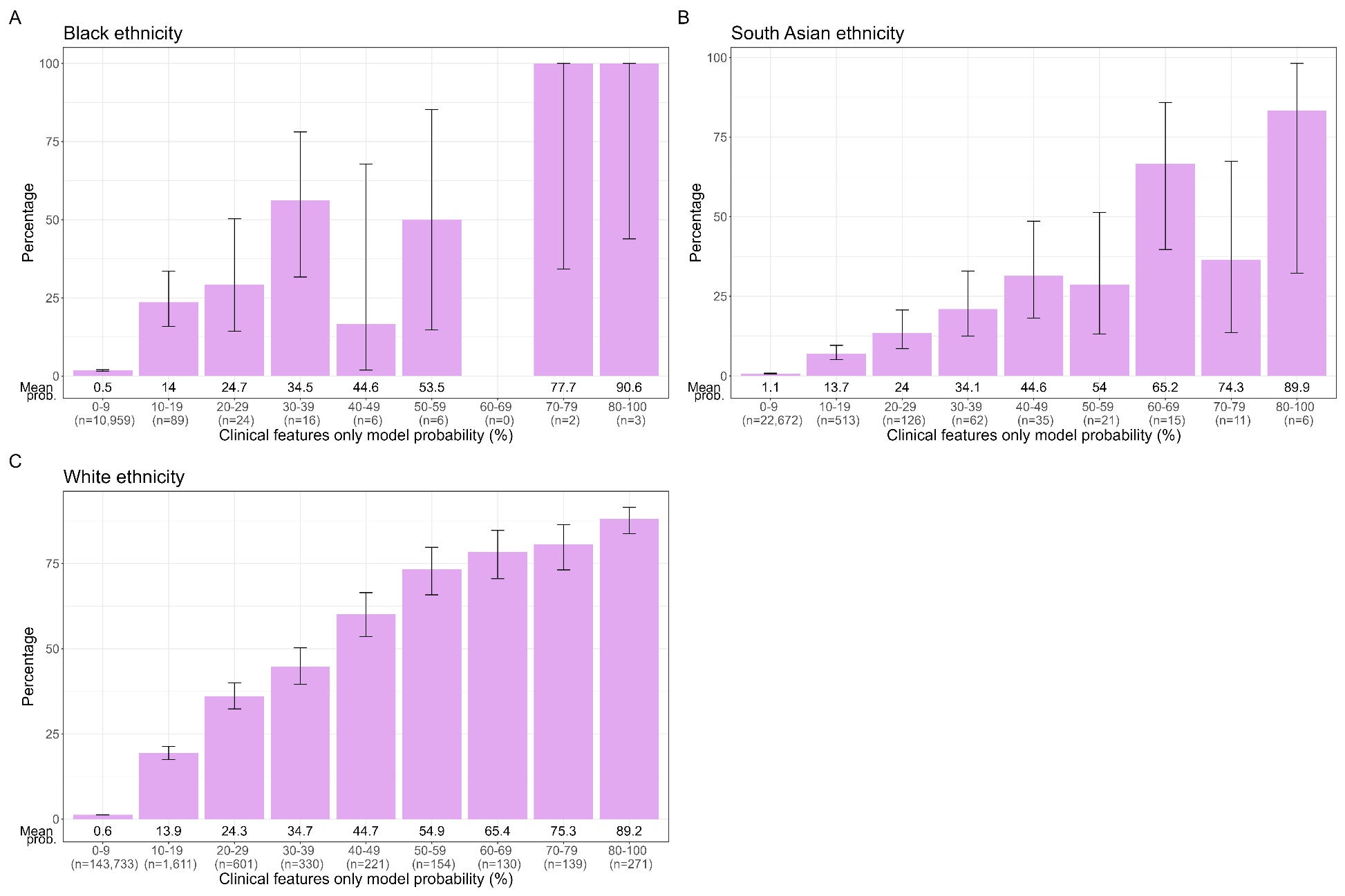

### Appendix Figure 36:

**Distribution of subsequent basal bolus insulin treatment at latest follow-up by clinical features only model probability in adults newly diagnosed with diabetes from UK primary care records** (CPRD: n=188,232), in individuals of (A) black, (B) south Asian, and (C) white ethnicity. Error bars denote logit-transformed 95% confidence intervals around the proportion; when proportion is 100%, Wilson method is used.

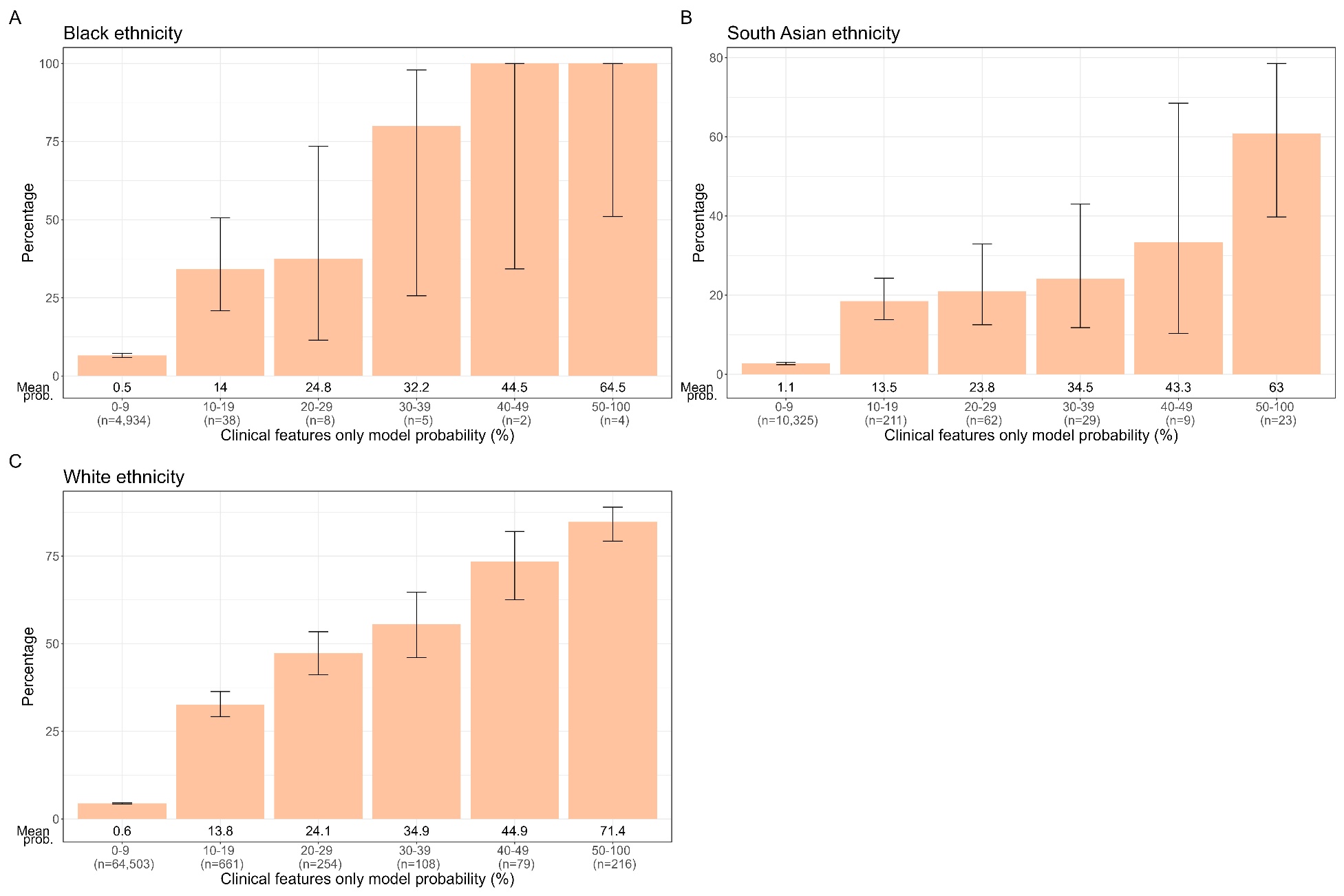

### Appendix Figure 37:

**Distribution of early progression to insulin by clinical features only model probability in adults newly diagnosed with diabetes, initially treated without insulin** (for ≥4 weeks), from UK primary care records (CPRD: n=84,194), in individuals of (A) black, (B) south Asian, and (C) white ethnicity. Error bars denote logit-transformed 95% confidence intervals around the proportion; when proportion is 100%, Wilson method is used.

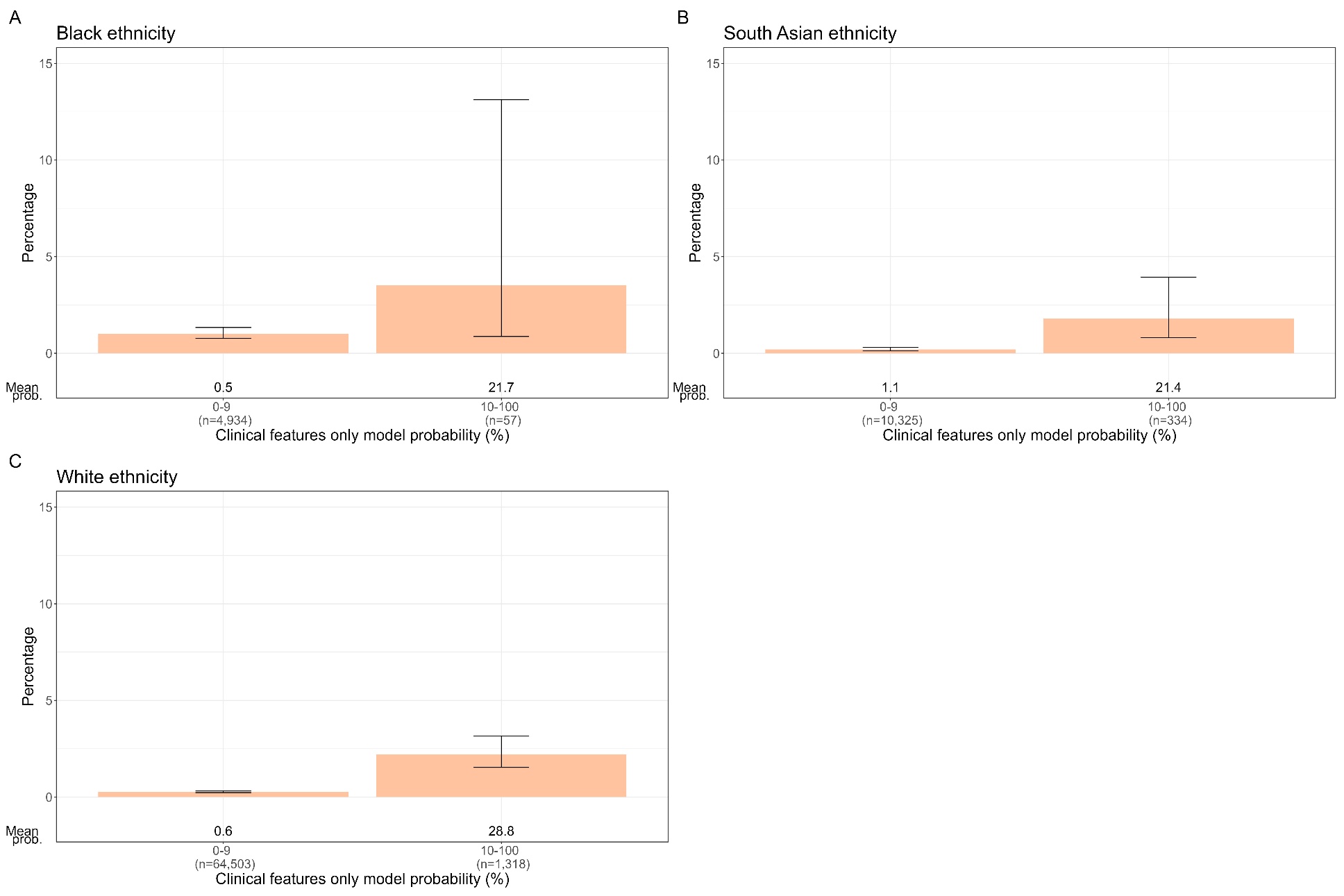

### Appendix Figure 38:

**Distribution of subsequent ketoacidosis by clinical features only model probability in adults newly diagnosed with diabetes, initially treated without insulin** (for ≥4 weeks), from UK primary care records (CPRD: n=84,194), in individuals of (A) black, (B) south Asian, and (C) white ethnicity. Error bars denote logit-transformed 95% confidence intervals around the proportion.

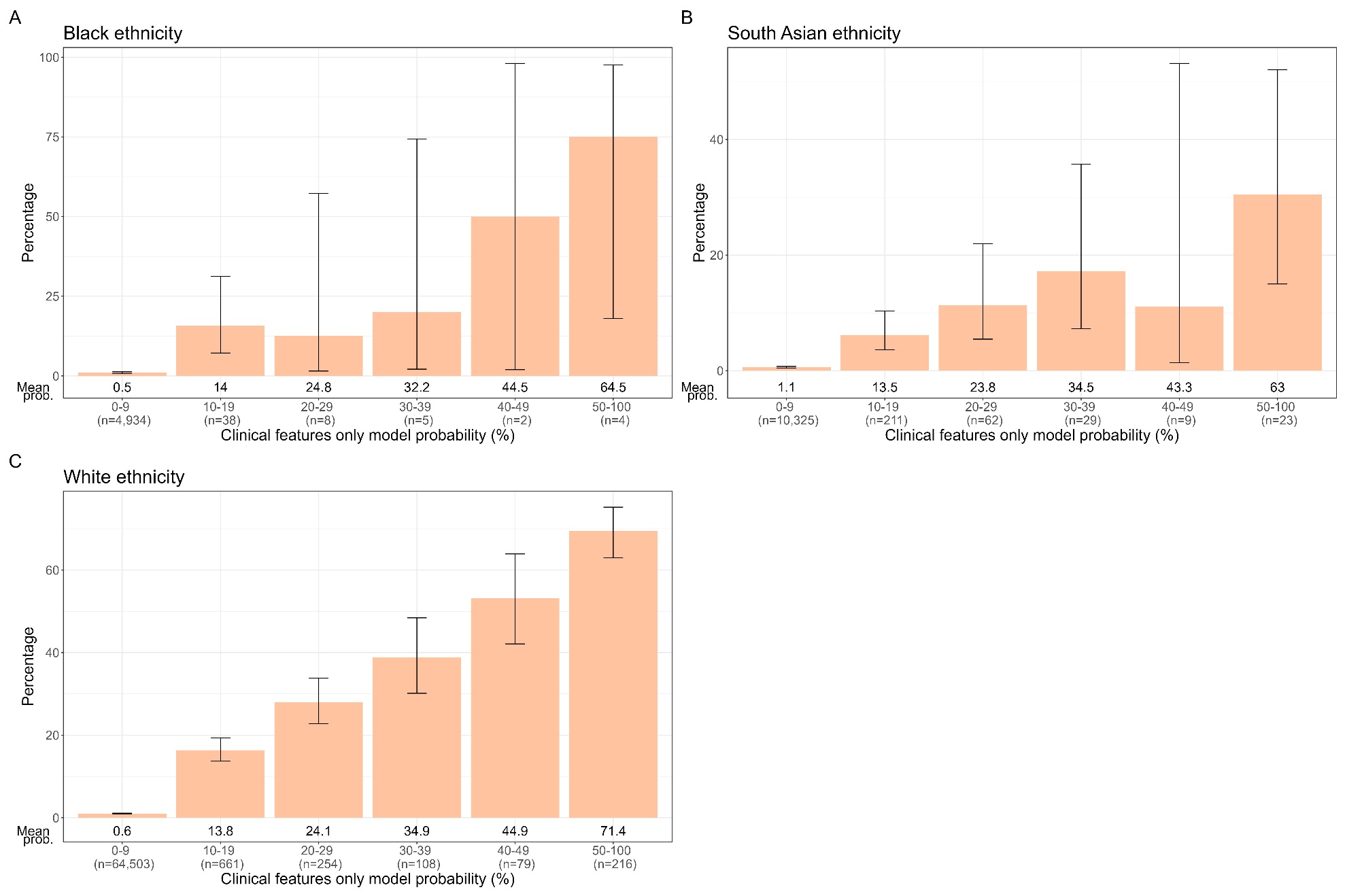

### Appendix Figure 39:

**Distribution of subsequent type 1 diabetes code by clinical features only model probability in adults newly diagnosed with diabetes, initially treated without insulin** (for ≥4 weeks), from UK primary care records (CPRD: n=84,194), in individuals of (A) black, (B) south Asian, and (C) white ethnicity. Error bars denote logit-transformed 95% confidence intervals around the proportion.

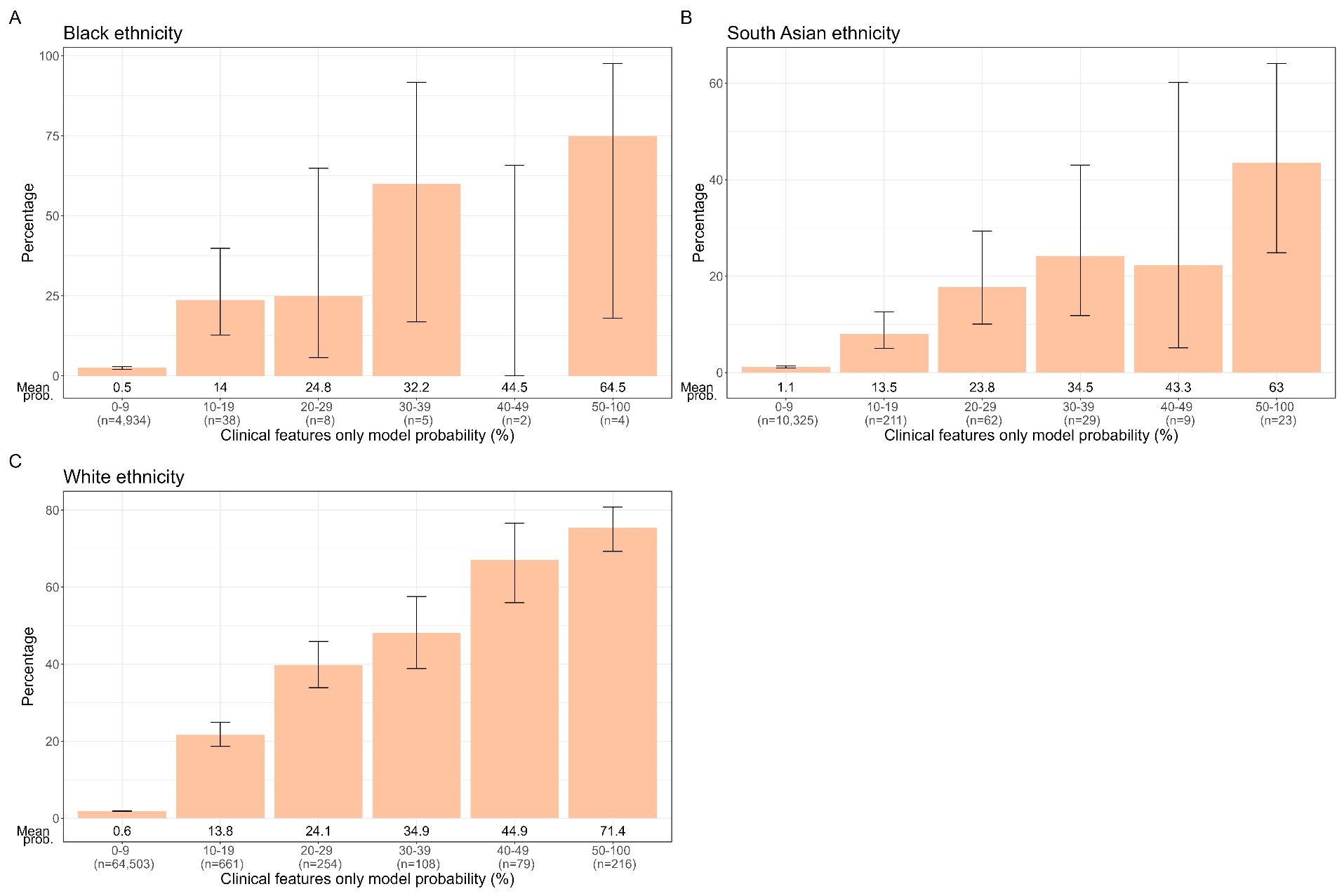

### Appendix Figure 40:

**Distribution of subsequent basal bolus insulin at latest follow up by clinical features only model probability in adults newly diagnosed with diabetes, initially treated without insulin** (for ≥4 weeks), from UK primary care records (CPRD: n=84,194) in individuals of (A) black, (B) south Asian, and (C) white ethnicity. Error bars denote logit-transformed 95% confidence intervals around the proportion; when proportion is 0%, Wilson method is used.

### Appendix Table 17:

**Approaches in identifying individuals to test islet-autoantibodies, and performance of combined approaches in identifying type 1 diabetes (T1D)**. †CPRD = 188,232 participants with diabetes diagnosis over the age of 18 years (population cohort) *Presence of either: DKA, unintentional weight loss, age-at-diagnosis <50, BMI <25, and presence of either personal or family history of an autoimmune disease. ‡EASD/ADA: Presence of either: DKA, unintentional weight-loss, age-at-diagnosis <30, BMI <25, and presence of either personal or family history of an autoimmune disease. **Family history of autoimmune disorder not available in CPRD. ***Type 1 defined as clinical features model ≥10% and clinical features + islet-autoantibodies model ≥50%; Type 2 diabetes defined as either clinical features only model <10% or clinical features +islet-autoantibodies model <50%. **** Type 1 defined as StartRight Score ≥7 and StartRight Score with islet-autoantibodies ≥8; Type 2 diabetes defined as either StartRight Score < 7 or StartRight Score with islet-autoantibodies <8

|  | **CPRD†** | **Combined StartRight and StartRight Prime (n=1,274; type 1 = 405)** | |
| --- | --- | --- | --- |
|  | **% eligible for islet-autoantibody testing** | **Sensitivity for T1D; % (95% CI) [n]** | **Specificity for T1D; % (95% CI) [n]** |
| Test islet-autoantibodies in everyone | 100 [188,232/188,232] | 85·2 (81·4; 88·3) [345/405] | 87·5 (85·1; 89·5) [760/869] |
| Test everyone with NICE criteria for T1D* | 40·9 [76,910/188,232] ** | 84·2 (80·3; 87·4) [341/405] | 88·7 (86·4; 90·7) [771/869] |
| Test everyone with ADA/EASD criteria for T1D‡ | 15·4 [29,044/188,232] ** | 82·7 (78·7; 86·1) [335/405] | 92·1 (90·1; 93·7) [800/869] |
| Test everyone with a clinical features only model probability ≥10% | 2·4 [4,476/188,232] | 81·0 (76·9; 84·5) [328/405] *** | 95·6 (94; 96·8) [831/869] *** |
| Test everyone with a StartRight Score ≥7 points | 6·3 [11,799/188,232] | 86·4 (82·7; 89·4) [350/405] **** | 93·8 (92; 95·2) [815/869] **** |

### Appendix: StartRight Consortium

**Recruiting Centres:** Basildon & Thurrock University Hospital NHS Foundation Trust, Dr Godwin Simon & Angelo Ramos RN (PIs); Hampshire Hospitals NHS Foundation Trust, Dr Andrea Norris (PI); Royal United Hospitals Bath NHS Foundation Trust, Dr Kai Tan(PI); University Hospitals Birmingham NHS Foundation Trust, Dr Parth Narendran(PI); East Lancashire Hospitals NHS Trust, Dr Shenaz Ramtoola(PI); Oakenhurst Medical Practice, Dr Amar Ali(PI); Bolton NHS Foundation Trust, Dr Moulinath Banerjee(PI); University Hospitals Dorset NHS Foundation Trust, Dr Augustin Brooks(PI); Brighton & Sussex University Hospital NHS Trust, Dr Ali Chakera(PI); North Bristol NHS Trust, Dr Andrew Johnson & Dr Danijela Tatovic (PIs); Buckinghamshire Healthcare NHS Trust, Dr Chitrabhanu Ballav (PI); Cardiff and Vale University Health Board - University Hospital of Wales, Prof Colin Dayan (PI); Countess of Chester Hospital NHS Foundation Trust, Dr Sunil Nair (PI); Derby Teaching Hospitals NHS Foundation Trust, Prof Francis Game(PI); Gloucestershire Hospitals NHS Foundation Trust, Susan Beames RN (PI); East Suffolk & North Essex NHS Foundation Trust, Prof Gerry Rayman (PI); United Lincolnshire Hospitals NHS Trust, Marie Snell RN, Susie Butler RN & Sarah Beck RN (PIs); Lincoln Community Health Services NHS Trust, Janet Beecham RN (PI); Liverpoot University Hospitals NHS Foundation Trust, Prof John Wilding (PI); Hywel Dda University Health Board, Dr Sam Rice (PI); St Georges University Hospital NHS Foundation Trust, Dr Mimi Chen (PI); Western Health & Social Care Trust, Dr Athinyaa Thiraviaraj (PI); Maindstone & Tunbridge Wells NHS Trust, Dr Siva Sivappriyan(PI); Manchester University NHS Foundation Trust, Dr Basil Issa(PI); Milton Keynes University Hospital NHS Foundation Trust, Dr Asif Humayun and Rebecca Hinch RN (PIs); James Paget University Hospital NHS Foundation Trust, Dr Leena Krishnan & Dr Khin Swe Myint (PIs); Northampton General Hospital NHS Trust, Dr Charles Fox & Dr Jennifer Prouten (PIs); Norfolk & Norwich University Hospital NHS Foundation Trust, Prof Mike Sampson (PI); Nottingham University Hospital NHS Trust, Dr Peter Mansell & Dr Carolyn Chee (PIs); Oxford University Hospitals NHS Foundation Trust, Dr Katherine Owen (PI); Plymouth Hospitals NHS Trust, Dr Ioannis Dimitropoulis (PI); Portsmouth Hospitals NHS Trust, Prof Michael Cummings (PI); Royal Berkshire NHS Foundation Trust, Dr Foteini Kavourra (PI); Salford Royal Hospital, Dr Adrian Heald (PI); Sheffield Teaching Hospitals NHS Foundation Trust, Prof Simon Heller (PI); Sherwood Forest Hospitals NHS Foundation Trust, Dr Sarbpreet Sihota & Dr Vakkat Muraleedharan (PIs); The Solent NHS Trust, Dr Tara Watson (PI); Southern Health NHS Foundation Trust, Dr Hermione Price (PI); Midlands Partnership NHS Foundation Trust, Mr Roger Whittaker & Sarah Orme RN (PIs); Surrey and Sussex Heathcare NHS Trust, Dr Ben Field (PI); Swansea Bay University Health Board, Prof Stephen Bain (PI); Great Western Hospitals NHS Foundation Trust, Dr Beas Battacharya Lesley Haxton & Suzannah Pegler (PIs); Somerset NHS Foundation Trust, Catherine Thompson RN & Prof Rob Andrew (PIs); Torbay and South Devon NHS Foundation Trust, Dr Jamie Smith (PI); Royal Cornwall Hospitals NHS Trust, Dr Duncan Browne & Dr Steve Creely (PIs); Warrington & Halton Hospitals NHS Foundation Trust, Dr Rahul Yadav (PI); South Warwickshire NHS Foundation Trust, Dr Rakhi Kakad (PI); Western Sussex Hospitals NHS Foundation Trust, Dr Ken Laji(PI); Wrightington,Wigan and Leigh Teaching Hospitals NHS Foundation Trust, Dr Mohit Kumar(PI); Berkshire Healthcare NHS Foundation Trust, Dr Alirezi Mohammadi (PI); The Royal Wolverhampton NHS Trust, Dr James Young (PI); Yeovil District Hospital NHS Foundation Trust, Dr Seshadri Pramodh (PI); York & Scarborough Teaching Hospitals NHS Foundation Trust, Dr Vijay Jayagopal (PI).

**Lead Site & Coordinating Centre:** NIHR Exeter Clinical Research Facility, Royal Devon & Exeter Hospital: Dr Angus Jones (CI), Anita Hill, Robert Bolt, Suzanne Hammersley, Migaila Aldred, Anna Steele, Peter Tippett
